# Dietary Interventions and Cognitive Function across the Dementia Continuum: A Systematic Review, Meta-Analysis, Meta-Regression and Call to Action for Research Reform

**DOI:** 10.1101/2025.10.09.25337399

**Authors:** Megan Kirk, Oliver Canfell, Meysam Pirbaglou, Forhad Chowdhury, Megan Smith, Rachel Reid, Amy Hitchcock, Joanne Baudin, Brandon Chang, Heather Knight, Eva Lash, Jason Grant, Erin L. Bellamy, Joel Katz, Ivan Koychev, Kamaldeep Bhui

## Abstract

**Background:** Dementia cases are projected to rise to 153 million worldwide by 2050. Diet is a modifiable risk factor, and interventions may delay cognitive decline. No review has synthesised the full randomised controlled trial (RCT) evidence across all dietary interventions and dementia stages.

**Objective:** To critically appraise and synthesise evidence of dietary interventions on cognitive function across the dementia disease continuum (healthy, at-risk/preclinical, mild cognitive impairment, clinical).

**Methods:** Following PRISMA guidelines, seven databases and three trial registries were searched for RCTs from inception to 28 January 2025. Eligible RCTs evaluated multidomain (e.g., diet plus exercise), whole dietary pattern (e.g., Mediterranean diet), or single food (e.g., blueberries) interventions ≥ 2-weeks in adults, 18+ years. Data extraction was triple coded; risk of bias was assessed using the Cochrane Risk of Bias (RoB2) tool. Narrative synthesis was complemented by pooled random-effects meta-analysis. Prespecified meta-regression explored reasons for heterogeneity. Sensitivity analyses were performed.

**Results:** Eighty-three RCTs (110 diet intervention comparisons; *n* = 24 063 participants) met inclusion criteria. Overall, 190 cognitive measures were extracted and mapped onto the six DSM-V neurocognitive domains. A total of 885 cognitive, neuroimaging, and blood biomarker outcomes were assessed. Pooled meta-analysis (*k* = 35) showed a small, significant improvement in global cognition (Hedges’ *g* = 0.25, 95%CI: 0.15 to 0.36). Multidomain (*g* = 0.25, 95%CI: 0.10 to 0.41) and total diet (*g* = 0.27, 95%CI: 0.10 to 0.44) interventions showed benefit; single food interventions were non-significant. Trial durations ≤ 12 weeks (β = 0.59) and presence of cardiometabolic comorbidities at baseline (β = 0.45) predicted greater cognitive improvements in multidomain interventions; for total diet interventions, lower risk of bias (β = 0.51) moderated effects. Neuroimaging revealed benefits in MCI across all intervention types. Certainty of evidence was rated very low.

**Discussion:** Dietary interventions modestly improve cognitive function across dementia stages, with multidomain and total diet interventions showing the most benefit. RCTs during the preclinical phase (e.g., MCI) of dementia in adults with co-occurring cardiometabolic risk are recommended. Given substantial heterogeneity, risk of bias, and measurement inconsistency, international consensus is urgently needed to standardise intervention protocols (e.g., duration, type, dosage) and cognitive outcome measures to strengthen evidence for dementia prevention and care.

**Funding:** NIHR Applied Research Collaboration Oxford and Thames Valley, NIHR Oxford Health Biomedical Research Centre (BRC)

**Registration:** NIHR PROSPERO database (registration: CRD42023488336)

## Introduction

Dementia is a progressive and neurodegenerative clinical syndrome that affects 55 million people worldwide and is projected to exceed 153 million by 2050^1,2^. Dementia is an umbrella term for several diseases that impair memory, decision-making and thinking, mood and daily functioning, with Alzheimer’s disease (AD) as the most common form^3^. Dementia prevalence is unequally distributed across the world with over 60% of cases in low- and middle-income countries (LMICs)^4,5^, expected to rise to 71% in 2050^3^. In the UK, nearly 1 million people live with dementia and it has been the leading cause of death for the last decade^6,7^. The economic burden of dementia is substantial with estimated global healthcare costs reaching USD $9.2 trillion^8^, and NHS costs projected to triple from £23 billion per year to > £60 billion by 2040 making it the costliest disease condition exceeding cancer, heart disease and stroke^9^.

Once diagnosed, dementia is irreversible. There is currently no known effective treatment that prevents disease progression. Cholinesterase inhibitors and memantine are approved pharmacotherapies for the management of AD symptoms and demonstrate mixed effectiveness for improving patient symptoms across the dementia continuum, with most benefit gained from stabilising or slowing cognitive decline in AD^10,11^. Novel amyloid-β protein (Aβ) immunotherapeutic treatments that target amyloid plaques as the primary molecular driver of AD do not demonstrate consistent clinical improvements, with meta-analyses showing mixed efficacy^12^. Behavioural interventions targeting modifiable risk factors hold potential for dementia prevention, and improving quality of life, particularly when implemented early in the disease course.

The 2024 report from the *Lancet* Commission on dementia prevention identified elevated low- density lipoprotein (LDL) cholesterol as a new modifiable risk factor for dementia, adding to a collection of 14 risk factors that explain over 45% of dementia cases^13^. When compounded with existing risk factors of excessive alcohol consumption, hypertension, obesity, and diabetes, the addition of high LDL cholesterol as a new dementia risk factor reinforces the argument for behavioural interventions that include a dietary-based component to reduce dementia risk. The 2024 *Lancet* report calls for ambitious and coordinated efforts to advance dementia prevention, ranging from policy change to individually tailored, multicomponent interventions that address clusters of co-occurring modifiable risk factors influenced by diet, including LDL cholesterol and hypertension^13^. The World Health Organisation (WHO) 2020 Guidelines for Risk Reduction of Cognitive Decline and Dementia similarly emphasise multicomponent lifestyle-based approaches to target multiple co-occurring risk factors at the individual and population level^14^. Preventive interventions that target modifiable risk factors are needed to transition from a reactive ‘break-fix’ approach to care that focuses on dementia symptom management to a proactive model of ’predict- prevent’ care.

Evidence of the effects of dietary interventions on cognition remains fragmented. Published review articles are highly heterogenous across populations, interventions, and study designs, and conclusions are review-specific and do not permit statements about the overall strength of evidence to inform clinical practice and public health policy. To date, reviews have examined *individual dietary components*, including micronutrients and minerals (e.g., B vitamins^15–18^, vitamin E^16,18,19^, vitamin C^16^, vitamin D^16^, folic acid^16^, magnesium^20^, omega-3 fatty acids^21–23^, nitrate/nitrite^24^, (poly)phenols^25–28^), macronutrients (e.g., protein^29^, fibre^30^, glucose^31,32^, fat^33,34^) and single foods (e.g., fish^35,36^, olive oil^37,38^, blueberry^39^, nuts^40^) . Reviews of *dietary patterns* have focused mostly on the Mediterranean diet^41–49^, with others including Dietary Approaches to Stop Hypertension (DASH)^50,51^, Mediterranean-DASH (MIND)^42,50–54^, ketogenic^42,55,56^, Nordic^57^, caloric restriction and weight loss interventions^58–60^, and ultra processed foods^61,62^. Evidence is still emerging for *multidomain interventions with a dietary component* that target several mechanisms and risk factors, such as diet and physical activity, with or without cognitive training^63–68^. Reviews have typically focused on one or two populations across the dementia disease continuum, either targeting healthy^18,29,30,44,69^ or at-risk populations^64,65^, or those with mild cognitive impairment^18,32,49,68,70^, all-type dementia^53^, or Alzheimer’s disease^32,49,53,56,70^. Many review methods have been used, including narrative reviews^63,65^, systematic reviews without meta-analysis^16,19,41,42,45,48,50,51,53–55,57,61,68^, meta-analysis of observational studies^23,32,33,43,47,49^, meta-analysis of randomised controlled trials^15,18,56,64,66–69^, dose-response meta-analysis^21,70^, and network meta-analysis and mendelian randomisation^71^.

Despite evidence supporting a positive association between certain dietary patterns and cognitive function, most of the evidence is from narrative reviews restricted in scope and target population, without meta-analysis, or from meta-analyses of observational studies. Issues of confounding, reverse causation, and selection biases also limit causal inference^72^. Mendelian randomisation studies on diet are also limited due to a mismatch between observational and trial evidence, and a lack of consensus in classifying dietary patterns and the dietary dose-response relationship^70,72^. Thus, key uncertainties remain regarding the overall causal effect of diet on cognitive function, the most beneficial intervention components and dosage of food or dietary pattern, and at which disease stage(s) of dementia these interventions work best.

To our best knowledge, no published synthesis has comprehensively evaluated the entire body of evidence from randomised controlled trials (RCTs) to support definitive evidence statements of the effect of diet on cognition, from healthy individuals to diagnosed dementia. This study addresses this gap through the largest systematic review and meta-analysis of dietary interventions and cognition to date. Our primary research question was *what is the efficacy of dietary interventions on cognitive function?* Additionally, we examined subgroup effects of intervention type and duration, clinical disease stage, and specified sociodemographic factors to ascertain where effect size differences are most notable. The present global evidence synthesis with meta-analysis and meta-regression will contribute to a comprehensive empirical basis to support the 2024 *Lancet* Commission report, provide recommendations for conducting high-quality dietary intervention trials to support dementia prevention, and inform future clinical guidelines for the prevention and management of dementia.

## Methods

We followed the Preferred Reporting Items for Systematic reviews and Meta-Analysis (PRISMA) guidelines^73^. A protocol was preregistered with PROSPERO (ID: CRD42023488336).

### Information Sources and Search Strategy

A systematic electronic search was conducted across seven databases: CINAHL (EBSCOHost, 1982- present), Cochrane Database of Systematic Reviews (Cochrane Library, Wiley) Embase (OvidSP, 1974-present), MEDLINE (OvidSP 1946-present), PsycINFO (OvidSP 1806-present), Science Citation Index (Web of Science Core Collection, 1900-present), Scopus (Elsevier), and three trial registries: WHO ICTRP, ClinicalTrials.gov, and Cochrane Central Register of Controlled Trials (Cochrane Library, Wiley) from inception to January 23, 2024 for peer-reviewed RCTs on whole-food dietary interventions and cognitive function. A second duplicate search (24 January 2024 - 28 January 2025) retrieved 810 additional citations. Our search combined relevant keywords and MeSH terms, developed with a Primary Care librarian (see Supplementary Table S1). Studies were limited to the English language and peer-reviewed publications due to resource constraints.

### Eligibility Criteria

Eligible studies were RCTs (parallel-arm, cluster or factorial) involving adults aged 18 years and older. We defined dietary interventions as those targeting modifying a whole dietary pattern (e.g., Mediterranean diet) or increased consumption of a whole food component (including fortified or blended foods). Micronutrient supplements and medical foods were excluded. Dietary interventions were categorised into three types: total dietary pattern, daily intake of a single whole food, or a multidomain intervention with a dietary component. Studies with short-term (<14 days) or single dose interventions were excluded due to our focus on behaviour change^74^. Studies of adults at any stage of the dementia continuum (healthy, preclinical/prodromal, sub-clinical, clinical) were eligible. Studies focused on a primary condition other than cognitive impairment (e.g., cancer) were excluded. No restrictions were placed on sex. Eligible comparators included no intervention (e.g., waitlist control), placebo, usual care, or active controls. Settings could include in-patient, hospital, long-term care, community, at-home, and virtual or digital modalities.

### Cognitive Outcome Measures

Eligible studies must have reported at least one cognitive outcome measure. The primary outcome was change in a validated global cognitive function measure (e.g., Mini-Mental State Examination [MMSE]) from baseline to post-intervention. Studies assessing cognitive subdomains (e.g., memory, executive function) were also included. When available, brain imaging and blood biomarkers were extracted for exploratory analysis. Cognitive outcome measures were extracted and, where possible, the measurement validation studies were identified. When a study did not use and cite a validated measure, we assumed the measure was generated for the purpose of the study by the authors and treated it separately. For inclusion in the meta-analysis, studies required a quantitative, between-group, pre-post measure of global cognitive function. Only conceptually comparable cognitive measures were pooled to enhance the interpretability of the meta-analysis. Thus, studies that reported only composite z-scores derived from multiple cognitive subtests or test batteries were excluded from the meta-analysis. Composite scores vary considerably across studies in terms of the cognitive domains included and the weighting of subtests which introduces substantial heterogeneity.

### Taxonomy of Cognitive Outcome Measures

Two reviewers (MK, OC) independently reviewed each study, extracted all cognitive outcome measures, and mapped each onto the six DSM-V neurocognitive domains^75^: executive function, complex attention, social cognition, learning and memory, language, and perceptual-motor function to generate a working taxonomy. We searched the article’s reference section for validation studies for each cognitive outcome measure used to classify the cognitive domain(s) assessed by each measure. Accuracy checks between the two reviewers revealed, 87.4% agreement on coding cognitive outcome measures to the six DSM-V neurocognitive domains, with 144 discrepancies. Most discrepancies in coding occurred in complex attention (n = 41, 21.6%) and executive function (n = 35, 18.4%) due to variation in assessment methods (e.g., timed task vs. error rate) and domain overlap.

### Study Selection

Study selection was conducted using Covidence (2024)^76^. Four reviewers (MK, OC, EL, HK) independently screened titles and abstracts based on predefined inclusion and exclusion criteria. Interrater reliability was adequate (Cohen’s *kappa =* 0.65). Decision conflicts (*n* = 387, 6.5%) were resolved by consensus among three reviewers. Full-text studies were independently screened by the same four reviewers. Interrater reliability was adequate (Cohen’s *kappa* = 0.65) and discrepancies (*n* = 59,7.1%) were resolved by consensus among three reviewers.

### Data Collection Process

A standardised data extraction form was developed, *a priori*, using Jisc online survey software^77^ and pilot-tested with three reviewers (RR, AH, JC). Two rounds of pilot testing were conducted. First, a subsample of 10 eligible studies underwent blind extraction. Definitions and terms were revised based on initial feedback. Then, a new set of 20 eligible studies underwent a second round of blind data extraction. Responses were assessed for accuracy and completeness by two lead authors (MK, OC). Full data extraction then commenced among three reviewers (RR, AH, JC) using an excel spreadsheet. All included studies were triple extracted by three reviewers for precision and cross-checked by lead reviewers (MK, OC). Extracted data included study demographics, intervention, and outcome characteristics, extracted according to the CONSORT Guidelines^78,79^ in conjunction with the Template for Intervention Description and Replication (TIDieR) checklist^80^.

### Data Synthesis

A narrative synthesis was conducted using three pre-specified summary tables to synthesise study and intervention characteristics, cognitive outcome measures, and key findings. For interventions and outcomes, studies were grouped by dietary intervention type: multidomain, total diet, and single food. Only between-group differences in cognitive function at post-intervention were extracted and synthesised. As our focus was causal inference between diet and cognition, within- group changes were not extracted due to vulnerability to confounding over time.

### Risk of Bias Assessment

Risk of bias and study quality were assessed using the Revised Cochrane Risk of Bias (RoB2) Tool^81^. A set of two reviewers (RR, AH, JC, FC) independently rated each trial, with 20% of studies triple coded and verified by a lead reviewer (MK, OC). Trials were classified as having low, some concern, or high risk of bias across six domains: sequence generation, allocation concealment, blinding, completeness of outcome data, outcome measurement, and selective reporting.

### Meta-Analysis

All quantitative analyses were conducted using RStudio^82^ (version 2024.04.2), and the Meta-Analysis Package for R (metafor)^83^. Random-effects meta-analysis was conducted to calculate the Hedges’ *g* standardised mean difference (SMD) based on the difference of pre-post change in mean global cognitive function score between the intervention and control groups, divided by their respective standard deviation (SD) at baseline^84^. Considering that several studies reported the SD of pre-post change in global cognitive function, we also calculated the SMD according to the Skvarc and Fuller-Tyszkiewicz approach^85^. In this approach, Hedges’ *g* SMD is calculated as the pre-post difference between the intervention and control groups, divided by the SD of the pre-post change. Hedges’ *g* calculations were performed for all intervention comparisons combined, and separately for multidomain, total diet, and single food comparisons. When standard errors of the mean of 95% confidence intervals (Cis) were originally reported, baseline SD was calculated according to procedures specified by the Cochrane Collaboration^86^. We adopted similar procedures to estimate the SD or change from baseline to post-intervention SD, or the effective sample size in cluster RCTs^86^. For the primary analysis, we assumed a moderate correlation of *r =* 0.5 as a midpoint value to avoid over- or under-estimating the variance of SMD estimates. To assess the robustness of the findings to this assumption, all Hedges’ *g* SMD calculations were repeated using a range of correlations from 0.10 to 0.90. All comparisons were expressed as pooled SMD with 95% Confidence Intervals (CIs). Effects were considered significant at *p* <.05. Hedges’ *g* calculations were interpreted based on prior recommendations: 0.2 = small effect, 0.5 = moderate effect, 0.8 = large effect^87^. Forest plots were presented to illustrate results.

Heterogeneity was evaluated using the Cochran’s Q statistic and *I*^2^ to quantify the percentage of variation. Publication bias and small study effects were assessed via funnel plots and trim-and-fill analysis using Egger’s test. We performed subgroup analysis of pre-specified intervention or participant-related subgroups using mixed-effects meta-regression.

### Mixed-Effects Meta-Regression

Meta-regression analysis was performed to explore sources of heterogeneity using mixed-effects models with maximum-likelihood estimation. For each prespecified moderator, regression models estimated the regression coefficient (β) and 95% CIs to quantify the change in effect size relative to the reference category (R). The omnibus test statistic for moderators (QM) and *p*-value were used to assess whether the moderator explained significant variability in effect sizes. Statistical significance of regression coefficients indicated the direction and magnitude of effect on cognitive function for specific moderator variables. Heterogeneity was reported using the *I²* statistic and Cochran’s *Q* statistic, respectively. The proportion of between-study variance explained by each moderator was reported using *R*^2^.

### Assessment of Minimal Clinically Important Difference (MCID)

To compare mean change scores in global cognitive function based on cited minimal clinically important difference (MCID), between-group mean difference scores for each intervention comparison were calculated (*M* intervention_T1_ – *M* intervention_T0_) – (*M* Control_T1_ – *M* Control_T0_). MCID for MMSE equates to a mean change of 1.4 (SD = 0.4) points^88^, ADAS-Cog mean change score of 2.6 (SD = 0.4)^88^, and MoCA mean change score of 2.15 (SD = 0.5) points^89^ and have been cited as clinically significant.

### Certainty Assessments

Three reviewers (FC, MK, MP) independently assessed the certainty of the evidence using the Grading of Recommendations Assessment, Development and Evaluation (GRADE) tool. Discussion with a fourth reviewer (OC) was completed until consensus was reached. GRADE evaluates the certainty of evidence across multiple domains including study design, risk of bias, inconsistency of results, indirectness, imprecision, and publication bias.

### Patient and Public involvement

This study was shared with the Oxford Health Biomedical Research Centre (BRC) Preventing Multiple Morbidities Theme Patient and Public Involvement (PPI) Panel in partnership with the McPin Foundation. The PPI Panel consists of 12 members, aged 19-70 years, 75% women (*n* = 9), and 50% (*n* = 6) ethnic minority with coexisting mental and physical health conditions, carer and patient experience, neurodiversity representation, and inpatient and outpatient experiences. Feedback from the PPI Panel indicated the topic of diet and cognition was a high priority. Two PPI members elected to provide written, verbal, and lived experience perspectives on the construction of the final manuscript and attended two writing retreats (July 2024 and July 2025). An additional member of the public, a practicing clinician (EB), was invited to contribute to the manuscript. Suggested revisions included clarifying definitions of the intervention types and outcome measurement summary, revisions to the introduction section, and a PPI “call to action” to discuss the implications of the findings and the issues researchers should consider from a lived-experience and public perspective.

## Results

### Database Search Results

Figure 1 shows the PRISMA flow diagram of the search results. A total of 17,727 citations were identified across CINAHL (*n* = 2141), ClinicalTrials.gov (*n* = 1043), Cochrane Central Register of Controlled Trials (*n* = 2638), Embase (*n* = 3559), MEDLINE (*n* = 4117), PsycINFO (*n* = 882), Science Citation Index (*n* = 750), Scopus (*n* = 1995), WHO ICTRP (*n* = 590), and other sources *(n* = 12). After removing 11,778 duplicates and ineligible records, 5,949 titles and abstracts were screened. Of these, 831 full-text articles were retrieved and assessed for eligibility of which 739 studies were excluded. Overall, 92 publications from 83 RCTs (110 intervention arm comparisons; 24,063 participants) met inclusion criteria. Secondary publications of main trials are listed in Supplementary Table S2. A total of 35 intervention comparisons from 25 studies (30.1%) provided sufficient data for meta-analysis. Authors of studies with missing data were contacted to request missing data and five (14.7%) shared additional data. Reasons for missing data from authors included non-response, the requested data was unavailable, inactive corresponding author email, maternity leave, or too busy.

**Figure 1.**
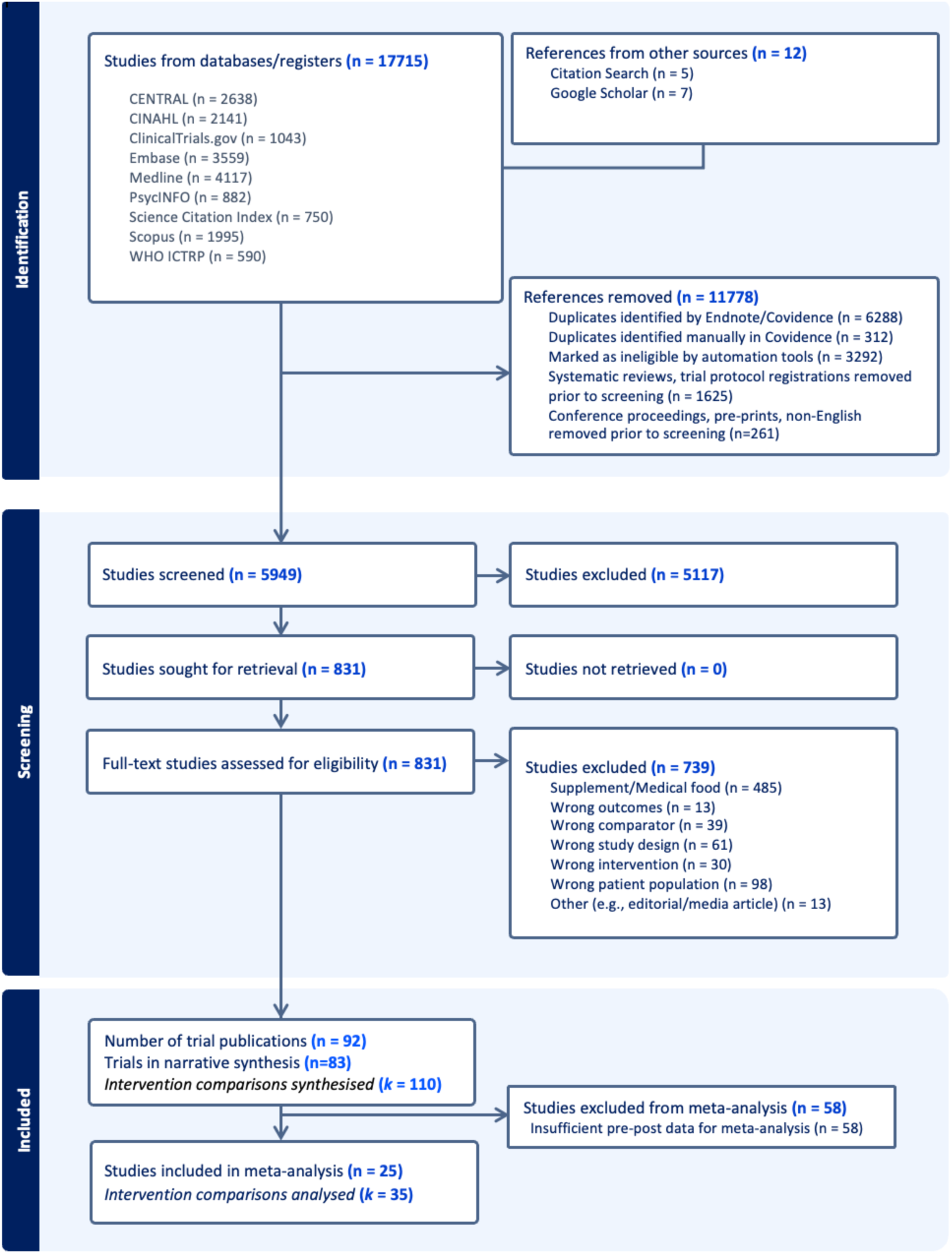
PRISMA flow diagram of systematic database searches, screening and studies included.

### Study Characteristics and Demographics of included RCTs (*n* = 83)

Supplementary Table S3. outlines the descriptive characteristics of the 83 RCTs^90–172^. Most trials (*n* = 36, 43.4%) were conducted in North America (USA: *n* = 34, Canada: *n* = 1, Mexico: *n* = 1) followed by 21 studies (23.3%) in Europe and Central Asia (EU: *n* = 10, UK: *n* = 6, Finland: *n* = 2, Denmark: *n* =1, Iceland: *n* = 1, Norway: *n* = 1), 19 studies (22.9%) in East Asia and Pacific (Australia: *n* = 8, Japan: *n* = 3, South Korea: *n* = 2, Taiwan: *n* = 2, China: *n* =1, Malaysia: *n* = 1, Indonesia: *n* = 1, Thailand: *n =* 1), three studies (3.6%) in Middle East and North Africa (Iran: *n* = 2, Tunisia: *n* = 1), and two studies (2.4%) in South Asia (India: *n* =1, Pakistan: *n* =1) and Latin America (Brazil: *n* = 2, 2.4%), respectively (Figure 2). Most studies (n = 72, 86.7%) were conducted in high income countries, with 8 (9.6%) conducted in upper-middle income countries, and only 3 (3.6%) conducted in low-middle income countries (LMICs).

**Figure 2.**
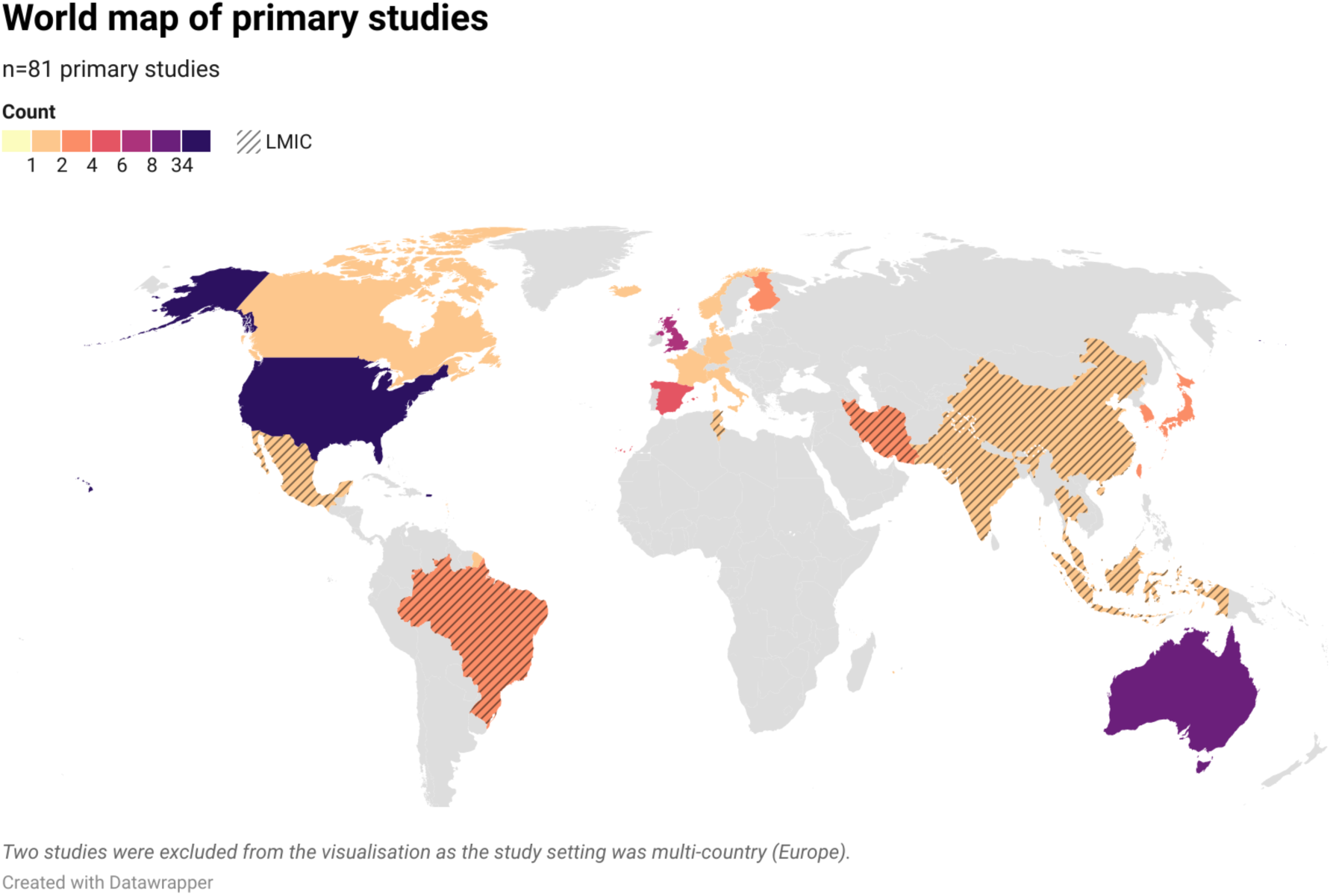
Geographic representation of primary studies included in the systematic review (k = 83)

Of the 24,063 participants enrolled across the included studies, 18,266 completed the RCT, yielding an overall study completion rate of 75.7%. Sample sizes ranged from 13^133^ to 6104^102^. The overall mean age of participants was 63.3 years (SD = 14.7). Eighty-one (97.6%) studies reported sex with an average of 61.2% female participants (range = 0% to 100%). More than half of studies did not report ethnicity of study participants. Where reported, 20.7% of participants were from a non-White background. Most studies included cognitively normal, healthy participants (*n* = 49, 59.0%), followed by diagnosed MCI (*n* = 17, 20.5%), at-risk participants (*n* =10, 12.4%), and diagnosed Alzheimer’s disease (*n* = 1, 1.2%). Six studies (7.2%) reported blended samples, either cognitively normal/MCI (n = 2), or MCI/Alzheimer’s disease (*n* = 4). A total of 30 studies (36.1%) examined participants with a co-occurring condition, the most common being overweight/obesity *(n* = 20, 24.1%).

### Intervention Characteristics (*k* = 110)

The intervention characteristics across 83 trials are summarised in Supplementary Tables S4a-S4c. A total of 110 diet-based intervention arm comparisons were summarised: multidomain intervention arms (*k* = 35); total diet intervention arms (*k* = 39); single food intervention arms (*k* = 36).

#### Multidomain Interventions (n = 30, k = 35)

Thirty studies^90,96,102,107,110,114,116,121,123,126,127,134,135,138,140,142,143,147–151,156,159,163–166,171,172^ examining 35 multidomain intervention arm comparisons were evaluated (Supplementary Table S4a). Attrition rates were higher in intervention groups (32.1%, baseline *n* = 7028; post-intervention *n* = 4775) than controls (24.1%, baseline *n* = 6290; post-intervention *n* = 4771). Most studies (n = 16, 53.3%) enrolled cognitively normal participants at baseline^102,107,110,114,116,121,123,126,127,135,138,140,143,149,163,171^, with others including at-risk (n = 5, 16.7%)^90,135,147,150,172^, mild cognitive impairment (MCI) (n = 6, 20.0%), ^96,142,148,159,164–166^, or blended samples with MCI or early-stage Alzheimer’s disease (n = 3, _7.2%)_^134,151,156^.

The most common dietary interventions were National Nutrition Guidelines (NNG) (n = 9, 30.0%)^90,114,127,135,143,150,159,165,172^ and the Mediterranean Diet (MEDI) (n = 7, 23.3%) ^102,107,110,116,121,142,166^, followed by MIND (n = 3, 10%)^134,147,156^ and DASH (n = 3, 10%)^96,163,164^. Less frequent dietary interventions were also reported: caloric restriction^138,149^, high-fibre diet^140,171^, intermittent fasting^123^, a vegan-based diet^151^, and a study-generated diet approach^148^ . Most studies (n = 28, 93.3%) provided guided nutrition support, with two offering partial support either through automated SMS text message^102^ or minimal guidance at baseline and end of study only^116^.

All 30 studies combined diet with up to six additional components. Components varied widely and included aerobic exercise, resistance training, or combined exercise, cognitive training, social engagement, stress management techniques, vascular risk management, GP consultations, goal setting, and/or therapy sessions. Intervention duration ranged from 8 weeks^110^ to 208 weeks (4 years)^127^, with half (n = 15, 50.0%) lasting 12-24 _weeks_^96,107,114,116,121,123,126,138,147,148,151,163–165,171^, and others spanning <12 weeks (n = 5, 16.7%)^110,134,140,142,143^, 1 year (n = 4, 13.3%)^102,135,149,166^, or >1 year _(n = 6, 20%)_^90,127,150,156,159,172^.

Control conditions included passive controls (n = 13, 43.3%) such as placebo, minimal/no intervention^90,110,116,123,126,127,138,149,163–166,171^, active controls (n = 7, 23.3%) including attention- matched and low-intensity active comparators ^96,102,107,121,135,142,148^, waitlist controls (n = 5, 16.7%) either alone or combined with TAU ^134,140,143,147,151^, and treatment as usual (TAU) (n = 5, 16.7%)^114,150,156,159,172^, including one study where general health education was delivered as part of usual care^172^.

#### Total Diet Interventions (n = 33, k = 39)

Thirty-three studies^91,93–96,101,103,109,113,116–121,123,125,127,129,136–139,149,155,162–164,166–169,171^ evaluating 39 total diet intervention comparisons were included (Supplementary Table S4b). Attrition rate was equivalent in intervention groups (17.2%, baseline *n* = 4094; post-intervention *n* = 3389) compared to control groups (17.6%, baseline *n* = 3638, post-intervention *n* = 2998). Most studies (n = 25, 75.8%) enrolled cognitively normal participants^91,93–95,101,109,113,116,119,120,123,125,127,136–139,149,155,162,163,167–169,171^, followed by MCI (n = 6, 18.2%) ^95,96,117,129,164,166^, and blended samples (n = 2, 6.1%), either cognitively normal/MCI^118^ or MCI/early AD^103^.

Thirteen unique dietary patterns were explored. The most common was the Mediterranean diet (n = 9 studies, 33.3%; n = 14 intervention arm comparisons, 39.4%)^116,118,121,125,137,139,166,168,169^, followed by a ketogenic-based diet (n = 5, 12.8%) including the Atkins diet or very low carbohydrate/high fat protocol;^101,103,113,129,136^ and caloric restriction (CR) diet (n = 5, 12.8%)^117,138,149,162^, intermittent fasting (n = 4, 10.3%)^94,120,123,155^, DASH (n = 3, 7.7%)^96,163,164^, low fat (n = 2, 5.1%)^109,169^, MIND with CR (n = 2, 5.1%)^91,93^, NNG (n = 2, 5.1%)^95,127^, study-generated custom diets (n = 2, 5.1%)^155,167^, high fibre (n = 1, 2.6%)^171^, high protein (n = 1, 2.6 %)^119^, or low-calorie liquid diet (n = 1, 2.6 %)^138^. Intervention durations ranged from 3 weeks^119^ to 442 weeks (8.5 years)^109^. Most intervention durations were between 12-24 weeks (n = 13, 39.4%)^91,95,103,116,121,123,125,138,155,163,164,169,171^, with others spanning less than 12 weeks (n = 5, 15.2%)^113,118–120,129^, 24-52 weeks (n = 9, 27.3%)^94,96,101,117,136,137,149,166,167^, and >1 year (n = 6, 18.2%)^93,109,127,139,162,168^. Most studies provided guided nutrition support (n = 30, 91%), with three studies providing no support^94,116,119^.

Control conditions varied and included passive controls (n = 16, 48.5%)^94,96,109,116,117,120,121,123,125,127,137,149,155,162–164^, active controls (n = 13, 39.4%), ^91,93,101,103,113,118,119,129,136,138,139,166,168^. waitlist controls (n = 2, 6.1%),)^169,171^, and TAU (n = 2, 6.1%)^95,167^.

#### Single Food Interventions (n = 31, k = 36)

Thirty-one studies^92,97–100,104–106,108,111,112,115,122,124,128,130–133,141,144–146,152–154,157,158,160,161,170^ examining 36 single food intervention arm comparisons were synthesised (Supplementary Table S4c). Attrition was lower in the intervention arm (13.1%, baseline *n* = 1278; post-intervention *n* = 1118) compared to the control arm (17.3%, baseline *n* = 1237; post-intervention *n* = 1023). Most studies enrolled cognitively normal participants (n = 22, 71.0 %)^92,98–100,105,108,111,122,124,131,132,141,144–146,152–154,157,158,160,170^, followed by MCI (n = 8, 25.8%)^97,104,112,115,128,130,133,161^ and Alzheimer’s disease (n = 1, 3.2%)^106^.

Single food interventions were categorised into four groups*: Fruits and Vegetables, Nuts and Seeds, Oils and Fats, and Meat and Alternative Protein.* Most interventions tested *Fruits and Vegetables* (*n* = 21, 67.7%)^92,97–100,105,108,112,124,128,130–133,144–146,157,158,161,170^. Of these, most (n = 20, 95.2%) examined fruit consumption, primarily blueberries, with only one study investigating vegetable consumption (beetroot)^92^. Seven interventions (22.6%) investigated *Nuts and Seeds* (e.g., almonds, walnuts, peanuts, and Brazil nuts)^104,111,122,152–154,160^, followed by *Oils and Fats (*n = 2, 6.5%) like coconut oil or extra virgin olive oil consumption^106,141^, and *Meat and Alternative Protein,* where one intervention examined a meat alternative, tempeh^115^.

Intervention durations ranged from 4-weeks^99,146^ to 104 weeks (2-years)^160^. Four studies (12.9%) assessed trials <12 weeks^98,99,122,146^, 24 studies (77.4%) evaluated trials between 12-24-weeks (6-months)^92,97,100,104–106,108,111,112,115,124,128,130–133,144,145,152–154,157,158,170^; two studies (6.5%) examined 52- week (1-year) trials^141,161^, and only one study^160^ (3.2%) had a 2-year intervention period. Most intervention arms (*n* = 26, 83.9%) did not include guided support. Dietitian support was included in in five studies (16.1%)^99,111,141,157,160^. Most studies used a placebo control (n = 20, 64.5%)^92,97–100,105,106,108,112,124,128,130–133,144,145,158,161,170^, followed by an active control (n = 6, 19.4%)^111,115,122,141,152,153^, and passive control (n = 5, 16.1%)^104,146,154,157,160^.

### Taxonomy of Cognitive Outcome Measures

A total of 190 unique cognitive outcome measures were extracted, reflecting high variability in outcome measurement. Twenty-five cognitive outcome measures (13.2%) assessed four or five of the six DSM-V neurocognitive domains and were classified as measures of global cognitive function (Figure 3). The most frequently used validated global cognitive function outcome measures were the Mini-Mental State Examination (MMSE; *n* = 22, 23.9%), Montreal Cognitive Assessment (MoCA; *n* = 10, 10.9%), and Clinical Dementia Rating (CDR; *n* = 8, 8.7%). Sixteen studies (17.4%) used a study generated composite score to represent global cognitive function. None of the cognitive outcome measures evaluated all six DSM-V neurocognitive domains. The most common DSM-V neurocognitive domains assessed by cognitive outcome measures were learning and memory (*n* = 119, 62.6%), followed by complex attention (*n* = 90, 47.4%), executive function (*n* = 88, 46.3%), perceptual-motor function (*n* = 81, 42.6%), language (*n* = 33, 17.4%), and social cognition (*n* = 5, 2.6%) (Supplementary File Table S4). The remaining 165 cognitive outcome measures were classified as measures of non-global cognitive function (≤ 3 DSM-V neurocognitive domains) with most of these measures mapping to two DSM-V neurocognitive domains (n = 72, 37.9%), a single DSM-V neurocognitive domain (*n* = 59, 31.1%), or three DSM-V neurocognitive domains (*n* = 34, 17.9%), respectively. Measures of non-global cognitive function were summarised as single-domain cognitive outcome measures (Supplementary File Table S5).

**Figure 3.**
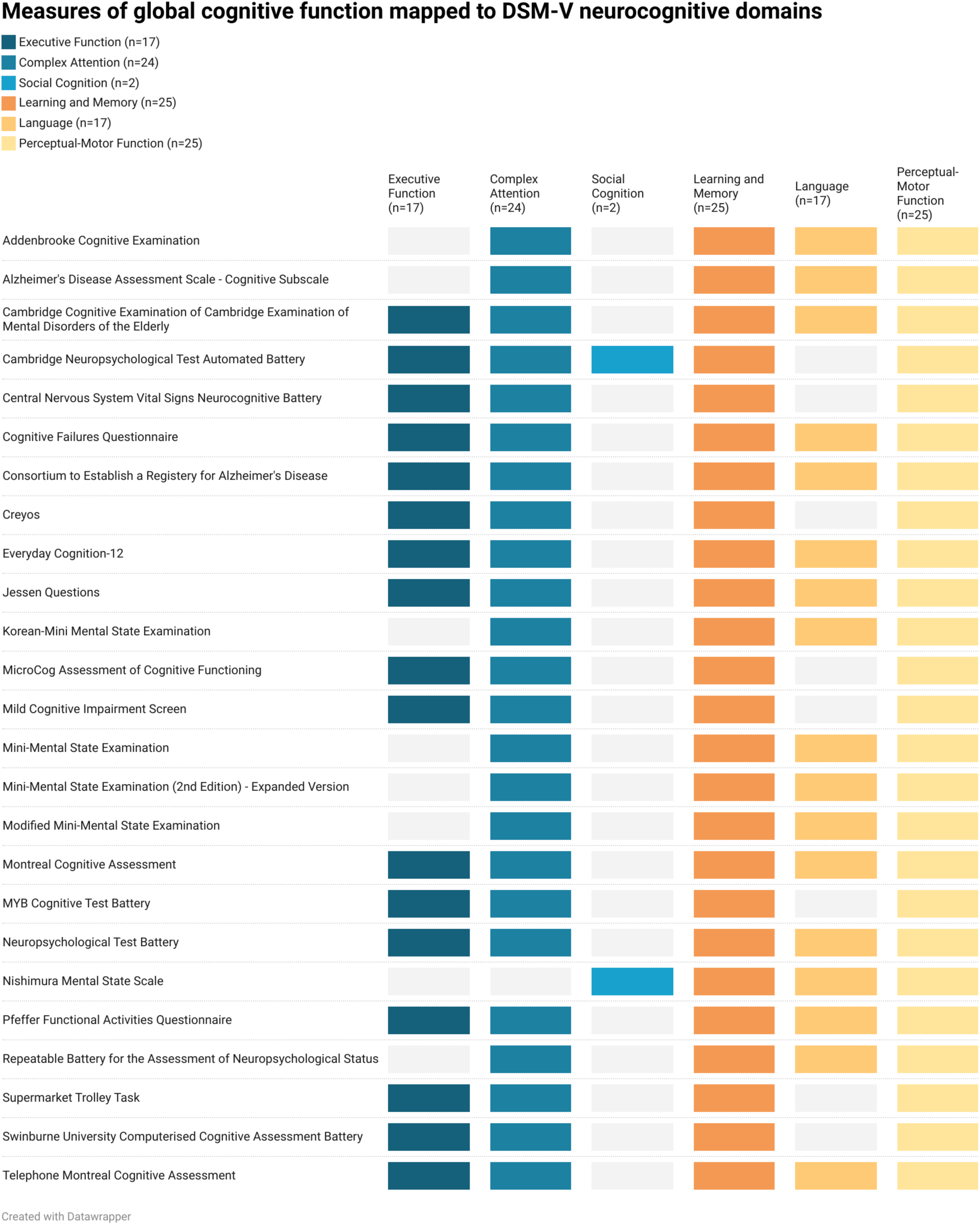
Measures of global cognitive function mapped onto the DSM-V six neurocognitive domains.

### Summary of Study Outcomes

Supplementary Tables 6a-c summarises the intervention effects on a total of 885 cognitive outcome measures of interest including our primary outcome, global cognitive function (*k* = 79, 7.9%), single- domain cognitive function (*k* = 457, 51.6%), brain imaging (*k* = 59, 6.7%), and blood biomarkers (*k* = 299, 33.8%).

### Primary Outcome: Global Cognitive Function

Across 47 (56.6%) studies, 79 global cognitive function (GCF) outcomes were reported (Supplementary Tables 6a-6c). Of these, 57.0% (45/79) showed no significant between-group differences at post-intervention, while 31.6% (25/79) favoured the intervention, mostly from multidomain studies (*k* = 17, 68.9%). Nine outcomes (11.4%) from seven studies were mentioned in the methods but not reported^97,112,115,121,127,133,156^.

#### Multidomain Interventions

Thirty-six GCF outcomes were reported across 21 multidomain studies^90,93,96,102,110,114,116,121,123,127,135,142,147,149–151,156,159,163,165,172^. Three studies did not provide between-group findings^121,127,156^. Of the remaining, 51.5% (17/33) showed significant post- intervention benefits for the intervention group compared to controls while 48.5% (16/33) did not. Cognitive status did not appear to influence outcomes. Non-significant results were similar in cognitively normal (68.6%) and cognitively impaired (MCI/AD) (63.6%) groups. Likewise, significant effects were comparable between normal (31.4%) and impaired (MCI/AD) (36.4%) groups. Results varied by the cognitive outcome measure used (Supplementary Table S6a).

#### Total Diet Interventions

Twenty-seven GCF outcomes were reported across 20 total diet studies^93–96,103,109,116–121,123,127,137,139,149,163,167,168^. Most (n = 21, 77.8%) showed no significant between-group differences at post-intervention (see Table 3b), while 18.5% (5/27) favoured the intervention. One study did not report between-group post-intervention results^121^. Findings varied by diet type and cognitive measure. Cognitive status had no apparent influence on outcomes (Supplementary Table S6b).

#### Single Food Interventions

Sixteen GCF outcomes were reported across 12 single food studies ^92,97,104,106,112,115,122,133,141,154,157,160^. Half (8/16) showed no significant between-group differences, while 18.8% (3/16) favoured the intervention. Five outcomes from four studies were not reported. ^97,112,115,133^. Results appeared to vary by cognitive measure. Studies using MoCA^122,157^ showed significant improvements in favour of the intervention group, while four studies using MMSE reported no effect^106,112,141,157^. Cognitive status did not appear to influence outcomes (Supplementary Table S6c).

### Non-Global, Single-Domain Cognitive Function Outcomes

There were 457 single-domain cognitive outcome results from 71 studies (85.5%). Most (k = 296, 64.8%) showed no significant post-intervention differences, while 19.7% (k = 90) favoured the intervention. Sixty-nine (15.1%) outcomes from 12 studies were mentioned in the methods but not reported ^101,103,113,115,117,121,144,145,155,158,164,166^. Two outcomes favoured the control condition^105,169^.

One study linked adverse effects in the intervention group to larger reductions in cholesterol^169^, and another reported high attrition (45%) due to GI issues^106^. Cognitive status did not appear to influence results. Non-significant findings were similar across cognitively normal (76.2%) and cognitively impaired (MCI/AD) (76.5%) groups, with comparable rates of significant effects among the same groups (Normal = 23.1% vs. MCI/AD 23.5%). Intervention type appeared to influence outcomes. Multidomain interventions had the highest proportion of significant between-group differences (33.7%) compared to total diet (20.2%) and single food (19.5%). Both total diet (78.8%) and single food interventions (80.1%) had a majority of non-significant results (Supplementary Table S6a-c)

### Brain Imaging

Sixty-five brain imaging results were extracted across 14 studies (multidomain *n* = 2, total diet *n* = 3, single food *n* = 9). Of the 59 reported results, most were from functional magnetic resonance imaging [fMRI/MRI] (n=43, 72.9%), followed by electroencephalogram[EEG] (n=6, 10.2%), quantitative and functional near infrared spectroscopy [qNIRS, fNIRS] (n=6, 10.2%), positron emission tomography [PET] (n=3, 8.8%) and transcranial Doppler ultrasound (n=1, 2.9%). Six results (9.2%) from three studies^97,133,134^ were mentioned in the methods but not reported. Overall, 54.2% (32/59) brain imaging results showed significant post-intervention effects favouring the intervention group, while 45.8% (27/59) were non-significant Brain regions assessed varied, with the hippocampal region (n=5), gyrus (n=4), cerebral blood flow (n=4), cortical thickness/volume (n=3), parietal cortex (n=3), and thalamus (n=2) most commonly reported. Cognitive status appeared to influence outcomes. Participants with cognitive impairment (MCI/AD) showed a much higher proportion of significant positive effects in favour of the intervention (91.7%) compared to cognitively normal participants (28.6%). Intervention type did not appear to influence results (Supplementary Tables S6a-c).

### Blood Biomarkers

A total of 345 blood biomarker results were extracted (see Tables 3a-c) across 43 studies. Forty-six blood biomarker results (13.3%) from nine studies were mentioned in the methods but not reported^99,104,106,113,117,127,140,165,167^. Most blood biomarker results (216/299, 72.2%) showed no significant between-group differences at post-intervention, while 27.4% (82/299) favoured the intervention, most commonly showing improved cholesterol (n=17), insulin (n=11), AD pathology biomarkers (n=5), and BDNF (n=3). One result (fasting insulin) favoured the control group, attributed to sugar intake from the prescribed fruit^128^. Cognitive status may have influenced outcomes.

Participants with cognitive impairment showed more significant effects (31.0%) and fewer non-significant results (68.4%) than cognitively normal participants (23.6% and 76.4%, respectively). Intervention type appeared to have no influence (Supplementary Tables 6a-c).

### Risk of Bias

#### Multidomain intervention comparisons

The Cochrane RoB2 assessment indicated that most multidomain studies performed well methodologically (Supplementary Figure S1). Bias was generally “low”, or with “some” risk of bias attributed to randomisation, adherence, missing data, measurement, and reporting. Overall, 25% of study comparisons were low overall risk, and 44.4% had “some” concerns (Figure 4a). A small proportion of studies (30.6%) were rated high-risk mainly attributed to missing data or measurement concerns^90,107,114,116,121,127,140,148^ (Supplementary Figure S1).

**Figure 4a.**
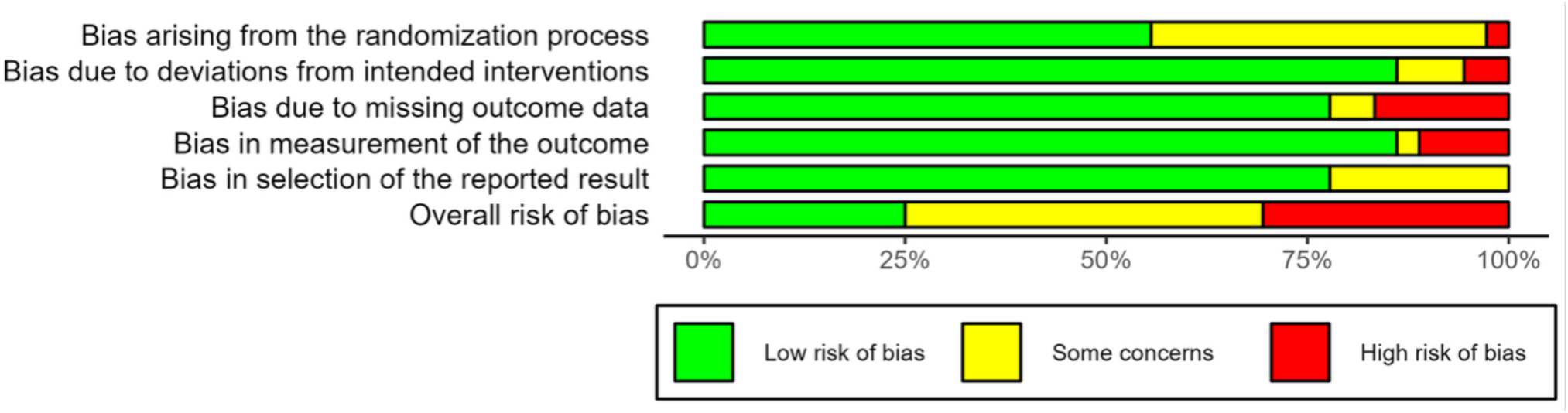
Overall Risk of Bias for Multidomain studies.

#### Total diet intervention comparisons

Total diet interventions showed higher bias than multidomain interventions with most studies having at least “some” concerns (Figure 4b). Primary issues were due to randomisation and outcome measurement, as well as bias from missing outcome data and deviations from intended interventions (Supplementary Figure S2). For overall risk, nearly 33.3% of the study comparisons showed “high” risk of bias, and 58.3% showed “some concerns” of bias. Only 10.4% of intervention comparisons were rated as “low” risk of bias^96,155,166^, highlighting more variability in study quality.

**Figure 4b.**
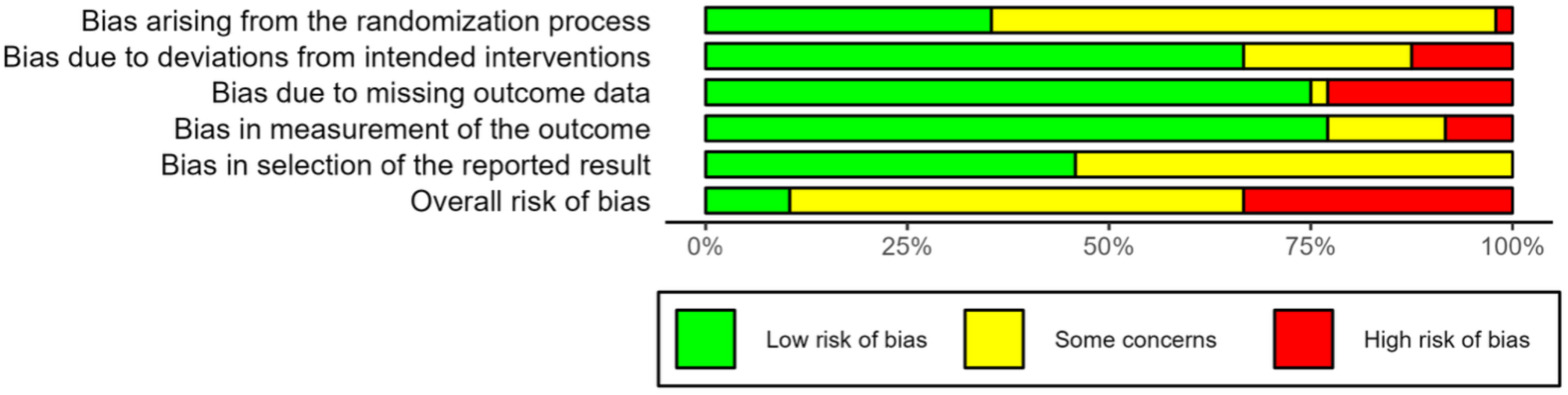
Overall Risk of Bias for Total Diet studies.

#### Single food intervention comparisons

Single food interventions showed the highest risk of bias (Figure 4c). Across all domains, most studies had “some concern” or “high” risk of bias mainly from randomisation, missing outcome data, and protocol deviations (Supplementary Figure S3). Overall, over half (55.6%) of the comparisons were rated “high” risk of bias, and 41.7% showed “some concerns” of bias. Only one study was classified as overall “low” risk of bias^124^ highlighting substantial methodological challenges in single food intervention studies.

**Figure 4c.**
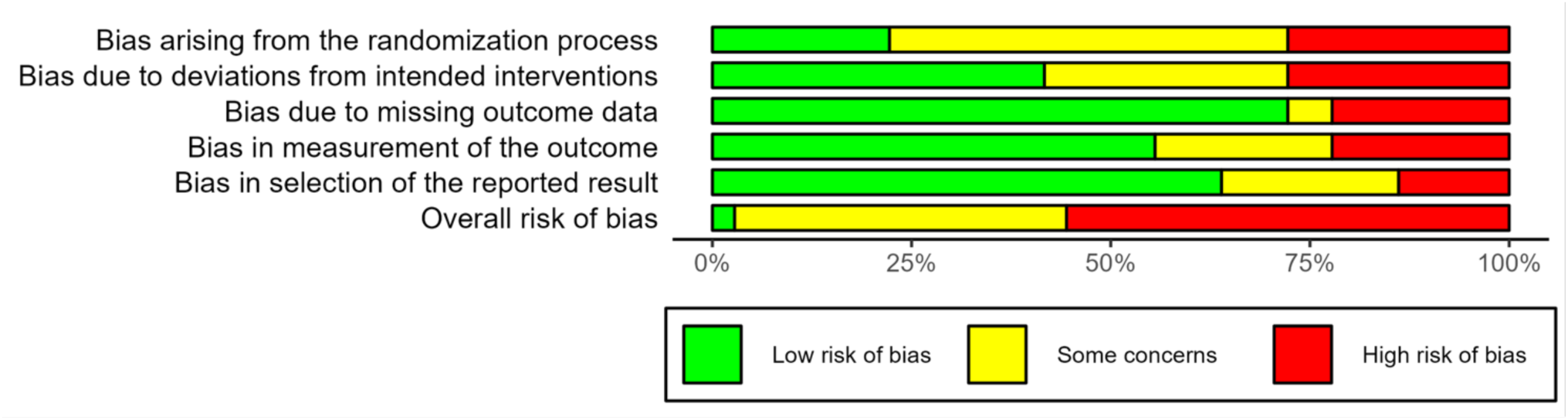
Overall Risk of Bias for Single Food studies.

#### Cluster RCTs

Four cluster RCTs^110,126,135,172^ were assessed with the Cochrane RoB2 tool for cluster trials (Supplementary Figure S4). All showed low risk of bias for timing of randomisation, handling of missing data, and outcome measurement. However, most cluster RCTs had “some concern” of bias for the randomisation process resulting in an overall “some concern” for risk of bias (Figure 4d).

**Figure 4d.**
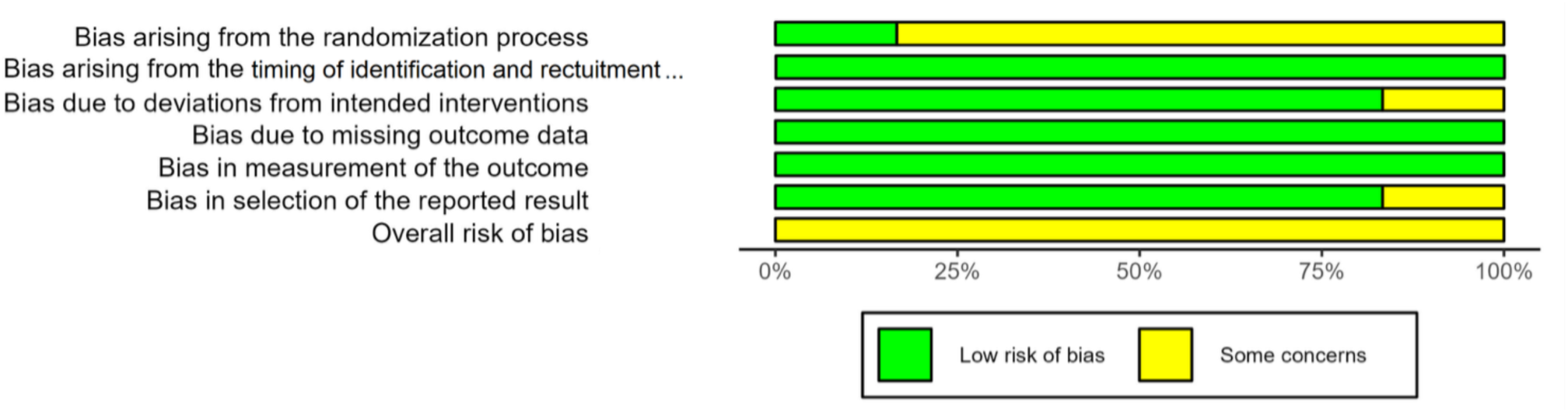
Overall Risk of Bias for Cluster RCT studies.

## Meta-Analysis Results

### Overall Main Effects (*k* = 35)

A meta-analysis of 35 dietary intervention comparisons from 25 studies was conducted using a random-effects model (residual maximum likelihood (REML) estimator). As seen in Figure 5, the pooled, adjusted effect size showed a statistically significant, small positive effect (Hedges’ *g* = 0.25; 95% CI: 0.15-0.36) for dietary interventions on global cognitive function (*z* = 4.87, *p* <.0001). There was moderate heterogeneity (*I*^2^ = 40.9%) suggesting the effect sizes were due to heterogeneity rather than sampling error. The Q-test for heterogeneity was significant (*Q*(34) = 67.74, *p* = .0005) indicating considerable between-study variability. The Egger test for publication bias showed significant funnel plot asymmetry (*t* = 3.35, *df* = 33, *p* = 0.002) and potential small-study effects or publication bias (Supplementary Figure S5). Sensitivity analyses were conducted with correlations between 0.10 to 0.90. All resulting effect sizes (range: *g* = 0.17 to 0.33) fell within the 95% confidence interval of the primary analysis, indicating the overall findings were robust to variations in the assumed pre-post correlation of 0.5 (Supplementary Table S7).

**Figure 5:**
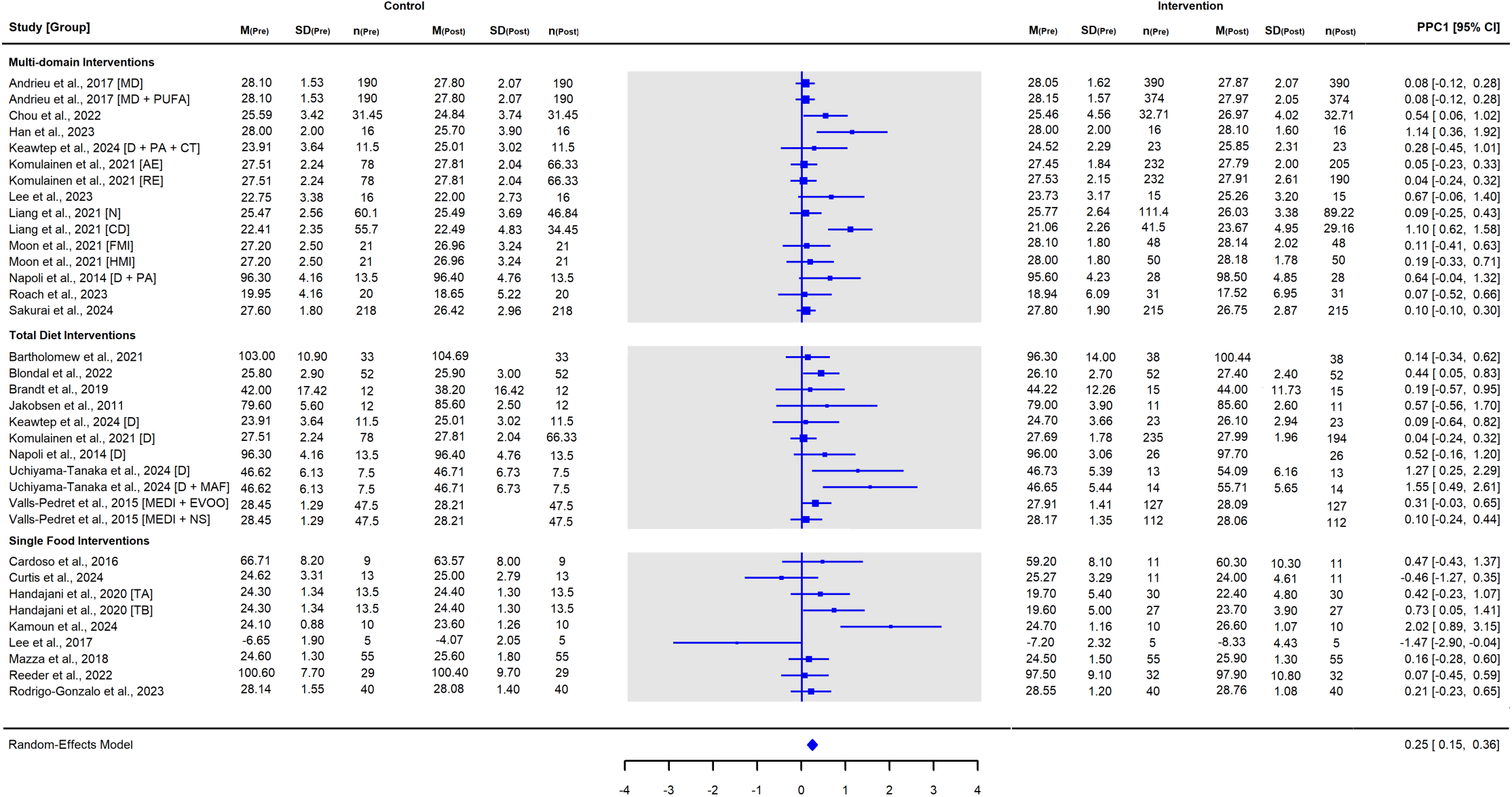
Meta-Analysis Forest Plot of the Pooled Effects of Dietary Interventions (k = 35) on Global Cognitive Function.

### Meta-Analysis for Multidomain Interventions (*k* = 15)

As seen in Figure 6, meta-analysis of 15 multidomain dietary intervention comparisons showed a small, statistically significant improvement in global cognitive function in favour of the intervention (Hedges’ *g* = 0.25, 95%CI: 0.10-0.41, *p* = .002). There was substantial heterogeneity (*I*^2^ = 67.5%) among included studies suggesting variability in effects across studies. The *Q* statistic was significant (*Q*(14) = 32.4, *p* = .004) further indicating notable between-study variability. Egger’s test for publication bias was significant (*t* = 3.58, *p* = .003) suggesting funnel plot asymmetry (Supplementary Figure S6) and likelihood of small-study effects and potential publication bias.

**Figure 6:**
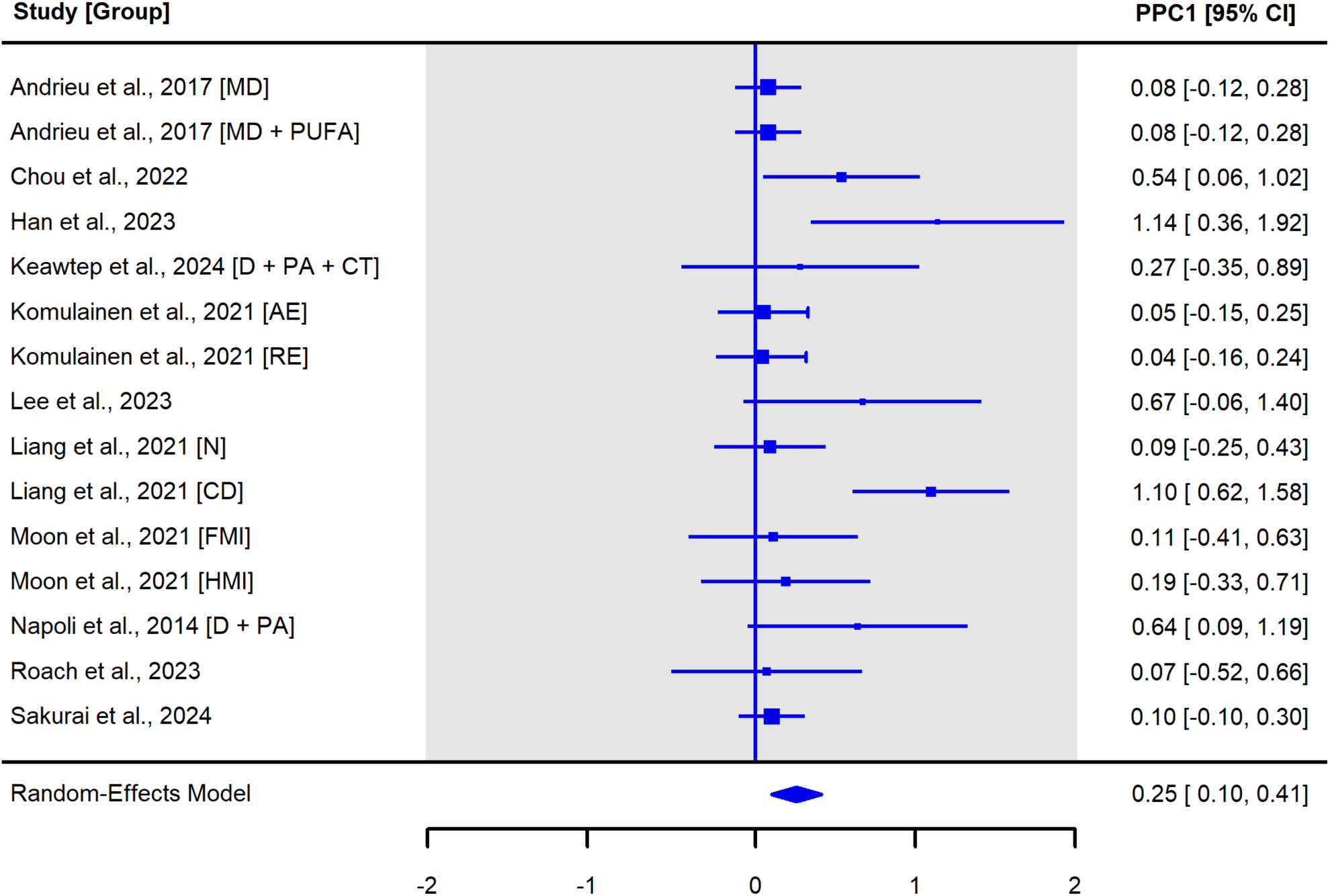
Meta-Analysis Forest Plot of the Pooled Effects of Multidomain Dietary Interventions (k = 15) on Global Cognitive Function.

### Meta-Analysis for Total Diet Interventions (*k* = 11)

As seen in Figure 7, meta-analysis of Total Diet intervention comparisons (k = 11) showed a significant small positive effect (Hedges’ *g* = 0.27, 95%CI: 0.10-0.44, *p* = .0021) on global cognitive function in favour of the intervention group. There was low heterogeneity across studies (*I*^2^ = 30.6%). The Q-statistic was borderline insignificant (*Q*(10) = 17.51, *p* = 0.06) suggesting a possibility of some variability in effect sizes across studies. Egger’s test indicated significant funnel plot asymmetry (*t* = 3.41, *p* = .008) suggesting potential for publication bias (Supplementary Figure S7).

**Figure 7:**
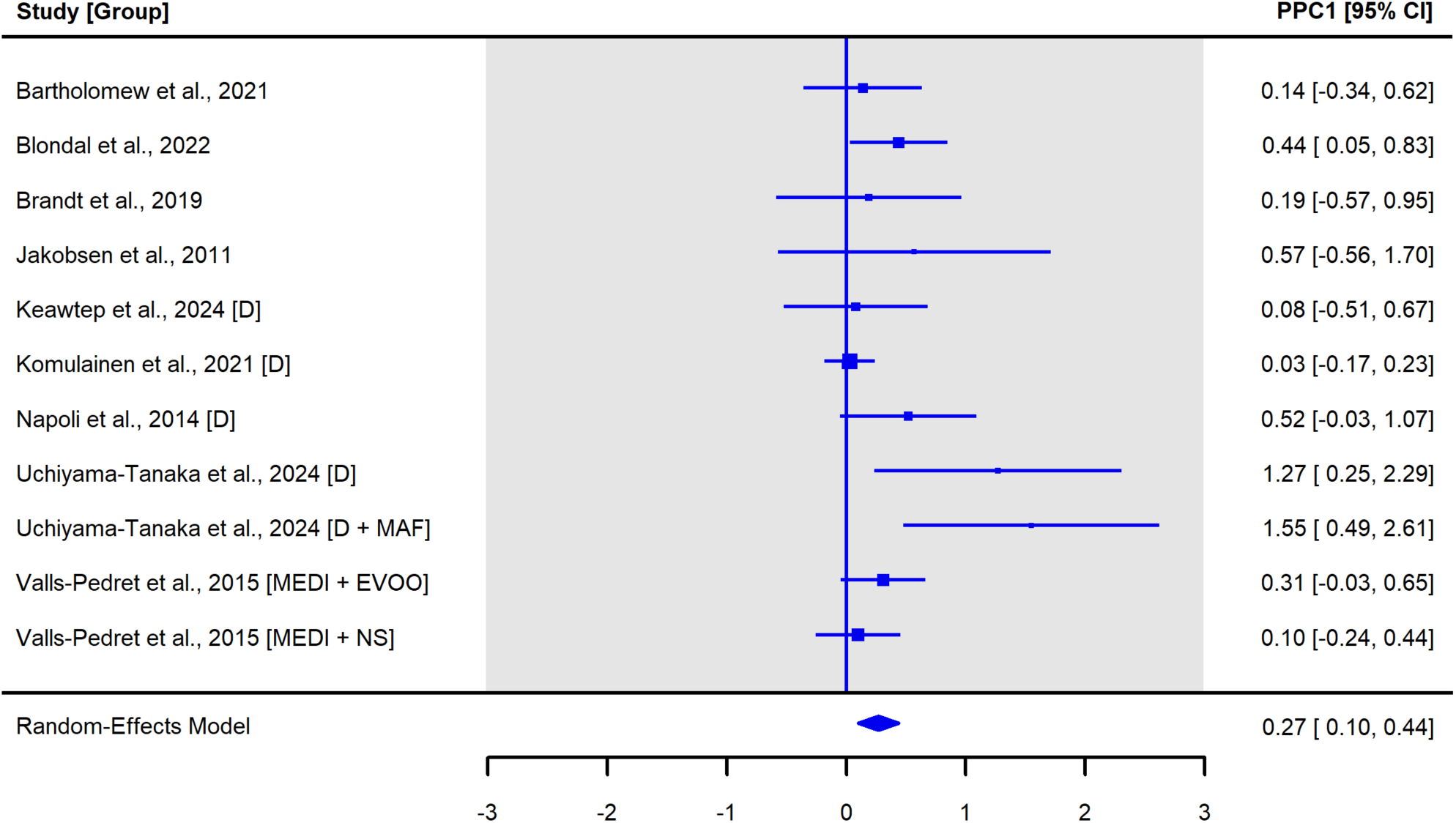
Meta-Analysis Forest Plot of the Pooled Effects of Total Dietary Interventions (k = 11) on Global Cognitive Function.

### Meta-Analysis for Single Food Dietary Interventions (*k* = 9)

A total of 9 single food intervention comparisons were included in the meta-analysis (Figure 8). Results showed a non-significant effect on global cognitive function (Hedges’ *g* = 0.27, 95%CI: -0.13- 0.66, *p* = 0.18). Substantial heterogeneity was observed across studies (*I*^2^ = 67.1%; Q(8) = 21.11, *p* = 0.007) suggesting considerable between-study variability. Egger’s regression test for funnel plot asymmetry was not significant (*t* = 0.17, *p* = 0.87) indicating no evidence of publication bias (Supplementary Figure S8).

**Figure 8.**
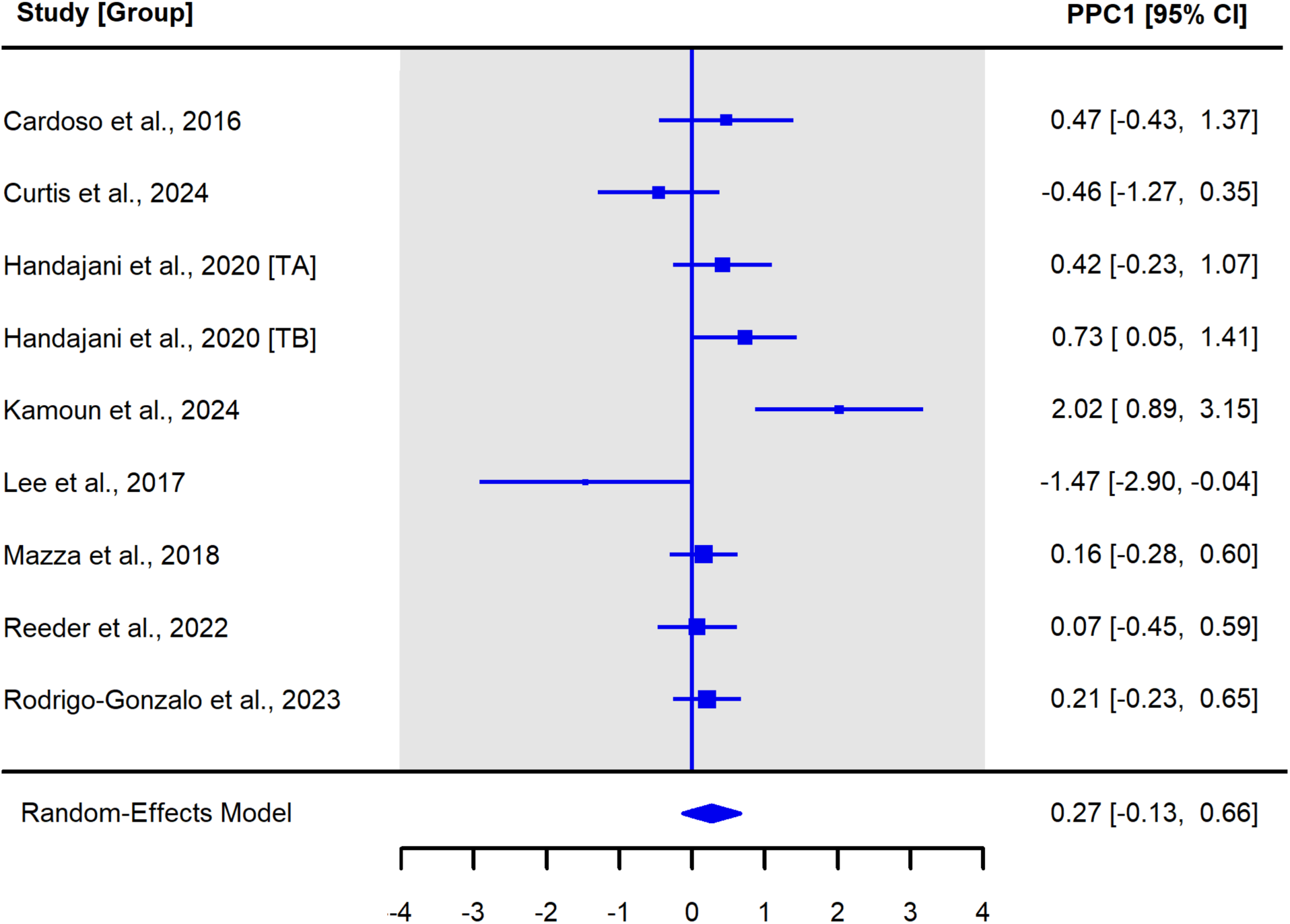
Meta-Analysis Forest Plot of the Pooled Effects of Single Food Interventions (k = 9) on Global Cognitive Function.

### Meta-Regression Analysis

Subgroup analyses revealed distinct patterns of moderator effects based on intervention type (Supplementary Table S8). For multidomain intervention comparisons (k = 15), significant moderation by intervention duration, (QM = 15.36, *p* < .001), comorbidity status (*QM* = 4.24, *p* = .04), and instrument type (*QM* = 5.87, *p* = .02) were found. Shorter intervention durations (≤12 weeks) produced the largest positive effects (β = 0.59, *p* < .001) compared to longer interventions (>52 weeks) which were associated with negative effects (β = -0.52, *p* = .002). Participants with cardiometabolic comorbidities saw larger gains in cognition (β = 0.45, *p* = .04) compared to participants without comorbidities (β = 0.18, *p* = .009). Studies using MMSE detected more conservative effects (β = 0.11, *p* = .01) than other global cognitive measures (β = 0.28, *p* = .02).

Total diet interventions (k = 11) were significantly moderated by risk of bias (*QM* = 7.41, *p* = .007), diet type (*QM* = 10.88, *p* = .05) and intervention duration (*QM* = 6.35, *p* .04). Low/moderate risk of bias studies had substantially larger effects (β = 0.51, *p* < .001) compared to high risk of bias studies that indicated negative effects (β = -0.41, *p* < .001). Only one diet category emerged as statistically significant, and the comparisons were underpowered. A study-generated diet^137^ to reduce advanced glycation end products (AGE) showed a statistically significant effect (β = 1.25, *p* = .002). For intervention duration, no single duration category emerged as statistically significant (*p* > .05), but the overall moderation suggests variability in effects across timeframes with durations of 24-52- weeks trending with the highest effects. No other moderators were statistically significant (*p* > .05).

Single food interventions (k = 9), had no moderators with a statistically significant *QM*, indicating none of the tested factors explained variability in effect sizes for this intervention type.

### Assessment of Minimal Clinically Important Difference (MCID)

Overall, 21 intervention comparisons assessed cognition using MMSE and findings revealed between-group mean difference in MMSE score did not reach MCID (Δ*M* = 1.01). However, when examining cognitively impaired samples only (*k* = 6)^103,112,115,134,159,173^, between-group difference in MMSE score (Δ*M* = 1.82) did reach MCID of 1.4 points, with 2 of these comparisons achieving MCID >2 MMSE points^93,115^ and 2 reaching MCID >3 points^103,115,173^. Seven intervention comparisons^110,122,123,135,156^ assessed cognition using the MoCA and between-group mean change score did not reach MCID (Δ*M* = 1.12). Cognitive status or intervention type had no influence. Only one intervention comparison with participants diagnosed with MCI^133^ assessed cognition using ADAS-Cog and the change score (Δ*M* = -3.71) did reach the MCID of -2.6 points.

### GRADE Certainty of Evidence

#### Multidomain interventions

The certainty of evidence for multidomain interventions on global cognitive function was rated very low. Sensitivity analysis showed no effect size differences by risk of bias, but substantial heterogeneity (*I*^2^ = 67.5%) indicated variability across studies. Variation in components, dosage, and follow-up contributed to indirectness. The overall CI did not cross zero, but spanned values above and below the MCID, with most comparisons (11/15) crossing zero, indicating imprecision. Funnel plot asymmetry and Egger’s test suggested publication bias. Certainty was downgraded for inconsistency, indirectness, imprecision, and suspected publication bias (See Supplementary File Table S9a).

#### Total diet interventions

For total diet interventions, certainty was also very low. All studies had high or some risk of bias. Low heterogeneity (*I*^2^ = 30.6%) suggested limited variability, but differences in intervention type, dosage, comparators, and outcomes contributed to indirectness. The overall CI did not cross zero, but spanned values above and below the MCID. Most studies (6/11) had CIs that crossed zero, indicating imprecision. Funnel plot asymmetry and a significant Egger’s test suggested publication bias. Certainty was downgraded for risk of bias, indirectness, imprecision, and publication bias (See Supplementary File Table S9b).

#### Single food interventions

For single food interventions, certainty was rated very low. Most studies (55.6%) and comparisons (7/9) had high risk of bias. Substantial heterogeneity (*I*^2^ = 67.1%) indicated variability in effects across studies. The overall CI crossed zero and most comparisons (7/9) included CIs reflecting both harm and benefit. Small sample sizes further increased imprecision. No publication bias was found. Certainty was downgraded two levels each for risk of bias and imprecision, and one level each for inconsistency and indirectness (See Supplementary File Table S9c).

## Discussion

This systematic review and meta-analysis synthesised data from 83 distinct trials, 24 063 participants, and 110 intervention comparisons. Meta-analytic results from 35 dietary intervention comparisons demonstrated a small, statistically significant positive effect on global cognitive function (*g* = 0.25, 95%CI: 0.15 to 0.36). Multidomain interventions that combined diet with other lifestyle modifications consistently produced the most robust cognitive gains. Total diet interventions showed similar effects, while single food interventions generally produced null or inconsistent results, likely reflecting the challenges of standardising diet intake and maintenance of an adequate control.

The overall effect size is consistent with previous meta-analyses of dietary interventions reporting small to moderate cognitive benefits^174,175^, particularly in MCI^175^. A prior review of 28 multidomain studies found a significant effect on global cognition and single domains (executive function, memory, and verbal fluency) compared to single interventions, although only two studies included a nutrition component of supplements^67^. Another review of 17 multidomain studies (88.2% included a nutrition component) showed small effects on dementia risk (6 RCTs) and cognitive composite scores (7 RCTs), but in contrast to our findings, found no significant improvement in global cognition (9 RCTs), possibly due to a smaller pooled sample size or a focus on healthy, at-risk or frail older adults, without cognitive impairment^64^.

We found that total diet interventions modestly improve global cognition, whereas single food interventions show no benefit, extending prior research. No prior meta-analysis of RCTs has examined the overall effects of *any* dietary pattern or single food. One meta-analysis examined any dietary pattern as a single intervention (e.g., Mediterranean, low carbohydrate diet, calorie restriction) or as part of a multidomain intervention and found a small effect on global cognitive function (10 RCTs, SMD=0.14, 95% CI 0.01-0.27, p=0.03) in healthy adults^69^, although the independent contribution of dietary patterns versus multidomain studies to the overall effect was unclear. Two meta-analyses of single food interventions, one investigating nut consumption (5 RCTs)^40^ and the other (poly)phenol supplementation (23 RCTs)^27^, both found a non-significant effect on cognitive function (global and single domain) and executive function, respectively, consistent with our null findings for *any* single food. These converging null findings highlight a gap between scientific evidence and public perception. Single food items are frequently marketed as ‘superfoods’ for dementia prevention, but this narrative may be overstating benefits and misdirect public health efforts. Redirecting the focus towards evidence-based strategies, such as total dietary pattern and multidomain lifestyle approaches, may offer greater promise for dementia risk reduction than single food consumption.

New evidence published after our search cut-off further supports our findings. The US POINTER Trial^176^, a 2-year large, multi-site RCT with 2,111 older adults at-risk for cognitive decline, showed that a structured, multidomain lifestyle intervention (MIND diet, exercise, cognitive and social engagement, and cardiovascular monitoring) delivered by trained navigators and peer supporters produced greater gains (*z-*score change = 0.243) in global cognition compared with a lower-intensity, self-guided program (*z-*score change = 0.213). Benefits were strongest for adults with lower baseline cognition, consistent with our findings. Structured support aligns with prior evidence that health coaching enhances participant engagement, self-efficacy, and intervention adherence in chronic disease management^177^. In accordance with our protocol and PRISMA guidance, the POINTER Trial was not included in our quantitative synthesis, but the magnitude and direction of the effects reinforce the patterns observed in our review and strengthen our overall conclusion that multidomain interventions, with structured support, are a promising approach for dementia prevention.

Meta-regression identified risk of bias, intervention duration, diet type, and comorbidity status as moderators. Shorter intervention durations showed the largest effect size improvements in cognition for multidomain interventions, possibly reflecting ceiling effects, or higher adherence and intervention intensity over a shorter period. For total diet interventions, up to 1 year appeared the most effective, likely to allow sufficient time to adopt and maintain dietary behaviour changes. The optimal diet protocol for cognitive benefit remains inconclusive from our review due to a small number of comparisons. However, our overall findings suggest that diets targeting neurodegeneration mechanisms of inflammation, oxidative stress, and impaired glucose metabolism may be the most beneficial. This generally includes a balanced, culturally-relevant, diet with sufficient complete protein intake^178,179^ (e.g., contain all nine essential amino acids) from a variety of animal-based and plant-based sources, use of oils and fats with high polyunsaturated or monounsaturated fatty acids^180,181^, increased consumption of above ground fruits (e.g., berries) and vegetables (e.g., green leafy vegetables) high in flavonoid content^182,183^, and avoiding excessive consumption of ultra-processed foods^184^, saturated or trans-fat sources^185^, and carbohydrate-dense sugary sweets and snacks^186^. Participants with MCI demonstrated greater benefits compared to cognitively normal samples, supporting the potential of diet modification when there is <u>measurable</u> <u>cognitive vulnerability, but before substantial neurodegeneration has occurred</u>^187^. Overall, intensive, time-limited dietary-based interventions in at-risk or MCI groups appear to be the most opportune to potentially reverse early cognitive decline or prevent rapid progression. However, the macronutrient composition of protein, fats, and carbohydrate intake remains unclear and requires further investigation to be able to identify the strongest evidence for diet protocols for cognitive benefit.

### Clinical Relevance

While the pooled meta-analytic effect on global cognitive function was small (*g* = 0.25), interpretation within a clinical context requires comparison with established MCIDs. Across 21 intervention comparisons using the MMSE, the overall between-group mean difference did not reach MCID threshold of 1.4 points. However, in participants with MCI (k = 6), the between-group mean difference increased to 1.82 points, with two comparisons achieving >2 point gains and two comparisons achieving >3 point gains in MMSE. Gains of 2-3 MMSE points may translate into meaningful cognitive and functional preservation and delayed progression to dementia^188^. For the MoCA, seven intervention comparisons yielded a pooled mean change below MCID thresholds. Only one intervention comparison used ADAS-Cog in participants with MCI, making generalisability difficult. MCID results suggest that dietary interventions hold the greatest potential in populations with measurable cognitive impairment, but not yet severe disease progression.

Importantly, in populations with cognitive impairment, *maintenance* of cognitive function or a slowed rate of cognitive decline, rather than improvement, is also clinically meaningful. For example, a population with mild cognitive impairment or Alzheimer’s disease who show no significant change in cognitive function in an intervention group over a long duration (e.g., 52 weeks) have potentially benefited from the intervention, despite null findings. These outcomes are not typically accounted for in current study designs or evidence statements, which emphasise improvement in cognitive test scores. Ultimately, the significance, direction and magnitude of effect between-groups must be considered in the context of dementia as a progressive, neurodegenerative disease.

### Cognitive Measurement Taxonomy

A notable contribution of this review is the development of a novel taxonomy that maps 190 cognitive outcome measures to the six DSM-V neurocognitive domains. This represents an important methodological advancement for a long-standing known problem in the field regarding the lack of standardisation and excessive study variation in how cognitive domains are defined, measured, and reported^189^. The DSM-V classification provides a consensus-based framework^75^, but has seldom been applied in dietary intervention research for cognitive function. Battista et al. (2017)^190^ applied machine learning techniques to 131 neuropsychological outcomes from the Alzheimer’s Disease Neuroimaging Initiative (ADNI) dataset to identify optimal subsets of neuropsychological outcomes for classifying stages of cognitive impairment with 85% (MCI vs. no impairment) to 90% (severe impairment vs. no impairment) accuracies^190^. The authors found ADAS-Cog, Wechsler Logical Memory Scale, Rey Auditory Verbal Learning Test, and Functional Activities Questionnaire (FAQ) were top predictors to differentiate between stages of cognitive impairment^190^. However, their classification framework was not designed for intervention research and did not use a universally recognised conceptual model. Our approach offers a preliminary framework that builds on the DSM-V and combines theory and empirical evidence specific to an intervention context.

While in its early stages, this mapping taxonomy can be used by researchers conducting future trials to reduce construct misclassification, improve comparability of findings across trials, and enhance the power and reliability of future meta-analyses. Prior to implementation, we suggest international consensus on a cognitive measurement taxonomy as a next step.

### Mechanistic Evidence from Brain Imaging

The inclusion of neuroimaging outcomes provides early preliminary mechanistic insights beyond cognitive assessments. Of the 59 reported brain imaging results, just over half (54.2%) favoured the intervention, with significant effects far more pronounced in participants with MCI (>90% significant findings) compared to cognitively normal individuals (29% significant findings). Improvements were most frequently observed in hippocampal volume, gyri, cerebral blood flow, cortical thickness, parietal cortex, and thalamus. These are known brain regions and processes associated with memory formation and consolidation, mood regulation and cognitive resilience. Our review suggests that neurocognitive changes may occur before they are detectable on cognitive assessment tests. This indicates that dietary interventions could have neuroprotective effects even before individuals notice signs of cognitive decline. Integrating imaging biomarkers into trials could help detect early effects and inform refinements to prevention strategies and guidelines. However, financial constraints and barriers to scalability are potential issues. Thus, ultra-sensitive blood assays testing plasma AD biomarkers (e.g., beta-amyloid, p-tau, Neurofilament light [NfL]) may present a cost-effective alternative to implement in future trials^191^. Our findings support the inclusion of *both* objective biomarkers alongside cognitive assessment tools to strengthen the mechanistic evidence, particularly in early disease detection (e.g., preclinical, MCI).

### Methodological Strengths and Limitations

Several methodological considerations and limitations need to be highlighted. First, a key limitation is that only 35 (31.8%) of the 110 intervention comparisons were eligible for inclusion in meta- analyses calculations due to inconsistent reporting, absence of key statistical data, or missing post- intervention results. This substantially reduced the statistical power of pooled estimates and limits the generalisability of the findings. Second, dietary intervention protocols were highly variable in content, dosage, and duration (3-weeks to 8.5 years), which complicates interpretation and comparability across studies. Third, blinding of participants and study personnel is a known challenge in dietary trials raising the risk of performance and detection bias. Fourth, we observed considerable heterogeneity of cognitive outcome measures across four levels: 1) global cognitive function derived from various tests (e.g., MMSE, ADAS-Cog); 2) global cognition derived from author-generated composite scores of varying cognitive domains, (e.g., z-scores of executive function, memory, and processing speed); 3) individual cognitive domains (e.g., tests of executive function) and differing tests that comprised the domain; and 4) inconsistent classification and overlap of cognitive domains themselves, such as ‘Attention, Information & Processing’ versus ‘Processing Speed.’ Fifth, selective reporting and missing outcomes, despite being prespecified in methods, were common, particularly for cognitive outcomes, which may have introduced bias and reduced the reliability of the evidence. Lastly, there was an underrepresentation of studies (13.3%) conducted in LMICs, despite these regions having disproportionately higher dementia cases, which further limits generalisability to diverse, global populations.

Despite methodological challenges, this review represents the largest and most comprehensive known synthesis of dietary intervention trials for cognitive function across the disease continuum of dementia. We offer an initial framework to guide cognitive measurement standardisation in future trials. We integrated psychometric measures of cognition alongside neurobiological outcomes, including brain imaging and blood biomarkers, to generate a broad perspective on intervention efficacy beyond traditional cognitive endpoints and provide potential mechanistic insights. The methodological rigour of this systematic review was ensured through a sensitive search strategy across multiple databases and trial registries, risk of bias assessment at the intervention comparison level, triple coding of data extraction process for accuracy, and sensitivity analysis to confirm the robustness of the findings. The evidence synthesised provides a detailed narrative synthesis with meta-analyses and pre-specified meta-regression to determine sources of heterogeneity.

### Gaps and Future Directions

This review highlights the limited exploration of the combined dietary and non-dietary intervention components that may be associated with the greatest cognitive benefits. This may differ depending on the disease progression stage or presence of co-morbidities. Future trials should test different combinations of components and dosages of dietary protocols to establish consensus on effective intervention components for broader implementation. Long-term follow-up is also recommended to assess sustained effects over time, and replication using standardised frameworks and outcome measures would address the inconsistencies found in our evidence base and improve generalisability. Future trials should also prioritise transparent, consistent reporting following established guidelines (e.g., CONSORT Statement) including presentation of all prespecified outcomes to strengthen reliability in the evidence.

### Call to Action for Research Reform

#### Member of the Public Call to Action

> *“It meant a lot to be involved in this study. With 190 cognitive measures mapped out, this study exposes a critical issue: the field is fragmented, making it difficult to translate the scientific results into actionable steps. When measures vary across studies, we can’t replicate results, compare outcomes, or build the knowledge base needed to guide practice. This review identifies mechanisms at play such as, neuroinflammation, oxidative stress, and dysregulated glucose metabolism, all key drivers of neurodegeneration. Unsurprisingly, these are hallmarks of metabolic dysfunction, the same mechanisms at the core of other conditions such as serious mental illness and type 2 diabetes. Improving metabolic health must become a priority. Researchers and clinicians must unite across disciplines to develop trials targeting these shared mechanistic pathways. For the best outcomes, interventions should include diet, exercise, sleep and structured support, learning from other fields that are addressing metabolic health. Until we do this, the evidence base will remain a maze of disconnected findings and stall progress to prevent dementia. This is a wake-up call and a starting point for reform.”*

> *– ELB, Metabolic Therapies Clinician*

#### Lived-Experience PPI Member Call to Action

> *“The writing retreat for this paper was wonderful. As someone living this daily, what I need the most is clarity. Headlines about diet and dementia are conflicting, leaving me unsure what to believe or how to help my loved one. We need researchers to be consistent and speak with one, unified voice, so when I sit down with my parents, I can look them in the eye and say, “this is worth doing and here is why.” This will help my family make decisions together and keep their independence for as long as possible. Trustworthy and consistent guidance gives my loved one the choice and dignity they deserve, while also releasing constant caregiver worries and responsibilities. This manuscript clearly shows that focusing on total dietary patterns and supporting other lifestyle factors, like sleep and exercise, are more beneficial than single food “superfoods.” This helps! Please focus on studies that confirm what we now know works, using agreed standards so families like mine can make sense of what to do. This will help me support my loved one with confidence, and hope.”*

> *– JB, Daughter of Parent diagnosed with MCI*

## Conclusions

Dietary interventions modestly improve global cognitive function. Multidomain and total diet approaches appear most promising in early cognitive decline. Mapping 190 cognitive assessment tools to DSM-V domains provides a preliminary taxonomy to guide measurement standardisation. Future trials should prioritise consistent reporting of intervention frameworks and cognitive outcomes to strengthen the evidence base for dementia prevention and care.

### Registration and Protocol

The systematic review and meta-analysis protocol, including the methodological details and prespecified outcomes was prospectively registered in the NIHR PROSPERO database (registration: CRD42023488336) and available online: https://www.crd.york.ac.uk/PROSPERO/view/CRD42023488336

### Protocol Amendments

All protocol amendments were documented on the PROSPERO registration record for transparency. A major amendment was the exclusion of trials examining supplements (e.g., vitamins, herbs, minerals) due to wide variation.

## Data Availability

All data generated or analysed in this study are included in this published article and supplementary files. The protocol registration is publicly available on PROSPERO. Additional data, analytic code, and supplementary materials are available from the corresponding author upon reasonable request.

## Acknowledgements

We would like to extend our thanks to Tanya McKay from the McPin Foundation and the 12- member PPI lived-experience panel that informs the NIHR Oxford Health BRC Preventing Multiple Morbidities Theme for their insights and suggestions in developing this project. We would also like to thank Nia Roberts, Senior Outreach Librarian, for their time and contribution supporting the search strategy, Dr. Elena Tsompanaki, for their contribution to screening eligible studies, and Ms. Afrida Asad and Jiani Chen, for their time and contribution to data extraction.

## Copyright

The corresponding author has the right to grant on behalf of all authors and does grant on behalf of all authors, an exclusive licence on a worldwide basis to the BMJ Publishing Group Ltd. to permit this article to be published in the BMJ editions and any other BMJPGL products and sublicences such use and exploit all subsidiary rights, as set out in our licence.

## Transparency Declaration

The lead author and manuscript’s guarantor, MK, affirms that the manuscript is an honest, accurate, and transparent account of the study being reported; that no important aspects of the study have been omitted; and that any discrepancies from the study as planned have been explained and documented in full.

## Ethical Approval

Not required.

## CREDIT Authorship Contribution

**MK:** Corresponding author; Conceptualisation, Methodology, Validation, Formal Analysis, Investigation, Resources, Data Curation, Writing – Original Draft, Writing – Review & Editing, Visualization, Supervision, Project Administration, Funding Acquisition

**OC:** Conceptualisation, Methodology, Formal Analysis, Investigation, Resources, Data Curation, Writing – Original Draft, Writing – Review & Editing, Visualization, Supervision

**MP:** Methodology, Software, Validation, Formal Analysis, Investigation, Data Curation, Writing – Original Draft, Writing – Review and Editing

**FC:** Validation, Software, Formal Analysis, Investigation, Data Curation, Visualization, Writing – Review and Editing

**MS:** Formal Analysis, Investigation, Visualisation, Writing – Review and Editing

**RR:** Formal Analysis, Investigation, Visualisation, Writing – Review and Editing

**AH:** Formal Analysis, Investigation, Visualisation, Writing – Review and Editing

**JB:** PPIE Lived Experience Perspective, Investigation, Writing – Original Draft, Writing – Review and Editing

**BC:** Investigation, Writing – Review and Editing **HK:** Investigation, Writing – Review and Editing

**EL:** Investigation, Writing – Review and Editing

**JG:** PPIE Lived Experience Perspective, Investigation, Writing – Original Draft, Writing – Review and Editing

**EB:** Writing – Review and Editing, PPIE Clinician Perspective

**JK:** Writing – Review and Editing

**IK:** Writing – Review & Editing

**KB:** Supervision, Writing – Review & Editing

## References

1. World Health Organization. Dementia. Key facts. March 15, 2023. https://www.who.int/news-room/fact-sheets/detail/dementia#:~:text=Key%20facts,injuries%20that%20affect%20the%20brain.

2. Nichols E, Steinmetz JD, Vollset SE, et al. Estimation of the global prevalence of dementia in 2019 and forecasted prevalence in 2050: an analysis for the Global Burden of Disease Study 2019. Lancet Public Health. 2022;7(2):e105–e125.

3. Alzheimer’s Research UK. Statistics about Dementia. Dementia Statistics Hub. 2022. https://dementiastatistics.org/about-dementia/

4. Alzheimer’s Disease International. Dementia Statistics. Dementia Statistics. n.d. https://www.alzint.org/about/dementia-facts-figures/dementia-statistics/

5. Kobayashi LC, Ehrlich JR. Closing the Data Gaps on Trends in Dementia and Related Care in Low-and Middle-Income Countries. J Gerontol A Biol Sci Med Sci. 2024;79(Supplement_1):S5–S6.

6. Office for National Statistics. Death registration summary statistics, England and Wales: 2022. Office for National Statistics. April 11, 2023. https://www.ons.gov.uk/peoplepopulationandcommunity/birthsdeathsandmarriages/deaths/articles/deathregistrationsummarystatisticsenglandandwales/2022#:~:text=The%20leading%20cause%20of%20death,10.4%25%20of%20all%20deaths).

7. Office for National Statistics. Leading causes of death, UK: 2001 to 2018. Office for National Statistics. March 27, 2020. https://www.ons.gov.uk/releases/leadingcausesofdeathuk

8. Nandi A, Counts N, Chen S, et al. Global and regional projections of the economic burden of Alzheimer’s disease and related dementias from 2019 to 2050: A value of statistical life approach. EClinicalMedicine. 2022;51.

9. NHS England. Dementia. Dementia. 2024. https://www.england.nhs.uk/mental-health/dementia/#:~:text=There%20is%20a%20considerable%20economic,cancer%2C%20heart%20disease%20and%20stroke.

10. Moreta MPG, Burgos-Alonso N, Torrecilla M, Marco-Contelles J, Bruzos-Cidón C. Efficacy of acetylcholinesterase inhibitors on cognitive function in Alzheimer’s disease. Review of reviews. Biomedicines. 2021;9(11):1689.

11. Dou KX, Tan MS, Tan CC, et al. Comparative safety and effectiveness of cholinesterase inhibitors and memantine for Alzheimer’s disease: a network meta-analysis of 41 randomized controlled trials. Alzheimers Res Ther. 2018;10:1–10.

12. Penninkilampi R, Brothers HM, Eslick GD. Safety and efficacy of anti-amyloid-β immunotherapy in Alzheimer’s disease: A systematic review and meta-analysis. J Neuroimmune Pharmacol. 2017;12:194–203.

13. Livingston G, Huntley J, Liu KY, et al. Dementia prevention, intervention, and care: 2024 report of the Lancet standing Commission. The Lancet. 2024;404(10452):572–628.

14. Stephen R, Barbera M, Peters R, et al. Development of the first WHO guidelines for risk reduction of cognitive decline and dementia: lessons learned and future directions. Front Neurol. 2021;12:763573.

15. Markun S, Gravestock I, Jäger L, Rosemann T, Pichierri G, Burgstaller JM. Effects of Vitamin B12 Supplementation on Cognitive Function, Depressive Symptoms, and Fatigue: A Systematic Review, Meta-Analysis, and Meta-Regression. Nutrients. 2021;13(3):923. doi:10.3390/nu13030923

16. Gil Martínez V, Avedillo Salas A, Santander Ballestín S. Vitamin Supplementation and Dementia: A Systematic Review. Nutrients. 2022;14(5):1033. doi:10.3390/nu14051033

17. Wang Z, Zhu W, Xing Y, Jia J, Tang Y. B vitamins and prevention of cognitive decline and incident dementia: a systematic review and meta-analysis. Nutr Rev. 2022;80(4):931–949. doi:10.1093/nutrit/nuab057

18. Forbes SC, Holroyd-Leduc JM, Poulin MJ, Hogan DB. Effect of Nutrients, Dietary Supplements and Vitamins on Cognition: a Systematic Review and Meta-Analysis of Randomized Controlled Trials. Can Geriatr J. 2015;18(4):231–245. doi:10.5770/cgj.18.189

19. Farina N, Llewellyn D, Isaac MGEKN, Tabet N. Vitamin E for Alzheimer’s dementia and mild cognitive impairment. Cochrane Database Syst Rev. 2017;1(1):CD002854. doi:10.1002/14651858.CD002854.pub4

20. Chen F, Wang J, Cheng Y, et al. Magnesium and Cognitive Health in Adults: A Systematic Review and Meta-Analysis. Adv Nutr Bethesda Md. 2024;15(8):100272. doi:10.1016/j.advnut.2024.100272

21. Suh SW, Lim E, Burm SY, et al. The influence of n-3 polyunsaturated fatty acids on cognitive function in individuals without dementia: a systematic review and dose-response meta-analysis. BMC Med. 2024;22(1):109. doi:10.1186/s12916-024-03296-0

22. Wei BZ, Li L, Dong CW, Tan CC, Alzheimer’s Disease Neuroimaging Initiative, Xu W. The Relationship of Omega-3 Fatty Acids with Dementia and Cognitive Decline: Evidence from Prospective Cohort Studies of Supplementation, Dietary Intake, and Blood Markers. Am J Clin Nutr. 2023;117(6):1096–1109. doi:10.1016/j.ajcnut.2023.04.001

23. Zhu RZ, Chen MQ, Zhang ZW, Wu TY, Zhao WH. Dietary fatty acids and risk for Alzheimer’s disease, dementia, and mild cognitive impairment: A prospective cohort meta-analysis. Nutr Burbank Los Angel Cty Calif. 2021;90:111355. doi:10.1016/j.nut.2021.111355

24. Clifford T, Babateen A, Shannon OM, et al. Effects of inorganic nitrate and nitrite consumption on cognitive function and cerebral blood flow: A systematic review and meta-analysis of randomized clinical trials. Crit Rev Food Sci Nutr. 2019;59(15):2400–2410. doi:10.1080/10408398.2018.1453779

25. Ammar A, Trabelsi K, Boukhris O, et al. Effects of Polyphenol-Rich Interventions on Cognition and Brain Health in Healthy Young and Middle-Aged Adults: Systematic Review and Meta-Analysis. J Clin Med. 2020;9(5):1598. doi:10.3390/jcm9051598

26. Colizzi C. The protective effects of polyphenols on Alzheimer’s disease: A systematic review. Alzheimers Dement N Y N. 2019;5:184–196. doi:10.1016/j.trci.2018.09.002

27. Farag S, Tsang C, Murphy PN. Polyphenol supplementation and executive functioning in overweight and obese adults at risk of cognitive impairment: A systematic review and meta-analysis. PloS One. 2023;18(5):e0286143. doi:10.1371/journal.pone.0286143

28. Godos J, Micek A, Mena P, et al. Dietary (Poly)phenols and Cognitive Decline: A Systematic Review and Meta-Analysis of Observational Studies. Mol Nutr Food Res. 2024;68(1):e2300472. doi:10.1002/mnfr.202300472

29. Coelho-Júnior HJ, Calvani R, Landi F, Picca A, Marzetti E. Protein Intake and Cognitive Function in Older Adults: A Systematic Review and Meta-Analysis. Nutr Metab Insights. 2021;14:11786388211022373. doi:10.1177/11786388211022373

30. Yang M, Cai C, Yang Z, et al. Effect of dietary fibre on cognitive function and mental health in children and adolescents: a systematic review and meta-analysis. Food Funct. 2024;15(17):8618–8628. doi:10.1039/D4FO02221A

31. Kirvalidze M, Hodkinson A, Storman D, et al. The role of glucose in cognition, risk of dementia, and related biomarkers in individuals without type 2 diabetes mellitus or the metabolic syndrome: A systematic review of observational studies. Neurosci Biobehav Rev. 2022;135:104551. doi:10.1016/j.neubiorev.2022.104551

32. Liu H, Liu Y, Shi M, Zhou Y, Zhao Y, Xia Y. Meta-analysis of sugar-sweetened beverage intake and the risk of cognitive disorders. J Affect Disord. 2022;313:177–185. doi:10.1016/j.jad.2022.06.048

33. Ruan Y, Tang J, Guo X, Li K, Li D. Dietary Fat Intake and Risk of Alzheimer’s Disease and Dementia: A Meta-Analysis of Cohort Studies. Curr Alzheimer Res. 15(9):869–876. doi:10.2174/1567205015666180427142350

34. Cao GY, Li M, Han L, et al. Dietary Fat Intake and Cognitive Function among Older Populations: A Systematic Review and Meta-Analysis. J Prev Alzheimers Dis. 2019;6(3):204–211. doi:10.14283/jpad.2019.9

35. Godos J, Micek A, Currenti W, et al. Fish consumption, cognitive impairment and dementia: an updated dose-response meta-analysis of observational studies. Aging Clin Exp Res. 2024;36(1):171. doi:10.1007/s40520-024-02823-6

36. Kosti RI, Kasdagli MI, Kyrozis A, et al. Fish intake, n-3 fatty acid body status, and risk of cognitive decline: a systematic review and a dose-response meta-analysis of observational and experimental studies. Nutr Rev. 2022;80(6):1445–1458. doi:10.1093/nutrit/nuab078

37. Fazlollahi A, Motlagh Asghari K, Aslan C, et al. The effects of olive oil consumption on cognitive performance: a systematic review. Front Nutr. 2023;10:1218538. doi:10.3389/fnut.2023.1218538

38. Alkhalifa AE, Al-Ghraiybah NF, Kaddoumi A. Extra-Virgin Olive Oil in Alzheimer’s Disease: A Comprehensive Review of Cellular, Animal, and Clinical Studies. Int J Mol Sci. 2024;25(3):1914. doi:10.3390/ijms25031914

39. Hein S, Whyte AR, Wood E, Rodriguez-Mateos A, Williams CM. Systematic Review of the Effects of Blueberry on Cognitive Performance as We Age. J Gerontol A Biol Sci Med Sci. 2019;74(7):984–995. doi:10.1093/gerona/glz082

40. Moabedi M, Aliakbari M, Erfanian S, Jibril AT, Milajerdi A. The effect of consuming nuts on cognitive function: a systematic review and meta-analysis of randomized clinical trials. Front Nutr. 2024;11:1463801. doi:10.3389/fnut.2024.1463801

41. Aridi YS, Walker JL, Wright ORL. The Association between the Mediterranean Dietary Pattern and Cognitive Health: A Systematic Review. Nutrients. 2017;9(7):674. doi:10.3390/nu9070674

42. Devranis P, Vassilopoulou Ε, Tsironis V, et al. Mediterranean Diet, Ketogenic Diet or MIND Diet for Aging Populations with Cognitive Decline: A Systematic Review. Life. 2023;13(1):173. doi:10.3390/life13010173

43. Fekete M, Varga P, Ungvari Z, et al. The role of the Mediterranean diet in reducing the risk of cognitive impairement, dementia, and Alzheimer’s disease: a meta-analysis. GeroScience. 2025;47(3):3111–3130. doi:10.1007/s11357-024-01488-3

44. Fu J, Tan LJ, Lee JE, Shin S. Association between the mediterranean diet and cognitive health among healthy adults: A systematic review and meta-analysis. Front Nutr. 2022;9. doi:10.3389/fnut.2022.946361

45. Gregory S, Pullen H, Ritchie CW, Shannon OM, Stevenson EJ, Muniz-Terrera G. Mediterranean diet and structural neuroimaging biomarkers of Alzheimer’s and cerebrovascular disease: A systematic review. Exp Gerontol. 2023;172:112065. doi:10.1016/j.exger.2022.112065

46. Loughrey DG, Lavecchia S, Brennan S, Lawlor BA, Kelly ME. The Impact of the Mediterranean Diet on the Cognitive Functioning of Healthy Older Adults: A Systematic Review and Meta-Analysis. Adv Nutr Bethesda Md. 2017;8(4):571–586. doi:10.3945/an.117.015495

47. Nucci D, Sommariva A, Degoni LM, et al. Association between Mediterranean diet and dementia and Alzheimer disease: a systematic review with meta-analysis. Aging Clin Exp Res. 2024;36(1):77. doi:10.1007/s40520-024-02718-6

48. Petersson SD, Philippou E. Mediterranean Diet, Cognitive Function, and Dementia: A Systematic Review of the Evidence. Adv Nutr. 2016;7(5):889–904. doi:10.3945/an.116.012138

49. Singh B, Parsaik AK, Mielke MM, et al. Association of mediterranean diet with mild cognitive impairment and Alzheimer’s disease: a systematic review and meta-analysis. J Alzheimers Dis JAD. 2014;39(2):271–282. doi:10.3233/JAD-130830

50. Chen X, Maguire B, Brodaty H, O’Leary F. Dietary Patterns and Cognitive Health in Older Adults: A Systematic Review. J Alzheimers Dis JAD. 2019;67(2):583–619. doi:10.3233/JAD-180468

51. Solfrizzi V, Custodero C, Lozupone M, et al. Relationships of Dietary Patterns, Foods, and Micro- and Macronutrients with Alzheimer’s Disease and Late-Life Cognitive Disorders: A Systematic Review. J Alzheimers Dis JAD. 2017;59(3):815–849. doi:10.3233/JAD-170248

52. Chen H, Dhana K, Huang Y, et al. Association of the Mediterranean Dietary Approaches to Stop Hypertension Intervention for Neurodegenerative Delay (MIND) Diet With the Risk of Dementia. JAMA Psychiatry. 2023;80(6):630–638. doi:10.1001/jamapsychiatry.2023.0800

53. van Soest AP, Beers S, van de Rest O, de Groot LC. The Mediterranean-Dietary Approaches to Stop Hypertension Intervention for Neurodegenerative Delay (MIND) Diet for the Aging Brain: A Systematic Review. Adv Nutr. 2024;15(3):100184. doi:10.1016/j.advnut.2024.100184

54. Kheirouri S, Alizadeh M. MIND diet and cognitive performance in older adults: a systematic review. Crit Rev Food Sci Nutr. 2022;62(29):8059–8077. doi:10.1080/10408398.2021.1925220

55. Chinna-Meyyappan A, Gomes FA, Koning E, Fabe J, Breda V, Brietzke E. Effects of the ketogenic diet on cognition: a systematic review. Nutr Neurosci. 2023;26(12):1258–1278. doi:10.1080/1028415X.2022.2143609

56. Rong L, Peng Y, Shen Q, Chen K, Fang B, Li W. Effects of ketogenic diet on cognitive function of patients with Alzheimer’s disease: a systematic review and meta-analysis. J Nutr Health Aging. 2024;28(8):100306. doi:10.1016/j.jnha.2024.100306

57. Christodoulou CC, Pitsillides M, Hadjisavvas A, Zamba-Papanicolaou E. Dietary Intake, Mediterranean and Nordic Diet Adherence in Alzheimer’s Disease and Dementia: A Systematic Review. Nutrients. 2025;17(2):336. doi:10.3390/nu17020336

58. Veronese N, Facchini S, Stubbs B, et al. Weight loss is associated with improvements in cognitive function among overweight and obese people: A systematic review and meta-analysis. Neurosci Biobehav Rev. 2017;72:87–94. doi:10.1016/j.neubiorev.2016.11.017

59. Lü W, Yu T, Kuang W. Effects of dietary restriction on cognitive function: a systematic review and meta-analysis. Nutr Neurosci. 2023;26(6):540–550. doi:10.1080/1028415X.2022.2068876

60. Siervo M, Arnold R, Wells JCK, et al. Intentional weight loss in overweight and obese individuals and cognitive function: a systematic review and meta-analysis. Obes Rev. 2011;12(11):968–983. doi:10.1111/j.1467-789X.2011.00903.x

61. Smith M, Watson P, Gallacher J, Bauermeister S. Ultra-processed food exposure and cognitive outcomes: A systematic review of observational studies. medRxiv. Preprint posted online February 13, 2025:2025.02.12.25322127. doi:10.1101/2025.02.12.25322127

62. Nguyen L, Walters J, Spies B, Coppus A, Massie J, Bertonha TP. Food for Thought: A Systematic Review and Meta-Analysis on the Effects of Ultra-Processed Foods on Cognition in Children and Adolescents. Food Front. 2025;6(4):1838–1866. doi:10.1002/fft2.70064

63. Kivipelto M, Mangialasche F, Ngandu T. Lifestyle interventions to prevent cognitive impairment, dementia and Alzheimer disease. Nat Rev Neurol. 2018;14(11):653–666. doi:10.1038/s41582-018-0070-3

64. Meng X, Fang S, Zhang S, et al. Multidomain lifestyle interventions for cognition and the risk of dementia: A systematic review and meta-analysis. Int J Nurs Stud. 2022;130:104236. doi:10.1016/j.ijnurstu.2022.104236

65. Noach S, Witteman B, Boss HM, Janse A. Effects of multidomain lifestyle interventions on cognitive decline and Alzheimer’s disease prevention: A literature review and future recommendations. Cereb Circ - Cogn Behav. 2023;4:100166. doi:10.1016/j.cccb.2023.100166

66. Reparaz-Escudero I, Izquierdo M, Bischoff-Ferrari HA, Martínez-Lage P, Sáez de Asteasu ML. Effect of long-term physical exercise and multidomain interventions on cognitive function and the risk of mild cognitive impairment and dementia in older adults: A systematic review with meta-analysis. Ageing Res Rev. 2024;100:102463. doi:10.1016/j.arr.2024.102463

67. Salzman T, Sarquis-Adamson Y, Son S, Montero-Odasso M, Fraser S. Associations of Multidomain Interventions With Improvements in Cognition in Mild Cognitive Impairment: A Systematic Review and Meta-analysis. JAMA Netw Open. 2022;5(5):e226744. doi:10.1001/jamanetworkopen.2022.6744

68. Zou C, Amos-Richards D, Jagannathan R, Kulshreshtha A. Effect of home-based lifestyle interventions on cognition in older adults with mild cognitive impairment: a systematic review. BMC Geriatr. 2024;24(1):200. doi:10.1186/s12877-024-04798-5

69. McEvoy CT, Leng Y, Peeters GM, Kaup AR, Allen IE, Yaffe K. Interventions involving a major dietary component improve cognitive function in cognitively healthy adults: a systematic review and meta-analysis. Nutr Res N Y N. 2019;66:1–12. doi:10.1016/j.nutres.2019.02.008

70. García-Casares N, Gallego Fuentes P, Barbancho MÁ, López-Gigosos R, García-Rodríguez A, Gutiérrez-Bedmar M. Alzheimer’s Disease, Mild Cognitive Impairment and Mediterranean Diet. A Systematic Review and Dose-Response Meta-Analysis. J Clin Med. 2021;10(20):4642. doi:10.3390/jcm10204642

71. He Q, Bennett AN, Zhang C, Zhang JY, Tong S, Chan KHK. Nutritional interventions for preventing cognitive decline in patients with mild cognitive impairment and Alzheimer’s disease: A comprehensive network meta-analysis and Mendelian Randomization study. Clin Nutr ESPEN. 2025;67:555–566. doi:10.1016/j.clnesp.2025.03.040

72. Anderson EL, Davies NM, Korologou-Linden R, Kivimäki M. Dementia prevention: the Mendelian randomisation perspective. J Neurol Neurosurg Psychiatry. 2024;95(4):384–390. doi:10.1136/jnnp-2023-332293

73. Page MJ, McKenzie JE, Bossuyt PM, et al. The PRISMA 2020 statement: an updated guideline for reporting systematic reviews. bmj. 2021;372.

74. Povey R, Conner M, Sparks P, James R, Shepherd R. A critical examination of the application of the Transtheoretical Model’s stages of change to dietary behaviours. Health Educ Res. 1999;14(5):641–651.

75. Sachdev PS, Blacker D, Blazer DG, et al. Classifying neurocognitive disorders: the DSM-5 approach. Nat Rev Neurol. 2014;10(11):634–642.

76. Covidence. Covidence Systematic Review Software. 2024. Accessed January 24, 2025. https://www.covidence.org

77. Jisc. Jisc Online Surveys. Accessed June 2, 2024. https://www.jisc.ac.uk/online-surveys

78. Schulz KF, Altman DG, Moher D, Group C.* CONSORT 2010 statement: updated guidelines for reporting parallel group randomized trials. Ann Intern Med. 2010;152(11):726–732.

79. Juszczak E, Altman DG, Hopewell S, Schulz K. Reporting of multi-arm parallel-group randomized trials: extension of the CONSORT 2010 statement. Jama. 2019;321(16):1610–1620.

80. Hoffmann TC, Glasziou PP, Boutron I, et al. Better reporting of interventions: template for intervention description and replication (TIDieR) checklist and guide. Bmj. 2014;348.

81. Sterne JA, Savović J, Page MJ, et al. RoB 2: a revised tool for assessing risk of bias in randomised trials. bmj. 2019;366.

82. Posit team. RStudio: Integrated Development Environment for R. Posit Software, PBC; 2025. http://www.posit.co/

83. Viechtbauer W. Conducting meta-analyses in R with the metafor package. J Stat Softw. 2010;36(3):1–48. doi:10.18637/jss.v036.i03

84. Morris SB. Estimating effect sizes from pretest-posttest-control group designs. Organ Res Methods. 2008;11(2):364–386.

85. Skvarc DR, Fuller-Tyszkiewicz M. Calculating repeated-measures meta-analytic effects for continuous outcomes: a tutorial on pretest–posttest-controlled designs. Adv Methods Pract Psychol Sci. 2024;7(1):25152459231217238.

86. Higgins J. Cochrane handbook for systematic reviews of interventions. *Cochrane Collab John Wiley Sons Ltd*. Published online 2008.

87. Cohen J. Statistical Power Analysis for the Behavioral Sciences. routledge; 2013.

88. Watt JA, Veroniki AA, Tricco AC, Straus SE. Using a distribution-based approach and systematic review methods to derive minimum clinically important differences. BMC Med Res Methodol. 2021;21(1):41.

89. Wu CY, Hung SJ, Lin K chung, Chen KH, Chen P, Tsay PK. Responsiveness, minimal clinically important difference, and validity of the MoCA in stroke rehabilitation. Occup Ther Int. 2019;2019(1):2517658.

90. Andrieu S, Guyonnet S, Coley N, et al. Effect of long-term omega 3 polyunsaturated fatty acid supplementation with or without multidomain intervention on cognitive function in elderly adults with memory complaints (MAPT): a randomised, placebo-controlled trial. Lancet Neurol. 2017;16(5):377–389. doi:10.1016/s1474-4422(17)30040-6

91. Arjmand G, Abbas-Zadeh M, Eftekhari MH. Effect of MIND diet intervention on cognitive performance and brain structure in healthy obese women: a randomized controlled trial. Sci Rep. 2022;12(1). doi:10.1038/s41598-021-04258-9

92. Babateen AM, Shannon OM, O’Brien GM, et al. Incremental doses of nitrate-rich beetroot juice do not modify cognitive function and cerebral blood flow in overweight and obese older adults: a 13-week pilot randomised clinical trial. Nutrients. 2022;14(5):1052.

93. Barnes LL, Dhana K, Liu X, et al. Trial of the MIND Diet for Prevention of Cognitive Decline in Older Persons. N Engl J Med. 2023;389(7):602–611. doi:10.1056/nejmoa2302368

94. Bartholomew CL, Muhlestein JB, May HT, et al. Randomized controlled trial of once-per-week intermittent fasting for health improvement: the WONDERFUL trial. Bäck M, ed. Eur Heart J Open. 2021;1(2). doi:10.1093/ehjopen/oeab026

95. Blondal BS, Geirsdottir OG, Halldorsson TI, Beck AM, Jonsson PV, Ramel A. HOMEFOOD randomised trial – Six-month nutrition therapy improves quality of life, self-rated health, cognitive function, and depression in older adults after hospital discharge. Clin Nutr ESPEN. 2022;48:74–81. doi:10.1016/j.clnesp.2022.01.010

96. Blumenthal JA, Smith PJ, Mabe S, et al. Longer Term Effects of Diet and Exercise on Neurocognition: 1-Year Follow-up of the ENLIGHTEN Trial. J Am Geriatr Soc. 2020;68(3):559–568. doi:10.1111/jgs.16252

97. Boespflug EL, Eliassen JC, Dudley JA, et al. Enhanced neural activation with blueberry supplementation in mild cognitive impairment. Nutr Neurosci. 2018;21(4):297–305.

98. Bøhn SK, Myhrstad MC, Thoresen M, et al. Bilberry/red grape juice decreases plasma biomarkers of inflammation and tissue damage in aged men with subjective memory impairment–a randomized clinical trial. BMC Nutr. 2021;7:1–17.

99. Bookheimer SY, Renner BA, Ekstrom A, et al. Pomegranate Juice Augments Memory and fMRI Activity in Middle-Aged and Older Adults with Mild Memory Complaints. Evid Based Complement Alternat Med. 2013;2013(1):946298.

100. Bowtell JL, Aboo-Bakkar Z, Conway ME, Adlam ALR, Fulford J. Enhanced task-related brain activation and resting perfusion in healthy older adults after chronic blueberry supplementation. Appl Physiol Nutr Metab. 2017;42(7):773–779.

101. Brinkworth GD. Long-term Effects of a Very Low-Carbohydrate Diet and a Low-Fat Diet on Mood and Cognitive Function. Arch Intern Med. 2009;169(20):1873. doi:10.1001/archinternmed.2009.329

102. Brodaty H, Chau T, Heffernan M, et al. An online multidomain lifestyle intervention to prevent cognitive decline in at-risk older adults: a randomized controlled trial. Nat Med. 2025;31(2):565–573. doi:10.1038/s41591-024-03351-6

103. Buchholz A, Deme P, Betz JF, Brandt J, Haughey N, Cervenka MC. A randomized feasibility trial of the modified Atkins diet in older adults with mild cognitive impairment due to Alzheimer’s disease. Front Endocrinol. 2024;15. doi:10.3389/fendo.2024.1182519

104. Cardoso BR, Apolinário D, da Silva Bandeira V, et al. Effects of Brazil nut consumption on selenium status and cognitive performance in older adults with mild cognitive impairment: a randomized controlled pilot trial. Eur J Nutr. 2016;55:107–116.

105. Chai SC, Jerusik J, Davis K, Wright RS, Zhang Z. Effect of Montmorency tart cherry juice on cognitive performance in older adults: a randomized controlled trial. Food Funct. 2019;10(7):4423–4431.

106. Chan SC, Esther GE, Yip HL, Sugathan S, Chin PS. Effect of cold pressed coconut oil on cognition and behavior among patients with Alzheimer s disease-A pilot intervention study. Natl J Physiol Pharm Pharmacol. 2017;7(12):1432–1432.

107. Chatterjee P, Kumar DA, Naqushbandi S, et al. Effect of Multimodal Intervention (computer based cognitive training, diet and exercise) in comparison to health awareness among older adults with Subjective Cognitive Impairment (MISCI-Trial)—A Pilot Randomized Control Trial. Abdelbasset WK, ed. PLOS ONE. 2022;17(11):e0276986. doi:10.1371/journal.pone.0276986

108. Cheatham CL, Canipe III LG, Millsap G, et al. Six-month intervention with wild blueberries improved speed of processing in mild cognitive decline: a double-blind, placebo-controlled, randomized clinical trial. Nutr Neurosci. 2023;26(10):1019–1033.

109. Chlebowski RT, Rapp S, Aragaki AK, et al. Low-fat dietary pattern and global cognitive function: Exploratory analyses of the Women’s Health Initiative (WHI) randomized Dietary Modification trial. eClinicalMedicine. 2020;18:100240. doi:10.1016/j.eclinm.2019.100240

110. Chou CC, Li YJ, Wang CJ, Lyu LC. A mini-flipped, game-based Mediterranean diet learning program on dietary behavior and cognitive function among community-dwelling older adults in Taiwan: A cluster-randomized controlled trial. Geriatr Nur (Lond). 2022;45:160–168. doi:10.1016/j.gerinurse.2022.03.009

111. Coates AM, Morgillo S, Yandell C, et al. Effect of a 12-week almond-enriched diet on biomarkers of cognitive performance, mood, and cardiometabolic health in older overweight adults. Nutrients. 2020;12(4):1180.

112. Curtis AF, Musich M, Costa AN, et al. Feasibility and preliminary efficacy of American elderberry juice for improving cognition and inflammation in patients with mild cognitive impairment. Int J Mol Sci. 2024;25(8):4352.

113. Halyburton AK, Brinkworth GD, Wilson CJ, et al. Low- and high-carbohydrate weight-loss diets have similar effects on mood but not cognitive performance. Am J Clin Nutr. 2007;86(3):580–587. doi:10.1093/ajcn/86.3.580

114. Han CY, Sharma Y, Yaxley A, Baldwin C, Woodman R, Miller M. Individualized Hospital to Home, Exercise-Nutrition Self-Managed Intervention for Pre-Frail and Frail Hospitalized Older Adults: The INDEPENDENCE Randomized Controlled Pilot Trial. Clin Interv Aging. 2023;Volume 18:809–825. doi:10.2147/cia.s405144

115. Handajani YS, Turana Y, Yogiara Y, et al. Tempeh consumption and cognitive improvement in mild cognitive impairment. Dement Geriatr Cogn Disord. 2020;49(5):497–502.

116. Hardman RJ, Meyer D, Kennedy G, Macpherson H, Scholey AB, Pipingas A. Findings of a Pilot Study Investigating the Effects of Mediterranean Diet and Aerobic Exercise on Cognition in Cognitively Healthy Older People Living Independently within Aged-Care Facilities: The Lifestyle Intervention in Independent Living Aged Care (LIILAC) Study. Curr Dev Nutr. 2020;4(5):nzaa077. doi:10.1093/cdn/nzaa077

117. Horie NC, Serrao VT, Simon SS, et al. Cognitive Effects of Intentional Weight Loss in Elderly Obese Individuals With Mild Cognitive Impairment. J Clin Endocrinol Metab. 2016;101(3):1104–1112. doi:10.1210/jc.2015-2315

118. Hoscheidt S, Sanderlin AH, Baker LD, et al. Mediterranean and Western diet effects on Alzheimer’s disease biomarkers, cerebral perfusion, and cognition in mid-life: A randomized trial. Alzheimers Dement. 2022;18(3):457–468. doi:10.1002/alz.12421

119. Jakobsen LH, Kondrup J, Zellner M, Tetens I, Roth E. Effect of a high protein meat diet on muscle and cognitive functions: A randomised controlled dietary intervention trial in healthy men. Clin Nutr. 2011;30(3):303–311. doi:10.1016/j.clnu.2010.12.010

120. James DL, Mun CJ, Larkey LK, et al. Health impacts of a remotely delivered prolonged nightly fasting intervention in stressed adults with memory decline and obesity: A nationwide randomized controlled pilot trial. J Clin Transl Sci. 2024;8(1). doi:10.1017/cts.2024.651

121. Jennings A, Shannon OM, Gillings R, et al. Effectiveness and feasibility of a theory-informed intervention to improve Mediterranean diet adherence, physical activity and cognition in older adults at risk of dementia: the MedEx-UK randomised controlled trial. BMC Med. 2024;22(1). doi:10.1186/s12916-024-03815-z

122. Kamoun A, Yahia A, Farjallah MA, et al. Concurrent training associated with moderate walnut consumption improved isokinetic strength, subjective sleep quality, cognitive performance and postural balance in elderly active men: A randomized controlled trial. Aging Clin Exp Res. 2024;36(1):50.

123. Keawtep P, Sungkarat S, Boripuntakul S, et al. Effects of combined dietary intervention and physical-cognitive exercise on cognitive function and cardiometabolic health of postmenopausal women with obesity: a randomized controlled trial. Int J Behav Nutr Phys Act. 2024;21(1). doi:10.1186/s12966-024-01580-z

124. Kimble R, Keane KM, Lodge JK, Cheung W, Haskell-Ramsay CF, Howatson G. Polyphenol-rich tart cherries (Prunus Cerasus, cv Montmorency) improve sustained attention, feelings of alertness and mental fatigue and influence the plasma metabolome in middle-aged adults: a randomised, placebo-controlled trial. Br J Nutr. 2022;128(12):2409–2420.

125. Knight A, Bryan J, Wilson C, Hodgson J, Davis C, Murphy K. The Mediterranean Diet and Cognitive Function among Healthy Older Adults in a 6-Month Randomised Controlled Trial: The MedLey Study. Nutrients. 2016;8(9):579. doi:10.3390/nu8090579

126. Koblinsky ND, Anderson ND, Ajwani F, et al. Feasibility and preliminary efficacy of the LEAD trial: a cluster randomized controlled lifestyle intervention to improve hippocampal volume in older adults at-risk for dementia. Pilot Feasibility Stud. 2022;8(1). doi:10.1186/s40814-022-00977-6

127. Komulainen P, Tuomilehto J, Savonen K, et al. Exercise, diet, and cognition in a 4-year randomized controlled trial: Dose-Responses to Exercise Training (DR’s EXTRA). Am J Clin Nutr. 2021;113(6):1428–1439. doi:10.1093/ajcn/nqab018

128. Krikorian R, Nash TA, Shidler MD, Shukitt-Hale B, Joseph JA. Concord grape juice supplementation improves memory function in older adults with mild cognitive impairment. Br J Nutr. 2010;103(5):730–734.

129. Krikorian R, Shidler MD, Dangelo K, Couch SC, Benoit SC, Clegg DJ. Dietary ketosis enhances memory in mild cognitive impairment. Neurobiol Aging. 2012;33(2):425.e19–425.e27. doi:10.1016/j.neurobiolaging.2010.10.006

130. Krikorian R, Boespflug EL, Fleck DE, et al. Concord grape juice supplementation and neurocognitive function in human aging. J Agric Food Chem. 2012;60(23):5736–5742.

131. Krikorian R, Skelton MR, Summer SS, Shidler MD, Sullivan PG. Blueberry supplementation in midlife for dementia risk reduction. Nutrients. 2022;14(8):1619.

132. Krikorian R, Shidler MD, Summer SS. Early intervention in cognitive aging with strawberry supplementation. Nutrients. 2023;15(20):4431.

133. Lee J, Torosyan N, Silverman DH. Examining the impact of grape consumption on brain metabolism and cognitive function in patients with mild decline in cognition: A double-blinded placebo controlled pilot study. Exp Gerontol. 2017;87:121–128.

134. Lee EH, Kim GH, Park HK, et al. Effects of the multidomain intervention with nutritional supplements on cognition and gut microbiome in early symptomatic Alzheimer’s disease: a randomized controlled trial. Front Aging Neurosci. 2023;15. doi:10.3389/fnagi.2023.1266955

135. Liang CK, Lee WJ, Hwang AC, et al. Efficacy of Multidomain Intervention Against Physio-cognitive Decline Syndrome: A Cluster-randomized Trial. Arch Gerontol Geriatr. 2021;95:104392. doi:10.1016/j.archger.2021.104392

136. Makris A, Darcey VL, Rosenbaum DL, et al. Similar effects on cognitive performance during high- and low-carbohydrate obesity treatment. Nutr Diabetes. 2013;3(9):e89–e89. doi:10.1038/nutd.2013.29

137. Marseglia A, Xu W, Fratiglioni L, et al. Effect of the NU-AGE Diet on Cognitive Functioning in Older Adults: A Randomized Controlled Trial. Front Physiol. 2018;9. doi:10.3389/fphys.2018.00349

138. Martin CK, Anton SD, Han H, et al. Examination of Cognitive Function During Six Months of Calorie Restriction: Results of a Randomized Controlled Trial. Rejuvenation Res. 2007;10(2):179–190. doi:10.1089/rej.2006.0502

139. Martínez-Lapiscina EH, Clavero P, Toledo E, et al. Mediterranean diet improves cognition: the PREDIMED-NAVARRA randomised trial. J Neurol Neurosurg Psychiatry. 2013;84(12):1318–1325. doi:10.1136/jnnp-2012-304792

140. Masley SC, Weaver W, Peri G, Phillips SE. Efficacy of lifestyle changes in modifying practical markers of wellness and aging. Altern Ther Health Med. 2008;14(2):24–31.

141. Mazza E, Fava A, Ferro Y, et al. Effect of the replacement of dietary vegetable oils with a low dose of extravirgin olive oil in the Mediterranean Diet on cognitive functions in the elderly. J Transl Med. 2018;16:1–10.

142. McMaster M, Kim S, Clare L, et al. Lifestyle Risk Factors and Cognitive Outcomes from the Multidomain Dementia Risk Reduction Randomized Controlled Trial, Body Brain Life for Cognitive Decline (BBL-CD). J Am Geriatr Soc. 2020;68(11):2629–2637. doi:10.1111/jgs.16762

143. Mendoza-Ruvalcaba N, Arias-Merino ED. “I am active”: effects of a program to promote active aging. Clin Interv Aging. Published online May 2015:829. doi:10.2147/cia.s79511

144. Miller MG, Hamilton DA, Joseph JA, Shukitt-Hale B. Dietary blueberry improves cognition among older adults in a randomized, double-blind, placebo-controlled trial. Eur J Nutr. 2018;57:1169–1180.

145. Miller MG, Thangthaeng N, Rutledge GA, Scott TM, Shukitt-Hale B. Dietary strawberry improves cognition in a randomised, double-blind, placebo-controlled trial in older adults. Br J Nutr. 2021;126(2):253–263.

146. Mirheidary R, Saber SSE, Shaeiri MR, others. The effect of “mavizˮ on memory improvement in university students: A randomized open-label clinical trial. Avicenna J Phytomedicine. 2020;10(4):352.

147. Moon SY, Hong CH, Jeong JH, et al. Facility-based and home-based multidomain interventions including cognitive training, exercise, diet, vascular risk management, and motivation for older adults: a randomized controlled feasibility trial. Aging. 2021;13(12):15898–15916. doi:10.18632/aging.203213

148. Nakazeko T, Shobako N, Shioya N, et al. Frailty-Preventing Effect of an Intervention Program Using a Novel Complete Nutritional “COMB-FP Meal”: A Pilot Randomized Control Trial. Nutrients. 2023;15(20):4317. doi:10.3390/nu15204317

149. Napoli N, Shah K, Waters DL, Sinacore DR, Qualls C, Villareal DT. Effect of weight loss, exercise, or both on cognition and quality of life in obese older adults. Am J Clin Nutr. 2014;100(1):189–198. doi:10.3945/ajcn.113.082883

150. Ngandu T, Lehtisalo J, Solomon A, et al. A 2 year multidomain intervention of diet, exercise, cognitive training, and vascular risk monitoring versus control to prevent cognitive decline in at-risk elderly people (FINGER): a randomised controlled trial. The Lancet. 2015;385(9984):2255–2263.

151. Ornish D, Madison C, Kivipelto M, et al. Effects of intensive lifestyle changes on the progression of mild cognitive impairment or early dementia due to Alzheimer’s disease: a randomized, controlled clinical trial. Alzheimers Res Ther. 2024;16(1). doi:10.1186/s13195-024-01482-z

152. Parilli-Moser I, Domínguez-López I, Trius-Soler M, et al. Consumption of peanut products improves memory and stress response in healthy adults from the ARISTOTLE study: A 6-month randomized controlled trial. Clin Nutr. 2021;40(11):5556–5567.

153. Rakic JM, Tanprasertsuk J, Scott TM, et al. Effects of daily almond consumption for six months on cognitive measures in healthy middle-aged to older adults: a randomized control trial. Nutr Neurosci. 2022;25(7):1466–1476.

154. Reeder N, Tolar-Peterson T, Adegoye GA, Dickinson E, McFatter E. The effect of daily peanut consumption on cognitive function and indicators of mental health among healthy young women. Funct Foods Health Dis. 2022;12(12):734–747.

155. Rizvi ZA, Saleem J, Zeb I, et al. Effects of intermittent fasting on body composition, clinical health markers and memory status in the adult population: a single-blind randomised controlled trial. Nutr J. 2024;23(1). doi:10.1186/s12937-024-01046-9

156. Roach JC, Rapozo MK, Hara J, et al. A remotely coached multimodal lifestyle intervention for Alzheimer’s disease ameliorates functional and cognitive outcomes. J Alzheimer’s Dis. 2023;96(2):591–607.

157. Rodrigo-Gonzalo MJ, González-Manzano S, Pablos-Hernández MC, et al. Effects of a Raisin Supplement on Cognitive Performance, Quality of Life, and Functional Activities in Healthy Older Adults—Randomized Clinical Trial. Nutrients. 2023;15(12):2811.

158. Rutledge GA, Sandhu AK, Miller MG, Edirisinghe I, Burton-Freeman BB, Shukitt-Hale B. Blueberry phenolics are associated with cognitive enhancement in supplemented healthy older adults. Food Funct. 2021;12(1):107–118.

159. Sakurai T, Sugimoto T, Akatsu H, et al. Japan-Multimodal Intervention Trial for the Prevention of Dementia: A randomized controlled trial. Alzheimers Dement. 2024;20(6):3918–3930. doi:10.1002/alz.13838

160. Sala-Vila A, Valls-Pedret C, Rajaram S, et al. Effect of a 2-year diet intervention with walnuts on cognitive decline. The Walnuts And Healthy Aging (WAHA) study: a randomized controlled trial. Am J Clin Nutr. 2020;111(3):590–600.

161. Siddarth P, Li Z, Miller KJ, et al. Randomized placebo-controlled study of the memory effects of pomegranate juice in middle-aged and older adults. Am J Clin Nutr. 2020;111(1):170–177.

162. Silver RE, Roberts SB, Kramer AF, Chui KKH, Das SK. No Effect of Calorie Restriction or Dietary Patterns on Spatial Working Memory During a 2-Year Intervention: A Secondary Analysis of the CALERIE Trial. J Nutr. 2023;153(3):733–740. doi:10.1016/j.tjnut.2023.01.019

163. Smith PJ, Blumenthal JA, Babyak MA, et al. Effects of the Dietary Approaches to Stop Hypertension Diet, Exercise, and Caloric Restriction on Neurocognition in Overweight Adults With High Blood Pressure. Hypertension. 2010;55(6):1331–1338. doi:10.1161/hypertensionaha.109.146795

164. Smith PJ, Mabe SM, Sherwood A, et al. Metabolic and Neurocognitive Changes Following Lifestyle Modification: Examination of Biomarkers from the ENLIGHTEN Randomized Clinical Trial. J Alzheimers Dis. 2020;77(4):1793–1803. doi:10.3233/jad-200374

165. Thunborg C, Wang R, Rosenberg A, et al. Integrating a multimodal lifestyle intervention with medical food in prodromal Alzheimer’s disease: the MIND-ADmini randomized controlled trial. Alzheimers Res Ther. 2024;16(1). doi:10.1186/s13195-024-01468-x

166. Tussing-Humphreys L, Lamar M, McLeod A, et al. Effect of Mediterranean diet and Mediterranean diet plus calorie restriction on cognition, lifestyle, and cardiometabolic health: A randomized clinical trial. Prev Med Rep. 2022;29:101955. doi:10.1016/j.pmedr.2022.101955

167. Uchiyama-Tanaka Y, Yamakage H, Inui T. The Effects of Dietary Intervention and Macrophage-Activating Factor Supplementation on Cognitive Function in Elderly Users of Outpatient Rehabilitation. Nutrients. 2024;16(13):2078. doi:10.3390/nu16132078

168. Valls-Pedret C, Sala-Vila A, Serra-Mir M, et al. Mediterranean Diet and Age-Related Cognitive Decline: A Randomized Clinical Trial. JAMA Intern Med. 2015;175(7):1094. doi:10.1001/jamainternmed.2015.1668

169. Wardle J, Rogers P, Judd P, et al. Randomized trial of the effects of cholesterol-lowering dietary treatment on psychological function∗∗Access the “Journal Club” discussion of this paper at http://www.elsevier.com/locate/ajmselect/. Am J Med. 2000;108(7):547–553. doi:10.1016/s0002-9343(00)00330-2

170. Wood E, Hein S, Mesnage R, et al. Wild blueberry (poly) phenols can improve vascular function and cognitive performance in healthy older individuals: a double-blind randomized controlled trial. Am J Clin Nutr. 2023;117(6):1306–1319.

171. Zhu L, Ming Y, Wu M, et al. Effect of Fiber-Rich Diet and Rope Skipping on Memory, Executive Function, and Gut Microbiota in Young Adults: A Randomized Controlled Trial. Mol Nutr Food Res. 2024;68(3). doi:10.1002/mnfr.202300673

172. Zülke AE, Pabst A, Luppa M, et al. A multidomain intervention against cognitive decline in an at-risk-population in Germany: Results from the cluster-randomized AgeWell.de trial. Alzheimers Dement. 2024;20(1):615–628. doi:10.1002/alz.13486

173. Brandt J, Buchholz A, Henry-Barron B, Vizthum D, Avramopoulos D, Cervenka MC. Preliminary Report on the Feasibility and Efficacy of the Modified Atkins Diet for Treatment of Mild Cognitive Impairment and Early Alzheimer’s Disease. J Alzheimers Dis. 2019;68(3):969–981. doi:10.3233/jad-180995

174. Radd-Vagenas S, Duffy SL, Naismith SL, Brew BJ, Flood VM, Singh MAF. Effect of the Mediterranean diet on cognition and brain morphology and function: a systematic review of randomized controlled trials. Am J Clin Nutr. 2018;107(3):389–404.

175. Charbit J, Vidal JS, Hanon O. Effects of Dietary Interventions on Cognitive Outcomes. Nutrients. 2025;17(12):1964.

176. Baker LD, Espeland MA, Whitmer RA, et al. Structured vs self-guided multidomain lifestyle interventions for global cognitive function: the US POINTER randomized clinical trial. JAMA. Published online 2025.

177. Pirbaglou M, Katz J, Motamed M, Pludwinski S, Walker K, Ritvo P. Personal health coaching as a type 2 diabetes mellitus self-management strategy: a systematic review and meta-analysis of randomized controlled trials. Am J Health Promot. 2018;32(7):1613–1626.

178. Keum M, Lee BC, Choe YM, et al. Protein intake and episodic memory: the moderating role of the apolipoprotein E ε4 status. Alzheimers Res Ther. 2024;16(1):181.

179. Gao R, Yang Z, Yan W, Du W, Zhou Y, Zhu F. Protein intake from different sources and cognitive decline over 9 years in community-dwelling older adults. Front Public Health. 2022;10:1016016.

180. Morris MC, Tangney CC. Dietary fat composition and dementia risk. Neurobiol Aging. 2014;35:S59–S64.

181. Tessier AJ, Cortese M, Yuan C, et al. Consumption of olive oil and diet quality and risk of dementia-related death. JAMA Netw Open. 2024;7(5):e2410021–e2410021.

182. Jennings A, Thompson AS, Tresserra-Rimbau A, et al. Flavonoid-rich foods, dementia risk, and interactions with genetic risk, hypertension, and depression. JAMA Netw Open. 2024;7(9):e2434136–e2434136.

183. Shishtar E, Rogers GT, Blumberg JB, Au R, Jacques PF. Long-term dietary flavonoid intake and risk of Alzheimer disease and related dementias in the Framingham Offspring Cohort. Am J Clin Nutr. 2020;112(2):343–353.

184. Gonçalves NG, Ferreira NV, Khandpur N, et al. Association between consumption of ultraprocessed foods and cognitive decline. JAMA Neurol. 2023;80(2):142–150.

185. Ruan Y, Tang J, Guo X, Li K, Li D. Dietary fat intake and risk of Alzheimer’s disease and dementia: a meta-analysis of cohort studies. Curr Alzheimer Res. 2018;15(9):869–876.

186. Zhang S, Xiao Y, Cheng Y, et al. Associations of sugar intake, high-sugar dietary pattern, and the risk of dementia: a prospective cohort study of 210,832 participants. BMC Med. 2024;22(1):298.

187. Daviglus ML, Plassman BL, Pirzada A, et al. Risk factors and preventive interventions for Alzheimer disease: state of the science. Arch Neurol. 2011;68(9):1185–1190.

188. Andrews JS, Desai U, Kirson NY, Zichlin ML, Ball DE, Matthews BR. Disease severity and minimal clinically important differences in clinical outcome assessments for Alzheimer’s disease clinical trials. Alzheimers Dement Transl Res Clin Interv. 2019;5(1):354–363.

189. Bahar-Fuchs A, Clare L, Woods B. Cognitive training and cognitive rehabilitation for mild to moderate Alzheimer’s disease and vascular dementia. Cochrane Database Syst Rev. 2013;(6).

190. Battista P, Salvatore C, Castiglioni I. Optimizing neuropsychological assessments for cognitive, behavioral, and functional impairment classification: a machine learning study. Behav Neurol. 2017;2017(1):1850909.

191. Pais MV, Forlenza OV, Diniz BS. Plasma biomarkers of Alzheimer’s disease: a review of available assays, recent developments, and implications for clinical practice. J Alzheimers Dis Rep. 2023;7(1):355–380.

