## Supplementary File for "Dietary Interventions and Cognitive Function across the Dementia Continuum: A Systematic Review, Meta-Analysis, Meta-Regression and Call to Action for Research Reform"

### Supplementary Material

### Table of Contents

***Supplementary Table S1.*** *PsychINFO search strategy* ... 2

***Supplementary Table S2.*** *Secondary reports of included studies (n = 9)* ... 5

***Supplementary Table S3.*** *Descriptive characteristics of included studies (n = 83)* ... 7

***Supplementary Table S4a.*** *Multidomain intervention study characteristics (n = 30)* ... 12

***Supplementary Table S4b.*** *Total diet intervention study characteristics (n = 33)* ... 27

***Supplementary Table S4c.*** *Single food intervention study characteristics (n = 31)* ... 34

***Supplementary Table S5.*** *Taxonomy of extracted cognitive outcome measures mapped to the six DSM-V neurocognitive domains* ... 39

***Supplementary Table S6a.*** *Between-group differences in cognitive outcomes at post-intervention in multi-domain interventions (n = 30)* ... 48

***Supplementary Table S6b.*** *Between-group differences in cognitive outcomes as post-intervention in total diet interventions (n = 33)* ... 53

***Supplementary Table S6c.*** *Between-group differences in cognitive outcomes at post-intervention in single food interventions (n = 31)* ... 59

***Supplementary Table S7.*** *Hedges' g pre-post change sensitivity analysis across correlation coefficients 0.10-0.90* ... 67

***Supplementary Table S8.*** *Meta-regression analysis of moderators of dietary intervention effects on global cognitive function* ... 68

***Supplementary Table S9a.*** *GRADE assessment for certainty of evidence for multidomain intervention arms (k = 15)* ... 71

***Supplementary Table S9b.*** *GRADE assessment for certainty of evidence for total diet intervention arms (k = 11)* ... 72

***Supplementary Table S9c.*** *GRADE assessment for certainty of evidence for single food intervention arms (k = 15)* ... 73

***Supplementary Figure S1.*** *Risk of bias assessment for multidomain intervention arms* ... 74

***Supplementary Figure S2.*** *Risk of bias assessment for total diet intervention arms* ... 75

***Supplementary Figure S3.*** *Risk of bias assessment for single food intervention arms* ... 76

***Supplementary Figure S4.*** *Risk of bias assessment for cluster randomised controlled trials (rcts)* ... 77

***Supplementary Figure S5.*** *Funnel plot of effect sizes versus standard errors for all studies included in the main meta-analysis* ... 78

***Supplementary Figure S6.*** *Funnel plot of effect sizes versus standard errors for all multidomain studies included in the meta-analysis* ... 78

***Supplementary Figure S7.*** *Funnel plot of effect sizes versus standard errors for all total diet intervention studies included in the meta-analysis* ... 79

***Supplementary Figure S8.*** *Funnel plot of effect sizes versus standard errors for all single-food intervention studies included in the meta-analysis* ... 79

**Supplementary Table S1.** PsychINFO search strategy

| | | |
|---|---|---|
| | PsycINFO 1806 to present | |
| 1 | dementia/ or alzheimer's disease/ or vascular dementia/ | 89409 |
| 2 | cognitive impairment/ or memory disorders/ or mild cognitive impairment/ | 52444 |
| 3 | Cognition/ or Memory/ | 110766 |
| 4 | mini mental state examination/ | 1028 |
| 5 | (dementia? or alzheimer* or cognition or memory or (cognitive adj2 (function or dysfunction or decline or disorder? or impair*))).ti,id. | 297223 |
| 6 | (mmse or mini mental state exam* or mmms or modified mini mental score).ti,ab,id. | 12416 |
| 7 | 1 or 2 or 3 or 4 or 5 or 6 | 336389 |
| 8 | exp diets/ or exp dietary supplements/ or healthy eating/ | 34433 |
| 9 | dietary treatment/ | 272 |
| 10 | food/ or fast food/ | 15634 |
| 11 | diet*.ti,id. | 20710 |
| 12 | ((diet* or nutrition* or food or health* eating or health* lifestyle) adj3 (intervention? or program*)).ti,ab,id. | 7532 |
| 13 | ((diet* or nutrition* or mineral* or vitamin*) adj2 supplement*).ti,ab,id. | 4322 |
| 14 | ((low or high) adj2 (fat? or carbohydrate? or protein or sodium) adj5 (diet? or intervention? or program*)).ti,ab,id. | 3021 |
| 15 | ((ketogenic or keto or paleo or mediterranean or nordic or vegan or vegetarian or gluten free) adj2 diet?).ti,ab,id. | 1642 |
| 16 | ((dash or mind or atkins) adj2 diet?).ti,ab,id. | 181 |
| 17 | (mindful eating or intuitive eating or intermittent fasting).ti,ab,id. | 592 |
| 18 | 8 or 9 or 10 or 11 or 12 or 13 or 14 or 15 or 16 or 17 | 61773 |
| 19 | 7 and 18 | 4303 |
| 20 | random*.ti,ab,hw,id. | 247592 |
| 21 | trial*.ti,ab,hw,id. | 221554 |
| 22 | controlled stud*.ti,ab,hw,id. | 14080 |
| 23 | placebo*.ti,ab,hw,id. | 44987 |
| 24 | ((singl* or doubl* or trebl* or tripl*) and (blind* or mask*)).ti,ab,hw,id. | 33065 |
| 25 | (cross over or crossover or factorial* or latin square).ti,ab,hw,id. | 35966 |
| 26 | (assign* or allocat* or volunteer*).ti,ab,hw,id. | 194087 |
| 27 | treatment effectiveness evaluation/ | 29058 |
| 28 | mental health program evaluation/ | 2449 |
| 29 | exp experimental design/ | 63936 |
| 30 | (clinical trial or treatment outcome).md. | 61793 |

| | | |
|---|---|---|
| 31 | 20 or 21 or 22 or 23 or 24 or 25 or 26 or 27 or 28 or 29 or 30 | 627362 |
| 32 | (animal not human).po. | 389732 |
| 33 | 31 not 32 | 589891 |
| 34 | 19 and 33 | 843 |
| 35 | limit 34 to English language | 816 |

Update search - 2025

<u>PsycINFO 1806 to present</u>

| | | |
|---|---|---|
| 1 | dementia/ or Alzheimer's disease/ or vascular dementia/ | 95422 |
| 2 | cognitive impairment/ or memory disorders/ or mild cognitive impairment/ | 56355 |
| 3 | Cognition/ or Memory/ | 114489 |
| 4 | mini mental state examination/ | 1132 |
| 5 | (dementia? or Alzheimer* or cognition or memory or (cognitive adj2 (function or dysfunction or decline or disorder? or impair*))).ti,id. | 311560 |
| 6 | (mmse or mini mental state exam* or mmms or modified mini mental score).ti,ab,id. | 13058 |
| 7 | 1 or 2 or 3 or 4 or 5 or 6 | 352428 |
| 8 | exp diets/ or exp dietary supplements/ or healthy eating/ | 36129 |
| 9 | dietary treatment/ | 313 |
| 10 | food/ or fast food/ | 16153 |
| 11 | diet*.ti,id. | 21705 |
| 12 | ((diet* or nutrition* or food or health* eating or health* lifestyle) adj3 (intervention? or program*)).ti,ab,id. | 8028 |
| 13 | ((diet* or nutrition* or mineral* or vitamin*) adj2 supplement*).ti,ab,id. | 4566 |
| 14 | ((low or high) adj2 (fat? or carbohydrate? or protein or sodium) adj5 (diet? or intervention? or program*)).ti,ab,id. | 3143 |
| 15 | ((ketogenic or keto or paleo or mediterranean or nordic or vegan or vegetarian or gluten free) adj2 diet?).ti,ab,id. | 1764 |
| 16 | ((dash or mind or atkins) adj2 diet?).ti,ab,id. | 207 |
| 17 | (mindful eating or intuitive eating or intermittent fasting).ti,ab,id. | 672 |
| 18 | 8 or 9 or 10 or 11 or 12 or 13 or 14 or 15 or 16 or 17 | 64638 |
| 19 | 7 and 18 | 4632 |
| 20 | random*.ti,ab,hw,id. | 262851 |
| 21 | trial*.ti,ab,hw,id. | 233665 |
| 22 | controlled stud*.ti,ab,hw,id. | 14651 |
| 23 | placebo*.ti,ab,hw,id. | 46202 |
| 24 | ((singl* or doubl* or trebl* or tripl*) and (blind* or mask*)).ti,ab,hw,id. | 34283 |

| | | |
|---|---|---|
| 25 | (cross over or crossover or factorial* or latin square).ti,ab,hw,id. | 37628 |
| 26 | (assign* or allocat* or volunteer*).ti,ab,hw,id. | 203000 |
| 27 | treatment effectiveness evaluation/ | 30846 |
| 28 | mental health program evaluation/ | 2547 |
| 29 | exp experimental design/ | 65054 |
| 30 | (clinical trial or treatment outcome).md. | 67521 |
| 31 | 20 or 21 or 22 or 23 or 24 or 25 or 26 or 27 or 28 or 29 or 30 | 657568 |
| 32 | (animal not human).po. | 393281 |
| 33 | 31 not 32 | 620956 |
| 34 | 19 and 33 | 904 |
| 35 | limit 34 to english language | 880 |
| 36 | (2024* or 2025*).up,yr. | 240527 |
| 37 | 35 and 36 | 66 |

**Supplementary Table S2.** Secondary reports of included studies (n = 9)

| First Author (Year)[Ref] | Country | Sample size (T0/T1) | Age *M(SD)* | % Female | % Non-White | Trial Design | RCT Type | Research Setting | Cognitive Health Status | Other Co-Morbidities | INT Type | Global Cognitive Outcome Measure | Other Cognitive Measures | Brain or Blood Markers |
|---|---|---|---|---|---|---|---|---|---|---|---|---|---|---|
| Blumenthal (2020)[1] | USA | 160/149 | 65.4 (6.8) | 47.0 | 46.9 | Factorial, 4-arm | Single-site | CRF | MCI | CVD risk | MD, TD | NTB, CDR | 10 | -- |
| Brandt (2019)[2] | USA | 27/14 | 71.9 (6.1) | 44.4 | 7.1 | Parallel, 2-arm | Single-site | ARI | MCI, early AD | -- | TD | MMSE-2 | 4 | BB |
| Chhetri (2018)[3] | France | 1680/1293 | 75.4 (4.3) | 63.4 | -- | Parallel, 4-arm | Multi-site | Clinic | At-Risk | -- | MD | MMSE | 12 | -- |
| Grigolon (2020)[4] | USA | 220/185 | 38.1 (7.2) | 69.6 | 22.7 | Parallel, 2-arm | Multi-site | Clinic | Normal | -- | TD | -- | 1 | -- |
| Komulainen (2010)[5] | Finland | 1410/1292 | 66.3 (5.4) | -- | -- | Parallel, 6-arm | Individual | Community | Normal | -- | MD, TD | CERAD, MMSE | 7 | -- |
| Leclerc (2020)[6] | USA | 220/185 | 38.1 (7.2) | 69.6 | 22.7 | Parallel, 2-arm | Multi-site | Clinic | Normal | -- | TD | -- | 1 | -- |
| Martinez-Lapiscina (2013b)[7] | Spain | 271/268 | 74.1 (5.7) | 55.2 | -- | Parallel, 3-arm | Single-site | CRF | Normal, MCI | T2DM, Vascular Risk | TD | MMSE | 12 | -- |
| Rosenberg (2018)[8] | Finland | 1260/1105 | 69.3 (4.7) | 46.3 | -- | Parallel, 2-arm | Multi-site | Clinic | At-Risk | -- | MD | NTB, MMSE | 14 | BB |
| Zellner (2011)[9] | Denmark | 23/23 | 24.2 (3.5) | 0.0 | -- | Parallel, 2-arm | Single-site | ARI | Normal | -- | TD | -- | 1 | -- |

Citations:

1. Blumenthal JA, Smith PJ, Mabe S, et al. Longer Term Effects of Diet and Exercise on Neurocognition: 1-Year Follow-up of the ENLIGHTEN Trial. *J American Geriatrics Society*. 2020;68(3):559-568. doi:10.1111/jgs.16252
2. Brandt J, Buchholz A, Henry-Barron B, Vizthum D, Avramopoulos D, Cervenka MC. Preliminary Report on the Feasibility and Efficacy of the Modified Atkins Diet for Treatment of Mild Cognitive Impairment and Early Alzheimer's Disease. *JAD*. 2019;68(3):969-981. doi:10.3233/jad-180995

3. Chhetri JK, De Souto Barreto P, Cantet C, et al. Effects of a 3-Year Multi-Domain Intervention with or without Omega-3 Supplementation on Cognitive Functions in Older Subjects with Increased CAIDE Dementia Scores. *Journal of Alzheimer's Disease*. 2018;64(1):71-78. doi:10.3233/jad-180209
4. Grigolon RB, Brietzke E, Trevizol AP, McIntyre RS, Mansur RB. Caloric restriction, resting metabolic rate and cognitive performance in Non-obese adults: A post-hoc analysis from CALERIE study. *Journal of Psychiatric Research*. 2020;128:16-22. doi:10.1016/j.jpsychires.2020.05.018
5. Komulainen P, Kivipelto M, Lakka TA, et al. Exercise, fitness and cognition – A randomised controlled trial in older individuals: The DR's EXTRA study. *European Geriatric Medicine*. 2010;1(5):266-272. doi:10.1016/j.eurger.2010.08.001
6. Leclerc E, Trevizol AP, Grigolon RB, et al. The effect of caloric restriction on working memory in healthy non-obese adults. *CNS Spectr*. 2020;25(1):2-8. doi:10.1017/s1092852918001566
7. Martinez-Lapiscina EH, Clavero P, Toledo E, et al. Virgin olive oil supplementation and long-term cognition: the Predimed-Navarra randomized, trial. *The Journal of nutrition, health and aging*. 2013;17(6):544-552. doi:10.1007/s12603-013-0027-6
8. Rosenberg A, Ngandu T, Rusanen M, et al. Multidomain lifestyle intervention benefits a large elderly population at risk for cognitive decline and dementia regardless of baseline characteristics: The FINGER trial. *Alzheimer's & Dementia*. 2018;14(3):263-270. doi:10.1016/j.jalz.2017.09.006
9. Zellner M, Babeluk R, Jakobsen LH, et al. A proteomics study reveals a predominant change in MaoB expression in platelets of healthy volunteers after high protein meat diet: relationship to the methylation cycle. *J Neural Transm*. 2011;118(5):653-662. doi:10.1007/s00702-011-0617-6

**Supplementary Table S3.** Descriptive characteristics of included studies (*n* = 83)

| First Author (Year)[Ref] | Country | Sample size (T0/T1) | Age *M(SD)* | % Female | % Non-White | Trial Design | RCT Type | Research Setting | Cognitive Health Status | Other Co-Morbid Conditions | INT Type | Global Cognitive Outcome Measure | Single-Domain Cognitive Measures | Brain Imaging or Blood Markers |
|---|---|---|---|---|---|---|---|---|---|---|---|---|---|---|
| Andrieu (2017) | France | 1680/1286 | 75.3 (4.4) | 64.0 | -- | Parallel, 4-arm | Multi-site | Clinic | At-Risk | -- | MD | MMSE, Composite | 12 | PET, BB |
| Arjmand (2022) | Iran | 40/37 | 48.0 (5.3) | 100 | -- | Parallel, 2-arm | Single-Site | Clinic | Normal | Obese | TD | -- | 8 | MRI, BB |
| Babateen (2022) | UK | 62/50 | 66.3 (3.7) | 61.3 | -- | Parallel, 4-arm | Individual | CRF | Normal | OW/Obese | SF | COMPASS Composite | 10 | CBF, BB |
| Barnes (2023) | USA | 604/564 | 70.4 (4.2) | 65.0 | 12.2 | Parallel, 2-arm | Multi-site | CRF | Normal | FHAD, OW/Obese | TD | Composite | 12 | MRI, BB |
| Bartholomew (2021) | USA | 103/71 | 48.1 (10.9) | 67.0 | 5.3 | Parallel, 2-arm | Individual | Clinic | Normal | Metabolic Syndrome | TD | MicroCog | -- | BB |
| Blondal (2022) | Iceland | 106/104 | 82.6 (6.4) | 62.3 | -- | Parallel, 2-arm | Individual | Community | At-Risk | Malnutrition | TD | MMSE | -- | -- |
| Blumenthal (2019) | USA | 160/160 | 65.4 (6.8) | 66.3 | 46.9 | Factorial, 4-arm | Single-site | CRF | MCI | CVD risk | MD, TD | NTB, CDR | 10 | -- |
| Boespflug (2017) | USA | 21/16 | 78.0 (6.5) | 56.3 | 12.5 | Parallel, 2-arm | Individual | CRF | MCI | -- | SF | -- | 1 | fMRI |
| Bohn (2021) | Norway | 64/60 | 71.5 (7.1) | 0 | -- | Parallel, 2-arm | Individual | Community | At-Risk | -- | SF | CANTAB | 5 | BB |
| Bookheimer (2013) | USA | 32/28 | 62.6 (7.8) | 75.1 | -- | Parallel, 2-arm | Individual | Clinic | At-Risk | -- | SF | -- | 4 | fMRI, BB |
| Bowtell (2017) | UK | 26/26 | 68.3 (3.2) | 50.0 | -- | Parallel, 2-arm | Individual | CRF | Normal | -- | SF | Composite | 7 | fMRI, BB |
| Brinkworth (2009) | Australia | 118/69 | 50.0 (8.2) | 64.8 | -- | Parallel, 2-arm | Single-site | Clinic | Normal | Obese, Metabolic Syndrome | TD | -- | 2 | -- |
| Brodaty (2025) | Australia | 6104/3500 | 64.9 (5.9) | 64.0 | 4.0 | Parallel, 2-arm | Individual | Community | At-Risk | -- | MD | MYB, Creyos | 8 | -- |
| Buchholz (2024) | USA | 38/22 | 74.2 (5.8) | 47.4 | 13.2 | Parallel, 2-arm | Individual | CRF | MCI, early AD | -- | TD | Composite | 2 | BB |
| Cardoso (2016) | Brazil | 31/20 | 77.7 (5.3) | 70.0 | -- | Parallel, 2-arm | Single-site | CRF | MCI | -- | SF | CERAD | 5 | -- |
| Chai (2019) | USA | 37/34 | 69.8 (3.7) | 54.1 | 13.5 | Parallel, 2-arm | Single-site | ARI | Normal | -- | SF | CANTAB | 5 | BB |
| Chan (2017) | Malaysia | 40/22 | -- >70 | 65.0 | 85.0 | Parallel, 2-arm | Individual | Community | AD | -- | SF | MMSE | 2 | BB |

| | | | | | | | | | | | | | | |
|---|---|---|---|---|---|---|---|---|---|---|---|---|---|---|
| Chatterjee (2022) | India | 60/60 | 68.3 (6.0) | 36.7 | -- | Parallel, 4-arm | Single-site | CRF | Normal | -- | MD | -- | 2 | -- |
| Cheatham (2023) | USA | 131/107 | 72.2 (4.3) | 55.8 | 3.1 | Parallel, 3-arm | Individual | Community | Normal | -- | SF | CANTAB | 2 | EEG |
| Chlebowski (2020) | USA | 1606/1452 | 69.9 (3.6) | 100 | 14.4 | Parallel, 2-arm | Individual | Community | Normal | -- | TD | MMSE (3MSE) | -- | -- |
| Chou (2022) | Taiwan | 84/80 | 72.2 (5.7) | 83.8 | -- | Cluster, 2-arm | Multi-site | Community | Normal | -- | MD | MoCA | 1 | -- |
| Coates (2020) | Australia | 151/128 | 65.0 (8.0) | 100 | 7.3 | Parallel, 2-arm | Individual | CRF | Normal | OW/Obese | SF | COMPASS, Composite | 6 | BB |
| Curtis (2024) | USA | 24/20 | 76.3 (7.9) | 58.3 | 0.0 | Parallel, 2-arm | Individual | CRF | MCI | -- | SF | CDR, MMSE | 5 | BB |
| Halyburton (2007) | Australia | 121/93 | 50.2 (1.3) | 60.2 | -- | Parallel, 2-arm | Single-site | CRF | Normal | OW/Obese | TD | -- | 2 | BB |
| Han (2023) | Australia | 32/28 | 79.2 (6.6) | 62.5 | -- | Parallel, 2-arm | Single-site | Hospital | Normal | Frailty | MD | MMSE | -- | -- |
| Handajani (2020) | Indonesia | 90/84 | -- >60 | 71.4 | -- | Parallel, 3-arm | Individual | CRF | MCI | -- | SF | MMSE | 3 | -- |
| Hardman (2020) | Australia | 102/81 | 77.5 (6.9) | 72.5 | -- | Factorial, 4-arm | Individual | Retirement home | Normal | -- | MD, TD | SUCCAB | 9 | -- |
| Horie (2016) | Brazil | 80/75 | 68.1 (4.9) | 83.8 | -- | Parallel, 2-arm | Single-site | Clinic | MCI | Obese | TD | CAMCog | 14 | BB |
| Hoscheidt (2021) | USA | 87/84 | 56.3 (5.1) | 66.7 | -- | Parallel, 2-arm | Single-site | Clinic | Normal, MCI | -- | TD | Composite | 2 | CSF, MRI, BB |
| Jakobsen (2011) | Denmark | 26/23 | 24.2 (3.5) | 0.0 | -- | Parallel, 2-arm | Single-site | ARI | Normal | -- | TD | ACE | 3 | -- |
| James (2024) | USA | 58/49 | 50.1 (5.1) | 86.0 | 29.0 | Parallel, 2-arm | Multi-site | Community | Normal | Obese | TD | T-MoCA | 1 | -- |
| Jennings (2024) | UK | 104/99 | 67.4 (4.6) | 74.0 | 1.0 | Parallel, 3-arm | Multi-site | Community | Normal | -- | MD, TD | NTB, STT, Composite | 8 | -- |
| Kamoun (2024) | Tunisia | 28/20 | 66.7 (2.4) | 0.0 | -- | Parallel, 2-arm | Individual | Community | Normal | -- | SF | MoCA | -- | -- |
| Keawtep (2024) | Thailand | 92/80 | 52.8 (3.4) | 100 | -- | Parallel, 4-arm | Multi-site | Community | Normal | OW/Obese | MD, TD | MoCA | 2 | BB |
| Kimble (2022) | UK | 56/50 | 48.0 (6.0) | 68 | -- | Parallel, 2-arm | Single-site | Community | Normal | OW/Obese, High BP | SF | COMPASS | 3 | -- |
| Knight (2016) | Australia | 166/137 | 72.0 (4.9) | 53.3 | -- | Parallel, 2-arm | Individual | Community | Normal | -- | TD | Composite | 11 | -- |
| Koblinsky (2022) | Canada | 14/11 | 72.0 (5.0) | 71.4 | -- | Cluster, 2-arm | Single-site | Community | MCI | CVD risk factors | MD | COMPASS, MoCA | | |
| Komulainen (2021) | Finland | 1401/1190 | 66.5 (5.4) | 50.7 | -- | Parallel, 6-arm | Individual | Community | Normal | -- | MD, TD | CERAD, MMSE | 7 | -- |

| | | | | | | | | | | | | | | |
|---|---|---|---|---|---|---|---|---|---|---|---|---|---|---|
| Krikorian (2009) | USA | 12/12 | 78.2 (5.0) | 33.3 | -- | Parallel, 2-arm | Individual | Community | MCI | -- | SF | -- | 3 | BB |
| Krikorian (2012a) | USA | 21/21 | 76.5 (5.5) | 47.6 | -- | Parallel, 2-arm | Individual | Community | MCI | -- | SF | CDR MOCA | 2 | BB |
| Krikorian (2012b) | USA | 23/23 | 70.1 (6.2) | 56.5 | -- | Parallel, 2-arm | Individual | Community | MCI | -- | TD | CDR, MoCA | 1 | BB |
| Krikorian (2022) | USA | 33/27 | 56.4 -- | -- | -- | Parallel, 2-arm | Individual | Community | At-risk | OW/Obese | SF | Composite | 5 | BB |
| Krikorian (2023) | USA | 34/30 | 56.4 (4.3) | 83.3 | -- | Parallel, 2-arm | Individual | Community | At-risk | OW/Obese | SF | -- | 5 | BB |
| Lee (2017) | USA | 13/10 | 72.2 (3.8) | 50.0 | 10.0 | Parallel, 2-arm | Single site | ARI | MCI | -- | SF | ADAS-Cog MMSE | 19 | FDG-PET |
| Lee (2023) | South Korea | 49/46 | 75.2 (5.7) | 57.4 | -- | Parallel, 3-arm | Single site | Clinic | MCI/AD | -- | MD | RBANS | 8 | PET BB |
| Liang (2021) | Taiwan | 733/555 | 74.0 (5.7) | 66.6 | -- | Cluster, 2-arm | Multi-site | Community | Normal | -- | MD | MoCA | 7 | -- |
| Makris (2013) | USA | 47/47 | 47.4 (8.7) | 46.8 | 30.0 | Parallel, 2-arm | Multi-site | CRF | Normal | Obese | TD | -- | 4 | -- |
| Marseglia (2018) | Europe | 1279/1144 | 70.9 (3.4) | 56.3 | -- | Parallel, 2-arm | Multi-site | Community | Normal | -- | TD | CERAD, MMSE | 8 | -- |
| Martin (2007) | USA | 48/48 | 37.5 (1.9) | 56.3 | 37.5 | Parallel, 4-arm | Multi-site | CRF | Normal | OW/Obese | MD, TD | -- | 4 | -- |
| Martinez-Lapiscina (2013a) | Spain | 1055/522 | 67.4 (5.7) | 55.4 | -- | Parallel, 3-arm | Single-site | CRF | Normal | T2DM, Vascular Risk | TD | MMSE | 1 | -- |
| Masley (2008) | USA | 56/47 | 45.3 (10.4) | 53.5 | -- | Parallel, 2-arm | Individual | Community | Normal | -- | MD | -- | 7 | BB |
| Mazza (2018) | Italy | 180/110 | 70.0 (4.0) | -- | -- | Parallel, 2-arm | Individual | Community | Normal | -- | SF | MMSE, ADAS-Cog | 1 | -- |
| McMaster (2020) | Australia | 119/101 | 73.1 (5.5) | 61.4 | -- | Parallel, 2-arm | Individual | Community | MCI | -- | MD | ADAS-Cog, Composite | 5 | -- |
| Mendoza-Ruvalcaba (2015) | Mexico | 83/64 | 70.6 (6.8) | 89.0 | -- | Parallel, 2-arm | Individual | Community | Normal | -- | MD | -- | 2 | -- |
| Miller (2018) | USA | 42/37 | 67.5 (4.6) | 67.4 | -- | Parallel, 2-arm | Single-site | ARI | Normal | -- | SF | -- | 6 | -- |
| Miller (2021) | USA | 47/37 | 67.6 (4.4) | 40.5 | -- | Parallel, 2-arm | Single-site | ARI | Normal | -- | SF | -- | 6 | -- |
| Mirheidary (2019) | Iran | 72/53 | 20.8 (1.9) | 81.1 | -- | Parallel, 2-arm | Individual | ARI | Normal | -- | SF | -- | 2 | -- |
| Moon (2021) | South Korea | 152/136 | 70.9 (4.8) | 74.3 | -- | Parallel, 3-arm | Multi-site | Clinic | At-Risk | -- | MD | RBANS, MMSE, CDR | 11 | BB |

| | | | | | | | | | | | | | | |
|---|---|---|---|---|---|---|---|---|---|---|---|---|---|---|
| Nakazeko (2023) | Japan | 110/93 | 64.9 (3.7) | 50.0 | -- | Parallel, 2-arm | Single-site | Clinic | Normal, MCI | Frailty | MD | -- | 1 | BB |
| Napoli (2014) | USA | 107/93 | 69.8 (4.0) | 62.6 | 15.0 | Parallel, 4-arm | Single-site | CRF | Normal | Obese, Frailty | MD, TD | 3MSE (MMSE) | 3 | BB |
| Ngandu (2015) | Finland | 1260/1105 | 69.3 (4.7) | 46.3 | -- | Parallel, 2-arm | Multi-site | Clinic | At-Risk | -- | MD | NTB, MMSE | 14 | BB |
| Ornish (2024) | USA | 51/49 | 73.5 (7.9) | 40.2 | 21.2 | Parallel, 2-arm | Multi-site | Community | MCI, early AD | -- | MD | ADAS-Cog, CDR | -- | BB |
| Parilli-Moser (2021) | Spain | 90/63 | 22.7 (3.1) | 69.8 | -- | Parallel, 3-arm | Single-site | ARI | Normal | -- | SF | -- | 10 | BB |
| Rakic (2022) | USA | 68/60 | 61.5 (6.3) | 45.0 | 33.0 | Parallel, 3-arm | Single-site | CRF | Normal | OW/Obese | SF | CANTAB | 7 | BB |
| Reeder (2022) | USA | 75/58 | 20.1 (1.5) | 93.8 | 26.2 | Parallel, 2-arm | Individual | ARI | Normal | -- | SF | CNS Vital Signs, NCI | 11 | -- |
| Rizvi (2024) | Pakistan | 90/90 | 48.8 (7.9) | 37.8 | -- | Parallel, 3-arm | Multi-site | Community | Normal | OW/Obese | TD | -- | 1 | BB |
| Roach (2023) | USA | 55/35 | 74.7 -- | 32.7 | 7.5 | Parallel, 2-arm | Single-site | Community | MCI, early AD | -- | MD | MoCA. | 3 | BB |
| Rodrigo-Gonzalo (2023) | Spain | 80/74 | 76.7 (4.6) | 66.3 | -- | Parallel, 2-arm | Single-site | Community | Normal | -- | SF | MoCA. MMSE | 19 | -- |
| Rutledge (2021) | USA | 38/38 | 67.6 (4.6) | 67.5 | -- | Parallel, 2-arm | Single-site | CRF | Normal | -- | SF | -- | 2 | -- |
| Sakurai (2024) | Japan | 531/406 | 74.4 (4.9) | 52.0 | -- | Parallel, 2-arm | Multi-site | Community | MCI | -- | MD | MMSE, Composite | 6 | BB |
| Sala-Vila (2020) | Spain | 708/636 | 69.2 (3.7) | 66.8 | -- | Parallel, 2-arm | Single-site | Clinic | Normal | -- | SF | Composite | 18 | fMRI |
| Siddarth (2019) | USA | 261/200 | 60.4 (6.5) | 66.7 | -- | Parallel, 2-arm | Single-site | CRF | MCI | -- | SF | -- | 6 | MRI, BB |
| Silver (2023) | USA | 218/182 | 38.1 (7.2) | 70.0 | -- | Parallel, 2-arm | Multi-site | CRF | Normal | -- | TD | -- | 1 | -- |
| Smith (2010) | USA | 124/120 | 52.3 (9.6) | 63.7 | 39.0 | Parallel, 3-arm | Single-site | CRF | Normal | OW/Obese, Elevated BP | MD, TD | -- | 8 | -- |
| Smith (2020) | USA | 160/132 | 67.4 (6.8) | 62.9 | 50.8 | Factorial, 4-arm | Single-site | Clinic | MCI | CVD risk | MD, TD | -- | 9 | BB |
| Thunborg (2024) | Europe | 93/85 | 73.2 -- | 53.8 | -- | Parallel, 3-arm | Multi-site | Clinic | MCI | Cardio-metabolic Risk | MD | CDR | 2 | -- |
| Tussing-Humphreys (2022) | USA | 185/149 | 66.3 (6.1) | 85.9 | 1.1 | Parallel, 3-arm | Multi-site | Community | MCI | OW/Obese | TD | Composite | 13 | -- |
| Uchiyama-Tanaka (2024) | Japan | 43/39 | 79.0 (8.8) | 59.5 | -- | Parallel, 3-arm | Single-site | Clinic | Normal | -- | TD | MCIS | -- | BB |

| | | | | | | | | | | | | | | |
|---|---|---|---|---|---|---|---|---|---|---|---|---|---|---|
| Valls-Pedret (2015) | Spain | 447/334 | 66.8 (5.5) | 50.9 | -- | Parallel, 3-arm | Single-site | CRF | Normal | T2D or OW/Obese or CVD risk | TD | MMSE, Composite | 5 | -- |
| Wardle (2000) | UK | 176/155 | 53.0 (10.0) | 51.7 | -- | Parallel, 3-arm | Single-site | Clinic | Normal | High cholesterol | TD | -- | 4 | -- |
| Wood (2023) | UK | 66/54 | 70.1 (3.7) | 60.7 | -- | Parallel, 2-arm | Individual | ARI | Normal | -- | SF | -- | 4 | BB |
| Zhu (2024) | China | 120/107 | 19.0 (1.0) | 85.0 | -- | Parallel, 4-arm | Single-site | ARI | Normal | -- | MD, TD | -- | 2 | -- |
| Zulke (2024) | Germany | 1030/819 | 69.0 (4.9) | 52.9 | -- | Cluster, 2-arm | Multi-site | Community | At-Risk | -- | MD | CERAD Composite | 6 | BB |

*Note.* 3MSE = Modified Mini-Mental State Examination; ACE-III = Addenbrooke's Cognitive Examination-III; ARI = academic research institute; BB = Blood Biomarkers; CAMCog = Cambridge Cognitive Examination for the elderly; CANTAB = Cambridge Neuropsychological Test Automated Battery; CERAD = Consortium to Establish a Registry for Alzheimer's Disease; CDR = Clinical Dementia Rating; COMPASS = Oxford COMPetency ASSessment; CRF = clinical research facility; CVD = cardiovascular disease; FHAD = 1$^{st}$ degree family history of Alzheimer's Disease; INT = intervention; MCI = mild cognitive impairment; MD = multi-domain dietary intervention; MMSE = Mini-Mental State Examination; MMSE-2 = Mini-Mental State Examination-Second Edition; MoCA = Montreal Cognitive Assessment; NTB = Neurocognitive Test Battery; OW = overweight; SF = single-food dietary intervention; SUCCAB = Swinburne University Computerised Cognitive Assessment Battery; TD = total diet intervention; T2D = Type II Diabetes Mellitus

**Supplementary Table S4a.** Multidomain intervention study characteristics (*n* = 30)

| **Multi-Domain Dietary Intervention Studies (n = 30)** | | | | | | | | | | |
|---|---|---|---|---|---|---|---|---|---|---|
| **Author (Year)[Ref]** | **Intervention Sample Size (T0/T1)** | **Duration (Weeks)** | **Diet Type** | **Components** | **Intervention Dosage/Protocol** | **Intervention Adherence** | **Guided Support (Y/N)** | **Frequency of Support** | **Control Sample Size (T0/T1) Condition** | **Control Description** |
| Andrieu (2017)<br><br>*Chhetri (2018)[b]* | 837/760<br><br>417/374 MD+PUFA<br><br>420/390 MD Only | 156 | French Nutrition Guidelines | PA, CT, GP, ± PUFA supplement | **12x2-hr sessions**<br>**Nutrition:** 15-min diet advice/session;<br>**PA:** 45-min/session;<br>**CT:** 60-min/session.<br>**GP:** 3 preventive consultations;<br>**PUFA**: 2 x/day (800mg DHA, 225mg EPA) | 53-85% | Y, IP+group | Weekly (first 2mo), monthly (3-12mo); + 3 consults | 420/380<br><br>Placebo | Placebo capsule matched for appearance/ taste, no multi-domain |
| Blumenthal (2020)<br><br>*Blumenthal (2019)* | 40/40 | 24 | DASH | PA | **Diet**: 18 30-min sessions: Weekly (0-3mo) then biweekly (3-6mo) to meet DASH guidelines<br>**PA**: Aerobic exercise (AE), moderate intensity walking or stationary cycling, 35-min/session, 3x/week | 88.6-99.6% | Y, IP+group | Exercise: 3mo supervised; DASH: weekly (0-3mo) / biweekly (3-6 mo) group counselling | 38/38<br><br>Attention-Matched AC | 18 30-min sessions: Weekly/ biweekly phone calls on CVD health topics; no diet/PA intervention |
| Brodaty (2025) | 3051/1403 | 52 | MEDI | PA, CT, CBT | **Online modules.**<br>**Diet:** MEDI diet (7 days/week)<br>**PA:** 150-300 mins MVPA per week; and ST >=3 days/week, vigorous intensity; balance training 7 days/week | 1/3 of intervention group completed ≥ 60% of tasks; 1/3 did some (1-59%); 1/3 did none | Partial Y, Automated digital SMS | Weekly for 10 weeks; then monthly | 3053/1997<br><br>AC | Access to same modules; Static public health information; No tailored goals or |

| | | | | | | | | | | |
|---|---|---|---|---|---|---|---|---|---|---|
| | | | | | **CT:** 3 x week, 45min per session (10 weeks) + Monthly boosters.<br>**CBT:** CBT-based 6 modules over 10 weeks for depression/ anxiety | | | | | feedback or monitoring. |
| Chatterjee (2022) | 30/30<br><br>15/15 MEDI+CT<br><br>15/15 MEDI+CT+PA | 24 | MEDI | PA, CT | **Diet:** 20% daily energy from protein, 25% fat (5% max saturated/trans fat), 20% PUFA, 55% carbs (<10% refined sugar), 30-35g fibre/day, < 5g salt/day, no alcohol<br>**CT:** 40 sessions, 2 sessions/week; 40-min/each, supervised.<br>**PA:** in-person, supervised 2x/week, 6 months; **AE:** walking 40-80% HRmax; **ST**: 2 sets 8-15 reps, 40-80% RM | 100% | Y, IP+Phone | Monthly IP check-in with dietitian + Weekly phone calls for diet; Supervised IP exercise and CT | 15/15<br><br>AC | Static health advice for brain stimulation and cognitive engagement (e.g., sudoku, mental math, new skills); structured monitoring by phone and bimonthly IP visits. |
| Chou (2022) | 43/41 | 8 | MEDI | CT | **Diet:** Mediterranean diet concepts, food preparation, and lifestyle skills; recipes and menus, guidebook<br>**CT:** Weekly 40-minute group sessions for 8-weeks of game-based learning through interactive card games; | 95% completion rate; Mediterranean diet scores pre-post: 10.48 to 12.70, *p* < .001 | Y, IP+group | 8 weekly 40-min group-based sessions by nutrition specialist | 41/39<br><br>Minimal HE | Taiwan National Health Agency standard booklet on balanced diet for older adults; no active dietary or cognitive intervention. |
| Han (2023) | 16/12 | 12 | INT, Australia | PA, SM | **Diet:** Individualised based on NNG to | 91%±13 (inpatient); | Y, IP+Phone+ home visits | In-hospital daily nutrition | 16/15 | Medical consultations |

| | | | | | | | | | | |
|---|---|---|---|---|---|---|---|---|---|---|
| | | | Nutrition Guidelines | | meet energy and protein needs (1-1.2 g/kg protein/day); oral supplements if needed, snacks, food fortification.<br>**PA:** Individualised, supervised inpatient: 5x/week, 30mins walking. Home-based walking and ST 3x/week (6 exercises, 3 sets of 8-10 reps)<br>**SM:** Motivational interviewing, printed guidebook | 92%±21 (home/ telehealth); Home-based exercise 66% ± 33% adherence; Nutrition 89% ± 17% of energy; 82% ± 20% of protein achieved; Mean daily intake 1604 ± 471 kcal, 71.1 ± 24.0g protein | | therapy; post-hospital 4 home visits + 4 phone calls over 12 weeks | TAU | and allied health professional care (e.g., dietitians, physiotherapists), during and after hospital, without intervention. |
| Hardman (2020) | 24/18 | 24 | MEDI | PA | **Diet:** 6-week MEDI diet meal plan as guidance. Received EVOO and instructed to use exclusively.<br>**PA:** 30-40 min/day walking with pedometer | Median self-reported adherence: 50 – 70% compliance to MEDI. | Partial Y, in-person | Initial and 3-month dietary counselling | 27/25<br><br>Passive Control (PC) | Maintain current lifestyle for a period of 6 months |
| Jennings (2024) | 35/33 | 24 | MEDI | PA | **Online platform:** personalised targets, self-assessment, tailored feedback. “Eating Well” and “Moving More” Modules.<br>**Diet**: Personalised MEDI nutrition education, recipe ideas and 4 (2hr) group sessions on behaviour change. Intervention goal: improve MEDAS score by ≥3 points. Food | **Platform Use:** 84% accessed online monthly or less (avg. session 15-30min)<br><br>**Group Sessions:** Mean 3.5/4 (SD = 0.9) attendance<br><br>**Food Voucher Uptake:** 100%<br><br>**Diet**: Median increase in | Y, IP+virtual+ web-based | Four group sessions (at weeks 0, 2, 4, and 12) dietary guidance, support, and information about the MEDI diet. | 34/32<br><br>Low-intensity AC | Brief dietary and PA advice based on UK NICE guidelines for CVD risk; 1-hour group session at baseline; no further active intervention; vouchers provided after trial. |

| | | | | | | | | | | |
|---|---|---|---|---|---|---|---|---|---|---|
| | | | | | vouchers (£30/week) /delivery.<br>**PA:** Personalised PA suggestions and goals, guidance on barriers. Intervention target: increase PA to 150 min moderate, or 75 vigorous/week. | MEDAS score = 3.7 points at 24 weeks<br><br>**PA:** steps, energy expenditure and mins of MVPA increased | | | | |
| Keawtep (2024) | 23/19 | 12 | IF | PA, CT | **Diet**: IF 2 days/week - weeks 1-4 (75% EER); weeks 5-8 (50% EER); weeks 9-12 (25% EER), *Ad libitum* eating 5 days/week<br>**AE+ST+CT:** 60min/day, 3 days/week, 3 months (home-based, YouTube guided) + progression via movement/cognitive challenge | **Diet+PA**: 87.30% (Diet), 93.01% (AE+ST+CT) adherence | Y, IP+Phone. +email+SMS | Weekly contact; mobile app reminders; single IP 2-hr training | 23/20<br><br>PC | Continue routine activities and usual lifestyle behaviour. |
| Koblinsky (2022) | 7/7 | 24 | Baycrest Brain Healthy Diet (MEDI+DASH) | PA | **Diet**: 1-hr/week group dietary counselling, education, goals setting + optional 30-min individual counselling with dietitian; weekly goal setting and monthly self-assessments<br>**AE+ST**: 1 hr/week supervised AE and ST; 5 AE, 2-3 ST sessions/week. | 90% exercise an 92% diet session attendance; 100% monthly self-assessment completion; 61% weekly exercise logs completed | Y, IP+group+ individual | Weekly 1hr/week group session for diet + individual 30-min as needed; monthly self-assessment w/ support | 7/7<br><br>Attention-Matched Placebo | Time-matched placebo brain health education; 2 individual sessions; same frequency of social contact. |
| Komulainen (2021) | 466/379<br><br>234/207 AE+Diet | 208 | Finnish Nutrition Guidelines | PA | **Diet:** 5 individualised sessions in year 1, then every 6-months; ≥400g/day of | **AE+Diet Group:** 71% adherence (57% for aerobic | Y, IP+ individual+ group | 11 30-min IP session with nutritionist; 3 group | 236/201<br><br>Minimal HE | General advice on physical activity and |

| | | | | | | | | | | |
|---|---|---|---|---|---|---|---|---|---|---|
| *Komulainen (2010)* | 234/192 ST+Diet | | | | vegetables, fruit, and berries ≥2 servings fish/week (≥30g/day) ≥14g fibre/1000 kcal ≤10% from saturated fats (SFAs); After 6-months, more plant foods and less red meat.<br>**AE**: 60-min MVPA 5x/wk. 60% of VO2 max, duration and frequency increased gradually.<br>**ST:** 1x/wk of 1 set of 10 repetitions at 40% of 1-repetition maximum (1RM) for the major muscle groups. After 6 months, 2 x/week with 2 sets of 15 repetitions at 60% of 1RM. | exercise and 85% for diet)<br><br>**ST+Diet Group:** 66% adherence (47% for ST and 84% for diet) | | counselling sessions over 4-years | | diet at baseline. |
| Lee (2023) | 16/15 | 8 | MIND | PA, CT, GP, VRM, Motivation, NS | **Diet**: MIND diet Korean context. More plant foods, healthy oils and fish; less red meat, sugar, and UPFs<br>**CT**: 2x 60-min sessions/week: 1 IP group session/week on global cognitive function; 1 home-based session via app. Weekly Zoom homework. | **MD Adherence:** 96-100% in all components; CT: 96.1%; PA: 94%; VRM: 100%; Diet Guidance: 100%; 99.1% Supplement<br><br>**NS Adherence:** 83.7% | Y, IP+ individual+ group+SMS | 30-min individual IP counselling every 3-weeks + 7 50-min group sessions, 2x/week CT and PA; Monthly IP VRM; Weekly SMS motivation | 17/16<br><br>HE WLC | Waitlist, dementia prevention education and usual care. Offered program after intervention period. |

| | | | | | | | | | | |
|---|---|---|---|---|---|---|---|---|---|---|
| | | | | | **PA**: 3 60-min sessions/week of combined AE and ST.<br>**VRM:** Personalised lifestyle modifications booklet based on risk factors.<br>**Motivation:** Baseline IP session; Weekly motivational SMS messages 2x/week.<br>**NS:** 2x/day; 2 x 150mL/day (EPA, DHA) | | | | | |
| Liang (2021) | 382/304<br><br>126/101 Normal<br><br>47/33 CD<br><br>104/87 Frailty<br><br>105/83 PCDS | 52 | Taiwan Nutrition Guidelines | PA, CT, HE | **Diet:** 15-mins general diet advice per session, 16 sessions/12 months<br>**ST:** 45-min group sessions, 16 sessions/12 months<br>**CT:** 1-hour group sessions; 16 sessions/12 months<br>**HE:** Chronic disease prevention | *NR* | Y, IP+group+ SMS | Weekly (4 sessions) in 1st month; 2 sessions biweekly in 2nd month; 1 session/mo for 10 months | 351/251<br><br>68/53 Normal<br><br>63/39 CD<br><br>120/83 Frailty<br><br>100/76 PCDS<br><br>AC | Periodic telephone calls offering general health advice without the structured interventions |
| Martin (2007) | 12/12 | 24 | CR | PA | **CR:** 12.5% caloric reduction in intake<br>**PA:** 12.5% increased energy expenditure through structured AE All meals provided in weeks 1 – 12; 22 – 24. Self-selected diet to adhere to CR weeks 13 – 21. | 96% completion of weekly behavioural meetings and meal provision | Y, IP+group | Weekly group meetings; daily monitoring during meal provision period | 12/11<br><br>Attention-Matched Isocaloric PC | Weight maintenance diet; food supplied. same weekly meeting contact |

| | | | | | | | | | | |
|---|---|---|---|---|---|---|---|---|---|---|
| Masley (2008) | 28/27 | 10 | High-Fibre, Low Saturated Fat | PA, SM | **Diet:** >30g fibre/day and <16g saturated fat/day; meal plans and recipes provided **PA:** AE 30-min 5-6 days/week at 70-85% max HR; ST 3 days/week, 1-2 sets of 10-15 reps for 10 body movements to exhaustion **SM:** 10-20 min/day of stress management activities (e.g., meditation, yoga, breathing, bath) | 100% attendance required for intervention group; 10 sessions. PA = 48% adherence. Diet = >70% adherence. | Y, IP+group | Weekly 1.5-hr group sessions with nutritionist and ACSM-certified exercise instructors | 28/20 WLC | Current dietary intake and activity level for the duration of the study. Offered full intervention after study end. |
| McMaster (2020) | 57/48 | 8 | MEDI | PA, CT | 4 weekly online educational modules covering dementia risk factors, the MEDI diet, PA, and CT. **Diet**: 1 x 1-hr session (week 3) to help follow the MEDI diet. **PA**: 1 x 1-hr session (week 7) with an exercise physiologist to create a personalized exercise plan. **CT**: 2-hr/week starting at week 5, using the BrainHQ platform. | 100% adherence for educational modules, Dietitian sessions and Exercise physiologist sessions 20% adherence on Brain training (10.8 hrs of planned 54) | Y, IP+ individual + online | 2 single x 1-hr sessions | 62/48 AC | Participants received four online educational modules on dementia risk factors, the Mediterranea n diet, physical activity, and cognitive engagement, with no additional active components. |
| Mendoza-Ruvalcaba (2015) | 31/27 | 8 | General Healthy Eating | PA, CT, RO | 16 group sessions (2-hr x twice weekly). **Diet:** 8 90-min sessions; Healthy eating, proportions, and meal planning | *NR* | Y, IP+group | 2 x/week | 33/30 Attention-Matched WLC | While on wait list, engaged in weekly social activities organized by |

| | | | | | | | | | | |
|---|---|---|---|---|---|---|---|---|---|---|
| | | | | | **CT**: 8 90-min sessions; Memory, attention, and processing speed enhancement<br>**PA:** 30-min every session focused on strength, balance, and mobility<br>**RO:** Warm-up activities to reinforce awareness of time, place, person.<br>User manual and personalised plan. | | | | | the senior centre. |
| Moon (2021) | 102/94<br><br>51/45<br>FMI<br><br>51/49<br>HMI | 24 | MIND | PA, CT, VRM, SE, Motivation | **FMI:** Facility, 3x/week<br>**Diet:** 3 individual and 7 group sessions<br>**CT/SE:** 2/wk, 50-min, group<br>**PA:** 3/wk, 60-min, group<br>**Motivation:** 4 group sessions and weekly self-assessments<br><br>**HMI:** Group session every 1-2 weeks at facility, rest at home<br>**Diet**: 3 Individual, 4 group, 3 home-based (workbook) on the MIND diet; bi-weekly diet checks.<br>**CT/SE:** 1 group, 1 home-based session per week for 2 mos, bi-weekly group sessions and 1-2 | FMI Total adherence 94.5% (95%CI: 91.4-97.6%)<br><br>VRM: 98.0%, CT: 97.4%, SE: 95.9%, PA 91.0%, Diet: 94.2%, Motivation: 97.7%<br><br>HMI total adherence 96.8% (95%CI:95.4-98.1%)<br><br>VRM: 100%, CT: 97.6%, SE: 100%, | Y, IP+group+ individual+ digital | FMI: 3x/week facility<br><br>HMI: 1-2x/month facility + weekly digital | 50/42<br><br>TAU+HE<br>WLC | General health advice, prescribed medications as needed, and educational booklets on lifestyle changes to reduce dementia risk. Met with a study doctor for VRM. Offered intervention after study. |

| | | | | | | | | | | |
|---|---|---|---|---|---|---|---|---|---|---|
| | | | | | home-based sessions.<br>**PA:** 3 60-min sessions per week, increased intensity/content every 2 months.<br>**Motivation**: Weekly self-assessments, family videos, dementia-related articles via mobile. | PA: 95.0%,<br>Diet: 94.4%,<br>Motivation: 99.4% | | | | |
| Nakazeko (2023) | 55/43 | 12 | COMpletely Balanced for Frailty Prevention (COMB-FP) | PA | **COMB-FP Diet:** Participants replaced two meals per day with COMB-FP meals for 12 weeks. Usual diet for 3rd meal. Meets Japanese DRI for 33 nutrients.<br>**PA**: Group ST exercise at weeks 1, 4, 8, and 10; 20-min home ST exercise 3-5 times per week. Provided textbook and exercise video. | > 90% of the test meals consumed (self-report);<br><br>Exercise frequency:<br>INT: 4.1x/wk ± 0.7<br>Control: 4.2x/week ± 0.8 | Y, IP+group | 4 IP group sessions Week 1, 4, 8, 10 | 55/50<br><br>AC | *Ad libitum* Diet and same exercise program as intervention. |
| Napoli (2014) | 28/25 | 52 | CR | PA | **Diet**: 500–750 kcal/day energy deficit for 10% weight loss in 6 months, maintained for 6 months. Weekly weigh-in, group therapy, weekly food diary.<br>**PA**: 90-min supervised group sessions 3x/wk; 15-min flexibility, 30-min AE (65-85% max HR), | Diet: 82% (IQR: 76-89%)<br><br>PA: 83% (IQR: 80-88%) | Y, IP+group+ individual | Weekly IP with RD 3x/week (PA) | 27/23<br><br>Elimination HE | General information about a healthy diet. Instructed not to join any weight loss or PA program. |

| | | | | | | | | | | |
|---|---|---|---|---|---|---|---|---|---|---|
| | | | | | 30-min of ST (65-80% of 1-RM), and 15-min balance exercises. | | | | | |
| Ngandu (2015)<br><br>*Rosenberg (2018)* | 631/576 | 104 | Finnish Nutrition Guidelines | PA, CT, VRM | **Diet**: 3 individual and 7-9 group sessions with a nutritionist<br>**PA**: Individually tailored PA plan consisting of ST: 1-3x/wk, AE: 2-5x/week, supervised by PT<br>**CT:** 10 sessions; computer-based training at home for 10-15 mins/session 3/wk for 6 months (twice). Included a range of cognitive tasks.<br>**VRM:** 6 visits with study nurse/physician for anthropometric measurements and physical examination. Included risk review and lifestyle management. | 100% of participants attended nutrition sessions, 90% participated in PA, 85% engaged in CT, and 87% VRM.<br><br>Overall, 72% of participants adhered to all four intervention domains, while 1% participated in only one domain. | Y, IP+group+ Individual | Monthly nutrition; Weekly PA/CT | 629/592<br><br>TAU+HE | General health advice on VRM at baseline (diet, PA, weight) and regular health assessments (e.g., blood pressure, BMI). No structure intervention. Usual care follow-up. |
| Ornish (2024) | 26/24 | 20 | Vegan Diet | PA, SM, HE, SS, Supplements | **Diet:** A whole food, plant-based diet; 14-18% kcal fat; 16-18% protein, 63-68% complex carbohydrate; no calorie restriction; provided 21 meals per week (three meals per day plus snacks).<br>**PA:** 30 min/day of AE and ST 3x/wk, | *NR* | Y, IP+Online+ group+ individual | 3x/week for 4-hrs per session | 25/25<br><br>WLC | Asked not to make any lifestyle changes for 20 weeks, after which they would be offered the intervention. |

| | | | | | | | | | | |
|---|---|---|---|---|---|---|---|---|---|---|
| | | | | | supervised by exercise physiologist.<br>**SM**: 1-hr/day of meditation, yoga, stretching, and relaxation techniques), supervised by a certified specialist.<br>**SS:** 1hr, 3x/wk with spouses in support groups.<br>**Education**: One hour of lifestyle education per week.<br>**Supplements:** Daily intake of various supplements (e.g., omega-3, curcumin, multivitamins, coenzyme Q10, vitamin C) | | | | | |
| Roach (2023) | 31/22 | 104 | MIND | PA, CT, SM, SS, Sleep, MA | **Diet:** Monthly phone coaching with RD or nutritionist on MIND diet recommendations.<br>**PA:** Personalised plan based on US public health guidelines.<br>**CT:** BrainHQ online training.<br>**MA:** Support to follow prescriptions.<br>**Fitbit** for tracking PA, HR, Sleep | 86% stayed engaged with their coach >= 1year. 77% completed 2-years. | Y, Phone+ Email+SMS | 1/month phone + weekly email/SMS | 24/13<br><br>SC | Standard care for Alzheimer's Disease, which included usual clinical practices and interventions. 3 coaching calls after 24 months. |
| Sakurai (2024) | 265/215 | 78 | Japanese Nutrition Guidelines | PA, CT, VRM | **Diet:** Well-balanced, diverse diet; increase fish, seafood, dairy, | PA attendance mean 64.9/78 sessions (83%); CT mean 69.2 | Y, IP+Phone+ group+ individual | Monthly nutrition; weekly PA; 1x IP + 4x phone | 266/218<br><br>TAU | Treatment for diabetes, hypertension, and |

| | | | | | | | | | | |
|---|---|---|---|---|---|---|---|---|---|---|
| | | | | | fruit, vegetables, green tea<br>**PA**: 90-min 1x/week with target of 78 sessions including AE, ST, stretching, dual-task, and group discussion; self-monitoring with FitBit<br>**CT**: BrainHQ online ≥30min/day at least, 4 days/wk during three intensive 3-mo training periods.<br>**VRM:** Medical management per guidelines. | ±96.9 days (18% met high adherence ≥156 days) | | counselling for 6-months, up to 3 IP + 12 calls over 18 months; | | dyslipidaemia, along with general health-related information every 2 months. |
| Smith (2010) | 43/40 | 16 | DASH | PA, BM | **Diet:** Weekly 30-min DASH diet education classes.<br>**PA**: 30-min, 3x/wk of supervised AE<br>**BM**: 10% CR, weekly group behavioural weight loss sessions | 90% attendance PA sessions and 92% adherence of weekly DASH diet classes. | Y, IP+group+ individual | Weekly group diet/exercise sessions, PA 3x/wk | 43/42<br><br>PC | Participants maintained their regular diet and lifestyle without any intervention. |
| Smith (2020) | 35/35 | 24 | DASH | PA | **Diet:** 18 sessions; Dietary counselling for DASH diet, weekly 30-min for 12-weeks, then bi-weekly week 13-24; increase fruit, vegetables, whole grains, low-fat dairy, reduce saturated fat, cholesterol, sodium; promote potassium, magnesium, calcium and DASH score improvement. | *NR* | Y, IP+group+ individual | IP 18 sessions 1:1 diet counselling | 31/31<br><br>Attention-Matched HE | Weekly 30-min educational phone calls by health educator for first 3-mo then bi-weekly for next 3-mo on CVD topics; maintain usual diet and PA habits. |

| | | | | | | | | | | |
|---|---|---|---|---|---|---|---|---|---|---|
| | | | | | Educational materials provided.<br>**PA:** 43-min, 3x/wk of AE for 6 mo, supervised. | | | | | |
| Thunborg (2024) | 63/56<br><br>32/28 MD Only<br><br>31/28 MD+MF | 24 | Finnish Nutrition Guidelines +/- MF | PA, CT, VRM, SS | **Diet:** 3 individual and 3–4 group sessions by dietitians, based on Nordic or national dietary recommendations<br>**PA:** 60-min 2x/week; endurance and ST; Supervised by physiotherapists/personal trainers<br>**CT:** Group (2–3 sessions, 60–75 min) and home-based computer training (2×/week, 15–30 min)<br>**VRM:** baseline, 3- and 6-month meeting w/nurse for CVD assessment and management<br>**Medical Food:** 125 m**l** of the medical food drink (Fortasyn Connect) once a day | MD only: 71.9-81.3% domain adherence<br><br>MD+MF: 80.7-90.3% domain adherence<br>MF intake: 87.1% | Y, IP+group+ Remote | Monthly; 3 individual counselling and 3–4 group sessions | 30/29<br><br>Minimal HE | General health education on healthy diet and physical, cognitive, and social activities beneficial for VRM factors and preventing disability. |
| Tussing-Humphreys (2022) | 75/64 | 32 | MEDI+CR | PA | **MEDI Diet + CR** (~25% kcal reduction by 500-750 kcal/day and 5-7% weight loss); Weekly MEDI education, goal setting, 25 90-min group sessions and food supplied(1oz/day | Median attendance: 75%<br>MEDI Score: +6.3 points | Y, IP+ Individual+ group by dietitian/ nutritionist | Weekly group for 8-months (25 sessions) + 1 individual session at baseline | 37/35<br><br>HE | One 60-min individual session. No diet or PA advice. Weekly newsletters. Offered 1:1 session and self-directed |

| | | | | | | | | | | |
|---|---|---|---|---|---|---|---|---|---|---|
| | | | | | almonds, 3tbsp/day EVOO).<br>**PA:** 150 mins/week of MVPA. 30-min/wk supervised sessions by certified instructor. | | | | | intervention materials post-study. |
| Zhu (2024) | 30/28 | 12 | High-Fibre Diet | PA | **Diet**): ≥20g of dietary fibre per day from rice (5.4g) and fruits or recommended list (15g).<br>**PA:** Rope Skipping (RS) group: 3x/wk of 2,000 times; 100 jumps/set, 30s rest between sets. Supervised for first 3-weeks, then self-directed. | *NR* | Y, IP+group+ digital | 3x/week IP RS (week 1-3); daily WeChat reporting diet | 30/25<br><br>PC | *Ad libitum* diet and usual PA habits. Received diet and PA guidance after intervention. |
| Zulke (2024) | 546/378 | 104 | German Nutrition Guidelines | PA, CT, VRM, SS, MA, MH | **Diet:** Counselling at baseline, 12 and 24 mo. ≥ 5 servings fruit/veg/day, fish, low salt/sugar, hydration<br>**PA:** 3–5x/wk of AE step goal, pedometer, 2x/wk ST, flexibility, balance.<br>**CT:** 3 x/wk 15-min session NeuroNation<br>**SS**: Individual goals monitored regularly.<br>**MA:** GP for drug optimisation<br>**MH**: As needed; GP check at 12 and 24mo | Mean adherence scores (/4)<br>Diet: 2.80 (SD = 0.71)<br>CT: 2.90 (SD = 0.78).<br>PA: 2.66 (SD = 0.82).<br>SS: 2.81 (SD = 0.76). | Y, IP+phone+ digital | IP baseline, 12 and 24 months; Phone call at 2, 4, 8, 16, 20 months | 630/441<br><br>TAU+HE | General written health advice on nutrition, PA, CT, and GP TAU. No structured intervention. |

**Abbreviations:** AC: Active Control; ACSM: American College of Sports Medicine; AE: Aerobic Exercise; AGE: advanced glycation end-products; BM: Behavioural Management; CBT: Cognitive Behavioural Therapy; CD: Cognitive dysfunction; COMB-FP: COMpletely Balanced for Frailty Prevention; CR: Caloric Restriction; CT: Cognitive Training; CTRL: Control; CVD: Cardiovascular Disease; DASH: Dietary Approaches to Stop Hypertension; DHA: Docosahexaenoic acid; DRI: Daily recommended intake; EER: Estimated energy requirements; EPA: Eicosapentaenoic acid; EVOO: Extra-Virgin Olive Oil; FMI: Facility-based multidomain intervention; GI: Glycaemic index; GP: Doctor of general practice; HC:

High carbohydrate; HE: Health Education; HF: High fat; HMI: Home-based multidomain intervention; HR: Heart rate; IF: Intermittent Fasting; INT: intervention; IP: In-Person; IPAQ: International Physical Activity Questionnaire; IQR: Interquartile range; LC: Low carbohydrate; LCD: Low-calorie diet; LF: low fat; MA: Medical Adherence; MAD: Modified Atkins diet; MAF: macrophage-activating factor; MD: Multidomain; MEDAS: Mediterranean Diet Adherence Screener; MEDI: Mediterranean Diet; MF: Medical food; MH: Mental Health; MIND: Mediterranean-DASH Diet Intervention for Neurodegenerative Delay; MVPA: Moderate-Vigorous PA; N = No; NIA: National Institute on Aging; NNG: National Nutrition Guidelines; $NO_3^-$ = Nitrate ion; NR: Not Reported; NS: Nutritional supplement; PA: Physical Activity; PB: Peanut butter; PC: Passive Control; PCDS: Physio-cognitive decline syndrome; PUFA: Polyunsaturated Fatty Acids; RM: Repetition maximum; RO: Reality Orientation; RS: Rope skipping; SC: Standard Care; SD: Standard deviation; SE: Social Engagement; SM: Stress Management; SMS: Short Message Service; SRP: Skin roasted peanuts; SS: Social Support; ST: Strength Training; TAU: Treatment As Usual; UC: Usual care; UPF: Ultra processed foods; VLC: very low carbohydrate; VRM: Vascular Risk Management; WK: Week; WLC: Waitlist Control; Y = Yes.

**Supplementary Table S4b.** Total diet intervention study characteristics (*n* = 33)

| Total Diet Intervention Studies (n = 33) | | | | | | | | | |
|---|---|---|---|---|---|---|---|---|---|
| **Author (Year)** | **Intervention sample size (T0/T1)** | **INT Duration (Weeks)** | **Total Diet Type** | **Intervention Dosage/Protocol** | **Intervention Adherence** | **Guided Support (Y/N)** | **Frequency of Support** | **Control Sample Size (T0/T1) Condition** | **Control Description** |
| Arjmand (2022) | 22/22 | 12 | MIND+CR | MIND Diet principles; CR ≤500 kcal from baseline intake, 50–55% CHO, 30% fat, 15–20% protein. | ≥80% of meals/snacks met MIND protocol. | Y, IP+ individual+ Phone | Weekly | 18/15<br><br>AC | CR control diet with same macronutrient. Same contact. |
| Barnes (2023) | 301/275 | 156 | MIND+CR | MIND Diet; mild CR ≤250 kcal deficit/day; 14tbsp EVOO/wk; 5oz nuts/wk; 2.5c blueberries/wk | MIND diet score mean increase +3.3 points at 6 mo | Y, IP+phone+ individual+ online+ group by RD | 1x/week (month 1-6); biweekly (7-12mo); 2x/mo year 2-3; + ≥5 group sessions | 303/289<br><br>AC | CR control diet ≤250 kcal deficit/day; same contact |
| Bartholomew (2021) | 50/38 | 26 | IF | IF: 24-hr, water-only, 2x/wk for 4 weeks, Weeks 5-26 1x/wk; ad libitum on non-fasting days | 95% ± 12 %; 95% had ≥80% adherence | N | None | 53/33<br><br>PC | *Ad Libitum* Diet |
| Blondal (2022) | 53/52 | 24 | INT, Iceland Nutrition Guidelines | 8 nutrition therapy sessions (5 home visits + 3 phone calls). Daily hot meal & two snacks/day delivered weekly, 24-wks of supplements. Counselling with RD. | 100% received all planned visits, meals, calls | Y, IP+phone | 5 IP, 3 phone calls | 53/52<br><br>TAU | UC; healthy ageing diet booklet, Meals on Wheels |
| Blumenthal (2020)<br><br>*Blumenthal (2019)* | 41/41 | 52 | DASH | 18 sessions; 30-min weekly (0-3mo) then biweekly (3-6mo) to meet DASH guidelines. | 99.6% attendance to IP sessions | Y, IP | Weekly, 0-12 wks; biweekly (13-24 wks) | 39/39<br><br>Attention-Matched HE (PC) | Weekly 30-min sessions for 3-months, then biweekly calls on CVD health topics; no diet/ PA intervention |
| Brinkworth (2009) | 55/32 | 52 | VLC+HF+CR (Ketogenic) | VLC: (4% carb, 35% protein, 61% fat (<20g carb/d for 8-weeks, then <40g/day remainder); CR: 1433 – 1672kcal /day; Received diet plan and key foods supplied. Counselling with dietitian. | 58% INT compliance rate via meetings, food records vs. 65% control | Y, IP+ individual | Fortnightly (8 wks), then monthly (44 wks) | 52/33<br><br>Isocaloric AC | HC (46% total energy), LF (30%), CR (approx..1433 – 1672kcal /day). |

| | | | | | | | | | |
|---|---|---|---|---|---|---|---|---|---|
| | | | | | | | | | Same dietetic support. |
| Buchholz (2024)<br><br>*Brandt (2019)* | 20/13 | 12 | Atkins (Ketogenic) | Modified Atkins diet (MAD): ≤20g net carbs/day; high fat, moderate protein. Ketone testing at home. | 2/13 participants met moderate ketosis at ≥3 visits; 0/9 controls | Y, IP+phone +individual | Every 3-weeks | 18/9<br><br>AC | NIA diet guidelines; same dietitian education/ support every 3-weeks. |
| Chlebowski (2020) | 652/592 | 442 | LF | LF Diet: ≤20% energy intake from fat; increase fruit, vegetables and grains; 18 group sessions in Yr 1, then quarterly | Mean fat intake 24.2% at Yr 1; 29.8% at 8.5yr; +1.3 servings/ day fruit/veg | Y, IP+group | 18 sessions (1st year), and then 4 sessions per year | 954/860<br><br>PC | *Ad Libitum* Diet |
| Halyburton (2007) | 56/48 | 8 | LCHF (Ketogenic) | LC/HF diet: 4% carbohydrates, 35% protein, 61% fat. Reduced energy intake by 30%, 6000kJ (women) to 7000kJ (men) | Higher plasma ketones in LCHF group | Y | Every 2-weeks | 49/45<br><br>Isocaloric AC | HCLF: 46% carbohydrate, 30% fat, 24% protein. Isocaloric diet. Same support. |
| Hardman (2020) | 25/18 | 24 | MEDI | MEDI 6-week meal plans and recipes provided. Increase fruit, vegetables, grains, low red meat, moderate alcohol. EVOO supplied. | 50 – 70% Median diet adherence | N | None | 27/25<br><br>PC | Maintain usual lifestyle. |
| Horie (2016) | 40/38 | 52 | CR | 500kcal/day deficit (≥ 1200 kcal/day), ≥1g protein/kg, fibre-rich, fruit, vegetable, whole grain; 26 1-hr group nutritional counselling sessions on healthy diet + TAU. | 6 participants attended > 75% of sessions | Y, IP | Every 2-weeks | 40/37<br><br>PC | TAU with geriatrician every 2mo + general health advice, no diet support. |
| Hoscheidt (2021) | 44/41 | 4 | MEDI | 40-45% carbohydrate (low GI <55), 15-20% protein, 40% fat diet. Low saturated fat (<7% total energy), low salt, 1 wine unit/day allowed. Meals and food items prepared and delivered (personalised to basal metabolic rate). | <1 non-compliant meal/week (MEDI 0.82 ± 0.24; CTRL: 0.66 ± 0.22) | Y, | Weekly contact, food provision every 2-weeks | 43/43<br><br>Negative AC | Western-style diet; high saturated fight, high GI>70, high sodium. Meals supplied Same contact. |

| | | | | | | | | | |
|---|---|---|---|---|---|---|---|---|---|
| Jakobsen (2011)<br><br>*Zellner (2011)* | 13/11 | 3 | High protein | 3.0g protein/kg /day, 30% total energy from protein (double usual protein), isoenergetic replacement of carb with animal protein. Controlled food all supplied by department. Supervised lunches. | 96% adherence via daily urine analysis; Protein intake higher in HP group. | N | None | 13/12<br><br>Isocaloric AC | Usual protein (15% energy; 1.5g/kg/day). Isocaloric. Controlled food supplied. |
| James (2024) | 28/22 | 8 | IF | 14-hr nightly IF; 6 night/week, 1 day "off." No calories after 8pm, eat as usual within 10hr window. Adherence/barrier support. | 59.0% IF Ps fasted for full 14 hours for all 48 days. 73% completed all 8 check-ins. | Y, Phone | Weekly, 10-min call | 30/27<br><br>Attention-Matched HE | Health ed programme: 10-15 min videos/week. Same contact. |
| Jennings (2024) | 35/34 | 24 | MEDI | Personalised MEDI nutrition education, recipe ideas and 4 (2hr) group sessions on behaviour change. Intervention goal: improve MEDAS score by ≥3 points. Food vouchers (£30/week) /delivery. Web-based platform (LEAP2): personalised targets, self-assessment, tailored feedback. | 3.5/4 sessions attended; Mean MEDAS score +3.7; low website use 84% <1x/mo. 95% acceptability. | Y, Group | 4 Sessions (Week 0, 2, 4, 12) | 34/31<br><br>Minimal HE | 1hr group session at baseline. General advice for CVD risk. No other active support. |
| Keawtep (2024) | 23/21 | 12 | IF | IF 2 days/week – weeks 1-4 (75% EER); weeks 5-8 (50% EER); weeks 9-12 (25% EER), *Ad libitum* eating 5 days/week. | 88.8% adherence | Y, IP+Phone. +email. +SMS | Weekly contact; app reminders; IP 2-hr training | 23/20<br><br>PC | *Ad libitum* Diet |
| Knight (2016) | 85/70 | 24 | MEDI | MEDI diet based on Cretan pattern, personalised. EVOO, legumes, nuts, yogurt, tuna provided (providing 30-35% energy required); Personalised dietitian guidance, 14-point compliance checklist. | 92% compliance | Y, IP | Fortnightly | 81/67<br><br>PC | *Ad libitum* Diet and food vouchers. |
| Komulainen (2021)<br><br>*Komulainen (2010)* | 236/196 | 208 | Finnish Nutrition Guidelines | 5 individualised sessions year 1, then every 6-months; ≥400g/day of vegetables, fruit, and berries ≥2 servings fish/week (≥30g/day) ≥14g fiber/1000 kcal ≤10% from saturated fats; After 6-months, plant foods & less meat. | 84% adherence to full diet prescription | Y, IP+ individual+ group | 11 x 30-min IP 1:1 nutritionist sessions; 3 group sessions over 4-years | 236/201<br><br>Minimal HE | General advice on physical activity and diet at baseline only. |

| | | | | | | | | | |
|---|---|---|---|---|---|---|---|---|---|
| Krikorian (2012b) | 12/12 | 6 | VLC (Ketogenic) | ≤ 20g of carbohydrate/day; 5-10% energy from carbs, protein and fat *ad libitum;* no fruit. Self-monitoring and individualised baseline counselling. | NR, Mean 5.4mg/dl ketones confirmed adherence. | Y, IP+phone | Weekly, and counselling session at baseline. | 11/11<br><br>AC | HC diet; ≥50% energy carbs from fruit/veg. Matched contact. |
| Makris (2013) | 22/22 | 26 | Atkins LC (Ketogenic) | <20g carbs/day for 12 weeks, then increase by 5g/week for continued weight loss; 1200-1500 kcal/day women, 1500-1800kcal/day men; group behavioural sessions. | Urinary ketones higher in LC group wks 1-12, declined over time. | Y, group | Weekly | 25/25<br><br>AC | LFHC: 55% carbs, 30% fat, 15% protein. Isocaloric diet. Same contact. |
| Marseglia (2018) | 638/ 573 | 52 | MEDI | Personalised MEDI diet, country-specific diet guidelines for older adults. Nutritional counselling sessions and phone. Provision of free foods (e.g., EVOO, LF cheese, whole grains). Vitamin D supplement provided. | Mean INT adherence 65.9% (SD 11.0) vs. CTRL (52.7%) *p* <.001 | Y, IP+ phone+ individual | Monthly phone calls; IP counselling at baseline, 4, 8 months; ongoing access to dietitian | 641/571<br><br>Minimal HE | Habitual diet; Given leaflet on national dietary guidelines. No food, contact or support. |
| Martin (2007) | 24/23<br><br>12/12<br>CR<br><br>12/11<br>LCD | 24 | CR<br><br><br>LCD | **CR**: 25% CR; Food provided wk 1 – 12; 22 – 24. Self-selected CR diet week 13 – 21.<br>**LCD:** 890kCal/day liquid diet (Optifast) 15% weight loss, then maintenance; all food provided during same controlled phases. | CR: 10.4% weight loss at 6-mo; LCD: 13.9% weight loss at 6-mo | Y, IP+group | Weekly group meetings led by a psychologist | 12/11<br><br>Attention-Matched AC | Weight maintenance diet, no caloric restriction or weight loss; same support as CR and LCD |
| Martinez-Lapiscina (2013a)<br><br><br>*Martinez-Lapiscina (2013b)* | 704/390<br><br>351/224<br>MEDI+EVOO<br><br>352/166<br>MEDI+Nuts | 338 | MEDI<br>± EVOO or Nuts | **MEDI+EVOO:** MEDI diet with free 1L/week EVOO supplied. No CR or PA promoted.<br>**MEDI+Nuts:** MEDI diet with free 30g/day mixed nuts: 15g walnuts, 7.5g almonds, 7.5g hazelnuts. No CR or PA promoted.<br>All: Intensive quarterly dietitian counselling; annual adherence, family physician support. | Mean 14-point adherence score after 6-years<br>MEDI+EVOO: 10.8 (±2.7)<br>MEDI+Nuts: 10.1 (±4.0)<br>CTRL: 6.3 (±4.7) | Y, IP+ individual | Quarterly diet counselling; annual assessment | 352/132<br><br>AC | LF diet advice and leaflet; regular GP visits for UC; No food supplied or no dietitian advice |
| Napoli (2014) | 26/23 | 52 | CR | 500-750 kcal/day deficit with a targeted 10% reduction in body weight in the first 6-months, then maintenance; weekly weigh-ins, behavioural therapy. | Adherence 83% (IQR 79-89%) | Y, IP+group+ individual | Weekly | 27/23<br><br>Elimination HE | General diet advice. Instructed not to join any weight loss or PA program. |

| | | | | | | | | | |
|---|---|---|---|---|---|---|---|---|---|
| Rizvi (2024) | 60/60<br><br>30/30<br>IF<br><br>30/30<br>Custom Diet | 12 | IF<br><br><br><br><br>Custom Diet | **IF:** 16:8 time-restricted eating; fasting daily 9pm-1pm; 8-hr eating window (1-9pm), hydration guidance and structured meal recommendations<br>**Custom Diet:** Macronutrient-balanced plan based on 24-hr recall; Customised diet based on BMI and caloric needs | 100% retention; compliance monitored by weekly phone/ messages, no metrics reported. | Y, phone+ individual | Weekly | 30/30<br><br>PC | *Ad libitum* diet |
| Silver (2023)<br><br>*Leclerc (2019)*<br><br>*Grigolon (2020)* | 143/140 | 104 | CR | 25% CR from baseline energy intake. Meals provided for the first 27 days and rotated LF, MEDI, low GI diets. Individualised behavioural support by psychologist and RD. Macronutrient supplement provided. | 11.9% CR via doubly labelled water; 98% retention at 12-months; 83% at 24-months | Y, IP+ phone+ group+ individual | Weekly for first month, biweekly from month 2-12, monthly for remaining 12-months | 117/65<br><br>PC | *Ad libitum* diet; quarterly check-ins |
| Smith (2010) | 38/38 | 16 | DASH | DASH guidelines with no kcal targets. Increase fruits, vegetables, LF dairy; reduce saturated/total fat & cholesterol; increase whole grains, poultry, fish, nuts; reduce red meat/sugar. Diet counselling. | 92% class attendance in group sessions | Y, IP+group | Weekly | 43/42<br><br>PC | *Ad libitum* diet |
| Smith (2020) | 34/34 | 24 | DASH | Individualised DASH diet, no specific kcal target. DASH dietary modification promoting high intake of fruits, vegetables, whole grains, LF dairy, and reduced sodium, saturated fat, cholesterol, red/processed meat, sugar. Weekly 30-min dietary counselling | 100% completion; >85% diet session adherence. Food diary and FFQ no metrics reported. | Y, IP+ individual | Weekly 30-min for 12-weeks, then biweekly for 12-weeks (18 sessions) | 31/31<br><br>Attention-Matched HE | HE sessions; Weekly 30-min phone calls for 12-weeks, biweekly for 12-weeks, no diet/PA intervention. |
| Tussing-Humphreys (2022) | 148/128<br><br>73/64<br>MEDI<br><br>75/64<br>MEDI+CR | 34 | MEDI | Isocaloric MEDI diet, 1 oz almonds & 3 tbsp/day EVOO supplied; Mediterranean diet with olive oil and almonds supplementation; instructed to maintain baseline weight and PA. Weekly support. | Median attendance: 70.8% of sessions; MEDI adherence score 4.8-point increase. | Y, IP+group+ individual | Weekly 60-min group sessions and one 1:1 session | 37/35<br><br>AC | One 60-min 1:1 session. No diet/PA advice. Weekly newsletters. Offered 1:1 session & MEDI post-study. |

| | | | | | | | | | |
|---|---|---|---|---|---|---|---|---|---|
| Uchiyama-Tanaka (2024) | 14/11 | 52 | Low AGE Diet | **Diet:** Dietary guidance to reduce intake of advanced glycation end-products (AGEs); e.g., avoid fried/ processed, high-fructose foods; education sessions and materials **MAF:** 1 capsule macrophage-activating factor (MAF) 2x/day. | *NR* | Y, IP+ individual | Monthly (dietitian), 3× physician-led sessions at baseline/6/12 months | 15/14<br><br>TAU | Usual care; Continued outpatient rehabilitation. No diet advice; placebo capsules. |
| Valls-Pedret (2015) | 302/239<br><br>155/127 MEDI+EVOO<br><br>147/112 MEDI+Nuts | 208 | MEDI ± EVOO or Nuts | **MEDI+EVOO:** MEDI diet with free 1L/week EVOO supplied. No CR or PA promoted.<br>**MEDI+Nuts:** MEDI diet with free 30g/day mixed nuts: 15g walnuts, 7.5g almonds, 7.5g hazelnuts. No CR or PA promoted.<br>All: Intensive quarterly dietitian counselling; annual adherence, family physician support. | +49.6 ug/L in urinary biomarker in MEDI+EVOO and increase of 0.19% in plasma a-linolenic acid in MEDI+Nuts | Y, IP+ individual+ group | Quarterly individual and group sessions | 145/95<br><br>AC | LF diet; General advice to reduce fat intake; no Medi diet education; control leaflets; quarterly sessions added post hoc |
| Wardle (2000) | 120/105<br><br>59/52 LF<br><br>61/53 MEDI | 12 | LF<br><br>MEDI | **LF:** target <20% energy from fat, (mainly polyunsaturated); avoid saturated fats. Dietitian sessions and psychologist for CBT support; free fats/oils for compliance.<br>**MEDI:** target 30% energy from fat (mainly monounsaturated), increase intake of fruit, veg, oily fish, oily fish. Replace saturated fats with unsaturated; dietitian and psychologist for CBT support; free fats/oils for compliance. | 88% completed LF; 87% completed MEDI; Posttreatment analysis included completers that attended ≥4 sessions | Y, IP+group +individual | 8 sessions over 12 weeks | 56/50<br><br>WLC | No dietary advice, offered treatment after waiting period. Minimal contact at 6-week intervals. |
| Zhu (2024) | 30/27 | 12 | High-Fibre | ≥20g fibre/day via wheat rice (5.4 g fiber/day) from wheat rice + ≥15 g/day from fruit/veg on weekdays; ad libitum diet on weekends by advice to avoid high sugar/fat foods. | Fiber intake of the FD group increased from 7.89 to 20.24g day at 12-weeks ($p$ <.05) | Y, IP+Online | Initial IP instruction; Daily via WeChat | 30/25<br><br>WLC | *Ad libitum* Diet; Usual exercise habits. Received diet guidance after intervention. |

**Abbreviations:** AC: Active Control; ACSM: American College of Sports Medicine; AE: Aerobic Exercise; AGE: advanced glycation end-products; BM: Behavioural Management; CBT: Cognitive Behavioural Therapy; CD: Cognitive dysfunction; COMB-FP: COMpletely Balanced for Frailty Prevention; CR: Caloric Restriction; CT: Cognitive Training; CTRL: Control; CVD: Cardiovascular Disease; DASH: Dietary Approaches to Stop Hypertension; DHA: Docosahexaenoic acid; DRI: Daily recommended intake; EER: Estimated energy requirements; EPA: Eicosapentaenoic acid; EVOO: Extra-Virgin Olive Oil; FMI: Facility-based multidomain intervention; GI: Glycaemic index; GP: Doctor of general practice; HC: High carbohydrate; HE: Health Education; HF: High fat; HMI: Home-based multidomain intervention; HR: Heart rate; IF: Intermittent Fasting; INT: intervention; IP: In-Person; IPAQ:

International Physical Activity Questionnaire; IQR: Interquartile range; LC: Low carbohydrate; LCD: Low-calorie diet; LF: low fat; MA: Medical Adherence; MAD: Modified Atkins diet; MAF: macrophage-activating factor; MD: Multidomain; MEDAS: Mediterranean Diet Adherence Screener; MEDI: Mediterranean Diet; MF: Medical food; MH: Mental Health; MIND: Mediterranean-DASH Diet Intervention for Neurodegenerative Delay; MVPA: Moderate-Vigorous PA; N = No; NIA: National Institute on Aging; NNG: National Nutrition Guidelines; $NO_3^-$ = Nitrate ion; NR: Not Reported; NS: Nutritional supplement; PA: Physical Activity; PB: Peanut butter; PC: Passive Control; PCDS: Physio-cognitive decline syndrome; PUFA: Polyunsaturated Fatty Acids; RM: Repetition maximum; RO: Reality Orientation; RS: Rope skipping; SC: Standard Care; SD: Standard deviation; SE: Social Engagement; SM: Stress Management; SMS: Short Message Service; SRP: Skin roasted peanuts; SS: Social Support; ST: Strength Training; TAU: Treatment As Usual; UC: Usual care; UPF: Ultra processed foods; VLC: very low carbohydrate; VRM: Vascular Risk Management; WK: Week; WLC: Waitlist Control; Y = Yes.

**Supplementary Table S4c.** Single food intervention study characteristics (*n* = 31)

| Single Food Dietary Interventions (n = 31) | | | | | | | | | |
|---|---|---|---|---|---|---|---|---|---|
| Author (Year)[Ref] | Intervention sample size (T0/T1) | INT Duration (Weeks) | Food Category And Description | Intervention Dosage/Protocol | Intervention Adherence | Guided Nutrition Support (Y/N) | Frequency of Support | Control Sample Size (T0/T1) Condition | Control Description |
| Babateen (2022) | 47/37<br><br>16/10 High Nitrate<br><br>17/13 Medium Nitrate<br><br>14/14 Low Nitrate | 13 | Fruit & Vegetables: Beetroot | High Nitrate Beetroot: 2x70mL juice/day; 800mg $NO_3$- per day Medium Nitrate Beetroot: 1x70ml juice/day; 400mg $NO_3$- per day. Low Nitrate Beetroot: 1x 70mL every other day; 200 $NO_3$- per day). | Plasma $NO_3$- rose in active arms. | N | None | 15/13<br><br>Placebo | Nitrate-depleted beetroot juice (1x70ml shot every 2 days) |
| Boespflug (2017) | NR/8 | 16 | Fruit & Vegetables: Blueberry | Whole freeze-dried blueberry powder; 25g (2 x 12.5g sachet) blueberry powder/day equivalent to 148g (1 cup) of whole blueberry fruit. 399 kcal/100g. Avoid anthocyanin foods. | *62%* completed all procedures | N | None | NR/8<br><br>Placebo | Isocaloric Placebo. No anthocyanins. 389 kcal/100g. |
| Bøhn (2021) | 32/30 | 9 | Fruits & Vegetables: Bilberry/Grape | 330ml Pure bilberry + red grape juice (50/50) 2x /day = 660 ml. No sugar or additives. | 83% INT group drank full amount | N | None | 32/30<br><br>Placebo | Isocaloric placebo. No polyphenols. (330mL 2x/day). |
| Bookheimer (2013) | NR/15 | 4 | Fruits & Vegetables: Pomegranate | Daily consumption of 8oz (~240 mL/day) of commercial pomegranate juice (PomWonderful) for 4 weeks. Maintained low-polyphenol diet. | 78% had biomarker increase; 93% INT completion. | Y, IP+ individual | weekly check-ins with the dietitian | NR/13<br><br>Attention-Matched Placebo | Isocaloric 8oz matched beverage daily; low-polyphenol diet; identical contact. |
| Bowtell (2017) | 12/12 | 12 | Fruits & Vegetables: Blueberry | 30 mL blueberry concentrate/day (BlueberryActive; 387 mg | 100% 12/12 bottles returned | N | None | 14/14<br><br>Placebo | Isoenergetic placebo 30mL synthetic |

| | | | | | | | | | |
|---|---|---|---|---|---|---|---|---|---|
| | | | | anthocyanins) diluted to 240 mL with water; maintained habitual diet. | | | | | cordial matched sugar/calories; habitual diet. |
| Cardoso (2016) | 16/11 | 24 | Nuts & Seeds: Brazil Nut | 1 Brazil nut per day (~5g; avg. ~288.75 µg selenium/day) for 6 months; Nuts supplied in 2-month batches; maintain usual diet, avoid additional Brazil nuts or selenium foods. | 73% adequate compliance (≥85% nuts consumed) | N | None | 15/9<br><br>Elimination PC | Usual diet; no consumption of nut or supplement intake. |
| Chai (2019) | 20/17 | 12 | Fruit & Vegetables: Cherry | 480 mL/day (2x 240mL) of Montmorency tart cherry juice; 68mL concentrate + 412mL water daily for 12-weeks. | 94.2% compliance via returned unused beverage | N | None | 17/17<br><br>Placebo | 480 mL/day isocaloric placebo drink (Kool-Aid). No polyphenols. |
| Chan (2017) | 20/8 | 24 | Oils & Fats: Coconut Oil | 30 ml/day of cold pressed coconut oil for 2 weeks, then 60 ml/day for remainder; 2 divided doses, direct or with food; no diet advice. | 40% completed intervention; high dropout. | N | None | 20/14<br><br>Placebo | Placebo oil; same colour/ smell; same dosing. |
| Cheatham (2023) | 44/29 | 24 | Fruits & Vegetables: Blueberry | 35g/day (2 x 17.5g packets) of lyophilized wild blueberry powder daily, mixed with water or food. | 78.5% full adherence (returned packets). | N | None | 42/36<br><br>Placebo | 35 g/day isocaloric placebo powder; same instructions. |
| Coates (2020) | 77/63 | 12 | Nuts & Seeds: Almond | 43g/day of whole natural almonds; approximately 15% of EER, six days per week for 12 weeks. | Compliance >90% in intervention group (self-report) | Y, IP | Every 3-weeks by dietitian | 74/65 Isocaloric Attention-Matched AC | Energy-matched, nut- and seed-free, biscuits/potato chips; 15% daily EER, 6 d/wk for 12wks. Same contact. |
| Curtis (2024) | 11/9 | 24 | Fruits & Vegetables: Elderberry | 5 mL juice 3x/day (15 mL/day) for 6 months; 15.9mg cyanidin-glucose | 97% dosage compliance | N | None | 13/11<br><br>Placebo | 5mL placebo juice; same dosage. |

| | | | | | | | | | |
|---|---|---|---|---|---|---|---|---|---|
| Handajani (2020) | 30/30 Tempeh A<br><br>30/27 Tempeh B | 24 | Meat & Alternative Protein: Tempeh | Tempeh A: 100 g/day of Tempeh A for 6 months. Prepared as preferred (not deep-fried); no other soy/fermented food intake; Lower bacteria count. Food delivered.<br><br>Tempeh B: 100 g/day of Tempeh B for 6 months; Higher bacteria count. Same provision as above. | *NR* | N | None | 30/27<br><br>Isocaloric AC | 100 g/day low-protein biscuit (wheat flour, milk, sugar, salt, and vegetable fat) no soy. |
| Kamoun (2024) | 15/10 | 6 | Nuts & Seeds: Walnut | 15 g/day of walnut at 10:00 am daily for 6-weeks; supervised PA 3x/week (AE+ST) | *NR* | N | None | 13/10<br><br>AC | Supervised PA only; *ad libitum* diet, no walnut; no diet advice. |
| Kimble (2022) | 28/25 | 12 | Fruits & Vegetables: Cherry | 30 ml, 2x/day (60mL/day) concentrate diluted in water for 12-weeks; avoid other high-anthocyanin foods; usual diet; written instructions. | 100% dietary adherence (IPAQ + diary) | N | None | 28/25<br><br>Placebo | Isoenergetic 30mL placebo (Kool-Aid). Same dosage. |
| Krikorian (2009) | 5/5 | 12 | Fruits & Vegetables: Grape | 6-9 mL/kg/day of 100% Concord grape juice daily for 12-weeks (prescribed by body weight); divided into 3 doses with meals. | *NR* | N | None | 7/7<br><br>Placebo | Isoenergetic placebo beverage; no polyphenols. |
| Krikorian (2012a) | 10/10 | 16 | Fruits & Vegetables: Grape | 6-9 mL/kg/day, 3 divided doses of 100% Concord grape juice daily for 16-weeks (355 mL/day to 621 mL/day.) | *NR* | N | None | 11/11<br><br>Placebo | Isoenergetic placebo beverage; no polyphenols. |
| Krikorian (2022) | 15/13 | 12 | Fruits & Vegetables: Blueberry | 35g/day blueberry powder supplementation for 12-weeks: daily dosage equivalent to 0.5 cups of whole fruit; instructed to abstain from all berry fruits. | 86.7% adherence via supplement return/diet records | N | None | 18/14<br><br>Placebo | Maltodextrin powder; no active ingredients. Same instructions. |
| Krikorian (2023) | 17/15 | 12 | Fruits & Vegetables: Strawberry | 13 g/day of freeze-dried strawberry powder for 12-weeks. 36.8mg anthocyanins, ~130g whole fruit equivalent. | 88.2% adherence via supplement return/diet records | N | None | 17/15<br><br>Placebo | Identical cellulose/fibre no polyphenols |
| Lee (2017) | 5/5 | 24 | Fruits & Vegetables: Grape | 36 g of freeze-dried grape powder 2x/day (total of 72 g/day) diluted in water for 6-months. | *NR* | N | None | 5/5<br><br>Placebo | Isoenergetic-matched placebo |

| | | | | | | | | | |
|---|---|---|---|---|---|---|---|---|---|
| | | | | | | | | | powder, no polyphenols. |
| Mazza (2018) | 60/55 | 52 | Oils & Fats: EVOO | 20–30 g/day of EVOO for 12-months; all vegetable oils replaced with EVOO; oil supplied free; combined with MEDI diet instruction and personalised meal plans by dietitian (oral and written). | 30g ±12g EVOO consumption in intervention | Y, IP+phone | Baseline IP advice; follow-up call every 3-months | 144/55<br><br>AC | Standard MEDI diet, vegetable oil, no EVOO, same dietetic contact. |
| Miller (2018) | 20/19 | 13 | Fruits & Vegetables: Blueberry | 12g 2x/day for total 24g/day of freeze-dried blueberry powder for 90-days; equivalent to 1 cup/day of fresh blueberry; abstain from all berry fruits. | 99.2% compliance via weekly calls, calendars, and empty packet counts. | N | None | 20/19<br><br>Placebo | Isocaloric matched placebo powder; same protocol; no berry intake. |
| Miller (2021) | 21/19 | 13 | Fruits & Vegetables: Strawberry | 12g 2x/day for total 24 g/day of freeze-dried strawberry for 90-days; equivalent to 2 cups/day of fresh strawberry; abstain from all berries. | Mean compliance >97% via packet count, calendar, weekly calls. | N | None | 26/18<br><br>Placebo | Isocaloric matched placebo powder; same protocol; no berry intake. |
| Mirheidary (2019) | 48/36 | 4 | Fruits & Vegetables: Grape | 25g of maviz (seedless dried grapes), orally every morning for 4 weeks. Usual diet and PA. | 22.6 days mean consumption period of maviz | N | None | 24/17<br><br>PC | Usual diet; did not receive "maviz” |
| Parilli-Moser (2021) | 60/44<br><br>30/21 SRP<br><br>30/23 PB | 24 | Nuts & Seeds: Peanuts | SRP: 25g/day of skin roasted peanut (SRP) consumption; maintain habitual diet; avoid other nuts.<br><br>PB: 32 g/day of peanut butter (PB); maintain habitual diet; avoid other nuts. | Median compliance: 98.8% SRP, 98.4% PB | N | None | 30/19<br><br>AC | Macronutrient-matched 32 g/day control butter (peanut oil based, no polyphenols/ fibre) |
| Rakic (2022) | 48/43<br><br>22/19 1.5oz<br><br>26/24 3 oz | 24 | Nuts & Seeds: Almond | Participants consumed either 1) 1.5 oz (42 g) of almonds per day, or 2) 3 oz (84 g) of almonds per day for 24-weeks. Advised to maintain usual diet and weight; avoid other nuts. | *NR*, “adherence was high” by returned container/ compliance log | N | None | 20/17<br><br>Macro-nutrient-matched AC | Macronutrient-matched snack mix (100 g) of cereal mix, coconut, meat jerky, butter; no almonds. |
| Reeder (2022) | 40/32 | 12 | Nuts & Seeds: Peanut | 49 g/day of dry-roasted, skinless peanuts (equivalent to 290 calories | 83.9% adherence int; 95.8% | N | None | 35/29 | Peanut-free diet for the |

| | | | | | | | | | |
|---|---|---|---|---|---|---|---|---|---|
| | | | | per serving) for 12 weeks; usual diet; avoid other nuts. | adherence ctrl (abstained) | | | Elimination PC | same period; habitual diet. |
| Rodrigo-Gonzalo (2023) | 40/38 | 24 | Fruits & Vegetables: Raisin | 50 g/day of Málaga muscatel raisins daily for 24-weeks; maintain habitual diet; avoid other dried fruit. | 89.4% average adherence rate (± 21.1%) | Y, IP+group | Baseline 15-min nutritionist session | 40/36<br><br>PC | Did not receive the raisins; habitual diet. Same support. |
| Rutledge (2021) | 19/19 | 12 | Fruits & Vegetables: Blueberry | 24 g/day (12g 2x/day) of freeze-dried, lyophilized blueberry powder (Tifblue variety) for 3-months; equivalent to 1 cup of fresh blueberries per day. Dose taken morning and evening; maintain usual diet. | 99.2% adherence via supplement count and weekly phone check. | N | None | 19/19<br><br>Placebo | 24g of a seemingly identical, isocaloric control powder |
| Sala-Vila (2020) | 362/336 | 104 | Nuts & Seeds: Walnut | 30-60 g/day walnuts (15% daily energy), provided as daily sachets; maintain usual diet. Dietitian support. | 90% retention | Y, IP | Every 2 months | 346/321<br><br>Elimination PC | Instructed to abstain from walnuts, avoiding other nuts >2 servings/ week. No food supply. |
| Siddarth (2019) | 130/98 | 52 | Fruits & Vegetables: Pomegranate | 8 oz (236.5 mL) daily pomegranate juice for 12 months. Juice supplied. Maintain usual diet. | 89% compliance | N | None | 131/102<br><br>Placebo | 8 oz matched placebo drink; no polyphenols. |
| Wood (2023) | 35/27 | 12 | Fruits & Vegetables: Blueberry | 26 g/day of freeze-dried Wild Blueberry powder daily (302 mg anthocyanins equivalent to 178g fresh berries); maintain usual diet. | 100% by sachet count; SD = 0.05 | N | None | 31/27<br><br>Active Placebo | Placebo power; macronutrient, fibre, vit-C matched. No anthocyanins. |

**Abbreviations:** AC: Active Control; ACSM: American College of Sports Medicine; AE: Aerobic Exercise; AGE: advanced glycation end-products; BM: Behavioural Management; CBT: Cognitive Behavioural Therapy; CD: Cognitive dysfunction; COMB-FP: COMpletely Balanced for Frailty Prevention; CR: Caloric Restriction; CT: Cognitive Training; CTRL: Control; CVD: Cardiovascular Disease; DASH: Dietary Approaches to Stop Hypertension; DHA: Docosahexaenoic acid; DRI: Daily recommended intake; EER: Estimated energy requirements; EPA: Eicosapentaenoic acid; EVOO: Extra-Virgin Olive Oil; FMI: Facility-based multidomain intervention; GI: Glycaemic index; GP: Doctor of general practice; HC: High carbohydrate; HE: Health Education; HF: High fat; HMI: Home-based multidomain intervention; HR: Heart rate; IF: Intermittent Fasting; INT: intervention; IP: In-Person; IPAQ: International Physical Activity Questionnaire; IQR: Interquartile range; LC: Low carbohydrate; LCD: Low-calorie diet; LF: low fat; MA: Medical Adherence; MAD: Modified Atkins diet; MAF: macrophage-activating factor; MD: Multidomain; MEDAS: Mediterranean Diet Adherence Screener; MEDI: Mediterranean Diet; MF: Medical food; MH: Mental Health; MIND: Mediterranean-DASH Diet Intervention for Neurodegenerative Delay; MVPA: Moderate-Vigorous PA; N = No; NIA: National Institute on Aging; NNG: National Nutrition Guidelines; $NO_3^-$ = Nitrate ion; NR: Not Reported; NS: Nutritional supplement; PA: Physical Activity; PB: Peanut butter; PC: Passive Control; PCDS: Physio-cognitive decline syndrome; PUFA: Polyunsaturated Fatty Acids; RM: Repetition maximum; RO: Reality Orientation; RS: Rope skipping; SC: Standard Care; SD: Standard deviation; SE: Social Engagement; SM: Stress Management; SMS: Short Message Service; SRP: Skin roasted peanuts; SS: Social Support; ST: Strength Training; TAU: Treatment As Usual; UC: Usual care; UPF: Ultra processed foods; VLC: very low carbohydrate; VRM: Vascular Risk Management; WK: Week; WLC: Waitlist Control; Y = Yes.

**Supplementary Table S5.** Taxonomy of extracted cognitive outcome measures mapped to the six DSM-V neurocognitive domains

| Cognitive Measure | Abbreviation | DSM-V Neurocognitive Domains | | | | | | |
|---|---|---|---|---|---|---|---|---|
| | | **Executive Function** | **Complex Attention** | **Social Cognition** | **Learning & Memory** | **Language** | **Perceptual-Motor Function** | **Number of Domains** |
| Addenbrooke Cognitive Examination | ACE-III | | ✓ | | ✓ | ✓ | ✓ | 4 |
| Alzheimer's Disease Assessment Scale - Cognitive Subscale | ADAS-Cog | | ✓ | | ✓ | ✓ | ✓ | 4 |
| Anagrams | | ✓ | | | | | | 1 |
| Animal Naming Test | ANT | | | | ✓ | | | 1 |
| Attention Network Task | ANT | ✓ | ✓ | | | | ✓ | 3 |
| Auditory Consonant Trigram | ACT | ✓ | ✓ | | ✓ | | | 3 |
| Babcock Story Recall Test | BSRT | | | | ✓ | ✓ | | 2 |
| Bakan Vigilance Task | BVT | | ✓ | | ✓ | | ✓ | 3 |
| Behavior Rating Inventory of Executive Function | BRIEF | ✓ | | ✓ | | | | 2 |
| Benton Visual Retention Test | BVRT | | | | ✓ | | ✓ | 2 |
| Bond-Lader Visual Analogue Scale | VAS | | | ✓ | | | | 1 |
| Boston Naming Test (60-Item) | BNT | | | | ✓ | ✓ | | 2 |
| Boston Naming Test Short Form | BNT-15 | | | | ✓ | ✓ | | 2 |
| Brief Visuospatial Memory Test-Revised | BVMT-R | | | | ✓ | | ✓ | 2 |
| Buschke-Fuld Selective Reminding Task (Memory) | SRT | | | | ✓ | | | 1 |
| California Verbal Learning Test | CVLT | | | | ✓ | | | 1 |
| California Verbal Learning Test - **2nd Edition** | CVLT-II | | | | ✓ | | | 1 |
| Cambridge Cognitive Examination of Cambridge Examination for Mental Disorders of the Elderly | CAMCOG | ✓ | ✓ | | ✓ | ✓ | ✓ | 5 |

| | | | | | | | | |
|---|---|---|---|---|---|---|---|---|
| Cambridge Neuropsychological Test Automated Battery | CANTAB | ✓ | ✓ | ✓ | ✓ | | ✓ | 5 |
| Category Fluency Test | CFT | ✓ | | | ✓ | ✓ | | 3 |
| Central Nervous System Vital Signs Neurocognitive Battery | CNS Vital Signs | ✓ | ✓ | | ✓ | | ✓ | 4 |
| Clinical Dementia Rating - Global | CDR Global | ✓ | | | ✓ | | ✓ | 3 |
| Clinical Dementia Rating - Sum of Boxes | CDR-SB | ✓ | | | ✓ | | ✓ | 3 |
| Clock Drawing Task | CDT | ✓ | | | ✓ | | ✓ | 3 |
| Cognitive Failures Questionnaire | CFQ | ✓ | ✓ | | ✓ | ✓ | ✓ | 5 |
| CogState Brief Battery | CSBB | ✓ | ✓ | | ✓ | | | 3 |
| Colour Trail Test | CTT | ✓ | ✓ | | | | ✓ | 3 |
| COMPetency ASSessment | COMPASS | ✓ | | | ✓ | | | 2 |
| Concept Shifting Test (Condition A) | CST | ✓ | ✓ | | | | ✓ | 3 |
| Concept Shifting Test (Condition C) | CST | ✓ | ✓ | | | | ✓ | 3 |
| Conners' Continuous Performance Test-II | CPT-II | | ✓ | | | | | 1 |
| Consortium to Establish a Registery for Alzheimer's Disease | CERAD | ✓ | ✓ | | ✓ | ✓ | ✓ | 5 |
| Consortium to Establish a Registery for Alzheimer's Disease (Constructional Praxis Copy Task) | CERAD - CPC | | | | | | ✓ | 1 |
| Consortium to Establish a Registery for Alzheimer's Disease (Verbal) | CERAD - Verbal | | | | ✓ | ✓ | | 2 |
| Consortium to Establish a Registery for Alzheimer's Disease (Wordlist Memory) | CERAD-WL | | | | ✓ | ✓ | | 2 |
| Contextual Memory Test | CMT | | | | ✓ | | | 1 |
| Continuous Performance Task | CPT | | ✓ | | | | | 1 |
| Controlled Oral Word Production (Controlled Oral Word Association Test) | COWAT | ✓ | | | ✓ | ✓ | | 3 |

| | | | | | | | | |
|---|---|---|---|---|---|---|---|---|
| Corsi Blocks Task | | ✓ | | | | | ✓ | 2 |
| Creyos | | ✓ | ✓ | | ✓ | | ✓ | 4 |
| Delayed Matching to Sample | | | | | ✓ | | | 1 |
| Delis-Kaplan Executive Function System | D-KEFS | ✓ | ✓ | | | | | 2 |
| Delis-Kaplan Executive Function System (Colour Word Interference) | D-KEFS (CWI) | ✓ | ✓ | | | | | 2 |
| Delis-Kaplan Executive Function System (Tower Test Subtest) | D-KEFS (TT) | ✓ | ✓ | | | | | 2 |
| Detection Speed | | | ✓ | | | | | 1 |
| Digit Cancellation | | ✓ | ✓ | | | | | 2 |
| Digit Span Backward Subtest | DSB | ✓ | ✓ | | ✓ | | | 3 |
| Digit Span Forward Subtest | DSF | | ✓ | | ✓ | | | 2 |
| Digit Span Forward Subtest - Barcelona Test | | | ✓ | | ✓ | | | 2 |
| Digit Span Sequencing Subtest | DSS | ✓ | ✓ | | ✓ | | | 3 |
| Digital Vigilance Test | DVT | | ✓ | | | | ✓ | 2 |
| Dodrill Stroop Test | | ✓ | ✓ | | | | | 2 |
| Dot Counting Test | DCT | | ✓ | | | | | 1 |
| East Boston Story Delayed Recall | | | | | ✓ | | | 1 |
| East Boston Story Immediate Recall | | | | | ✓ | | | 1 |
| Eriksen Flanker Task - Choice Reaction Time | | ✓ | ✓ | | | | | 2 |
| Everyday Cognition-12 | E-Cog-12 | ✓ | ✓ | | ✓ | ✓ | ✓ | 5 |
| Everyday Memory Questionnaire | EMQ | | ✓ | | ✓ | | | 2 |
| Excluded Letter Fluency | ELF | ✓ | | | | ✓ | | 2 |
| Face-Name Association Test (immediate/delayed) | | | | | ✓ | | | 1 |
| Finger Tapping Test | FTT | | ✓ | | | | ✓ | 2 |
| Flanker Inhibitory Control and Attention Test | | ✓ | ✓ | | | | | 2 |

| | | | | | | | | |
|---|---|---|---|---|---|---|---|---|
| Formal Fluency (PMR, Spanish) | | ✓ | | | ✓ | ✓ | | 3 |
| Four-Choice Visual Reaction Time Test | | | ✓ | | | | ✓ | 2 |
| Free and Cued Selective Reminding Test | FCSRT | | | | ✓ | | | 1 |
| Go/No-Go Test | | ✓ | ✓ | | | | | 2 |
| Grammatical Reasoning Test | | | | | ✓ | ✓ | | 2 |
| Grooved Pegboard Test | | ✓ | ✓ | | | | ✓ | 3 |
| Groton Maze Chase Speed | GMLT | | ✓ | | | | ✓ | 2 |
| Groton Maze Learning Speed | GMLT | ✓ | ✓ | | ✓ | | | 3 |
| Groton Maze Recall Speed | GMLT | | ✓ | | ✓ | | | 2 |
| Hayling Test | | ✓ | | | | | | 1 |
| Hopkins Verbal Learning Test Revised | Hopkins VLT-R | | | | ✓ | | | 1 |
| Hopkins Verbal Learning Test-Revised (Delayed Memory) | Hopkins VLT | | | | ✓ | | | 1 |
| Hopkins Verbal Learning Test-Revised (Immediate Memory) | Hopkins VLT | | | | ✓ | | | 1 |
| Identification Speed (Reaction Time) | | | ✓ | | | | | 1 |
| Immediate and Delayed Recognition | | | | | ✓ | | | 1 |
| Inspection Time Task | IT | | ✓ | | | | | 1 |
| Intrinsic/Phasic Alertness | | | ✓ | | | | | 1 |
| IQCODE | | ✓ | | | ✓ | | ✓ | 3 |
| Jessen questions **https://pmc.ncbi.nlm.nih.gov/articles/PMC4317324/** | SCD | ✓ | ✓ | | ✓ | ✓ | ✓ | 5 |
| Korean-Mini Mental State Examination | K-MMSE-2 | | ✓ | | ✓ | ✓ | ✓ | 4 |
| Letter Digit Substitution Test | LDST | ✓ | ✓ | | | | ✓ | 3 |
| Logical Memory Test (Immediate & Delayed Recall) | LM | | | | ✓ | | | 1 |

| | | | | | | | | |
|---|---|---|---|---|---|---|---|---|
| Medical College of Georgia Complex Figure Test | MCG | | | | ✓ | | ✓ | 2 |
| Memory Performance Index | MPI | | | | ✓ | | | 1 |
| Mental Rotation Task | | ✓ | | | | | ✓ | 2 |
| MicroCog Assessment of Cognitive Functioning | MicroCog | ✓ | ✓ | | ✓ | | ✓ | 4 |
| Mild Cognitive Impairment Screen | MCIS | ✓ | ✓ | | ✓ | | ✓ | 4 |
| Mini-Mental State Examination | MMSE | | ✓ | | ✓ | ✓ | ✓ | 4 |
| Mini-Mental State Examination (2nd Edition) - Expanded Version | MMSE-2-EV | | ✓ | | ✓ | ✓ | ✓ | 4 |
| Minimum Data Set – Home Care Instrument | MDS-HCI | ✓ | | | ✓ | | | 2 |
| Modified Boston Naming test | mBNT | | | | ✓ | ✓ | | 2 |
| Modified Clinical Dementia Rating - Sum of Boxes | mCDR-SB | ✓ | | | ✓ | | ✓ | 3 |
| Modified Mini-Mental State Examination | 3MS | | ✓ | | ✓ | ✓ | ✓ | 4 |
| Modified RBANS Delayed Recall | mRBANS-DR | | | | ✓ | | | 1 |
| Modified RBANS Immediate Recall | mRBANS-IR | | | | ✓ | | | 1 |
| Modified RBANS Verbal List Recall | mRBANS-VR | | | | ✓ | | | 1 |
| Modified Wisconsin Card Sorting Test - 48 Cards | M-WCST | ✓ | | | | | | 1 |
| Montreal Cognitive Assessment | MoCA | ✓ | ✓ | | ✓ | ✓ | ✓ | 5 |
| Mood and Feelings Questionnaire (Frequency of Forgetting) | MFQ-FF | | | | ✓ | | | 1 |
| Mood and Feelings Questionnaire (Seriousness of Forgetting) | MFQ-SF | | | | ✓ | | | 1 |
| Motor Screening Task | | | ✓ | | | | ✓ | 2 |
| Multifactorial Memory Questionnaire | MMQ | | | | ✓ | | | 1 |
| Multifactorial Memory Questionnaire - Memory Toolbox Task | MMQ-MTT | | | | ✓ | | | 1 |

| | | | | | | | | |
|---|---|---|---|---|---|---|---|---|
| Multilingual Naming Test | MINT | | | | ✓ | ✓ | | 2 |
| MYB Cognitive Test Battery | MYB | ✓ | ✓ | | ✓ | | ✓ | 4 |
| N-Back Test | | ✓ | ✓ | | | | | 2 |
| Neuropsychological Test Battery | NTB | ✓ | ✓ | | ✓ | ✓ | ✓ | 5 |
| Nishimura Mental State Scale | NM Scale | | | ✓ | ✓ | ✓ | ✓ | 4 |
| Number-letter computer task | | ✓ | ✓ | | | | | 2 |
| One Back Test Speed | | ✓ | ✓ | | | | | 2 |
| One Touch Stockings of Cambridge | OTS | ✓ | | | | | ✓ | 2 |
| Oral Symbol Digit Modality Test | SDMT | | ✓ | | | | ✓ | 2 |
| Paired Associate Learning | PAL | | | | ✓ | | ✓ | 2 |
| Pattern Comparison Test | | | ✓ | | | | | 1 |
| Pfeffer Functional Activities Questionnaire | FAQ | ✓ | ✓ | | ✓ | ✓ | ✓ | 5 |
| Phonemic fluency test | | ✓ | | | ✓ | ✓ | | 3 |
| Porteus Maze Test | PMT | ✓ | | | | | ✓ | 2 |
| Post Graduate Institute Memory Scale | PGI-MS | | ✓ | | ✓ | | | 2 |
| Prospective and Retrospective Memory Questionnaire | PRMQ | | | | ✓ | | | 1 |
| Rapid Visual Information Processing | RVP | | ✓ | | | | | 1 |
| Rapid Visual Processing | RVP | | ✓ | | | | | 1 |
| Rappel Libre/Rappel Indice (RL/RI-16) Test | RL/RI16 | | | | ✓ | | | 1 |
| Reaction Time Test | RTI | | ✓ | | | | ✓ | 2 |
| Reading the Mind in the Eyes (Revised) | | | | ✓ | | | | 1 |
| Repeatable Battery for the Assessment of Neuropsychological Status | RBANS | | ✓ | | ✓ | ✓ | ✓ | 4 |
| Repeated Acquisition Test | | | ✓ | | ✓ | | ✓ | 3 |

| | | | | | | | | |
|---|---|---|---|---|---|---|---|---|
| Rey Auditory Verbal Learning Test | RAVLT | | | | ✓ | | | 1 |
| Rey Auditory Verbal Learning Test (Delayed Recall) | RAVLT - DR | | | | ✓ | | | 1 |
| Rey-Osterreth Complex Figure Test (Delayed Memory) | ROCF-DM | | | | ✓ | | ✓ | 2 |
| Ruff 2 & 7 Test | | | ✓ | | | | ✓ | 2 |
| Scanning Visual Vigilance Test | | | ✓ | | | | ✓ | 2 |
| Sea Hero Quest Test | | | | | | | ✓ | 1 |
| Self-ordered Pointing Task | SOPT | ✓ | | | ✓ | | ✓ | 3 |
| Semantic Fluency | | ✓ | | | ✓ | ✓ | | 3 |
| Semantic Fluency (Spanish) | | ✓ | | | ✓ | ✓ | | 3 |
| Serial 3s and 7s Subtraction Tasks | | ✓ | ✓ | | | | | 2 |
| Shopping List Recall Accuracy | | | | | ✓ | | | 1 |
| Simple and Choice Reaction Times | SCRT | ✓ | ✓ | | | | ✓ | 3 |
| Simple Reaction Time | | | ✓ | | ✓ | | ✓ | 3 |
| Spain-Complutense Verbal Learning Test | TAVEC | | | | ✓ | | | 1 |
| Spanish version of the California Verbal Learning Test | CVLT-S | | | | ✓ | | | 1 |
| Spatial Paired Associate Learning Test | PAL | | | | ✓ | | ✓ | 2 |
| Spatial Span Forward | | ✓ | | | | | ✓ | 2 |
| Spatial Span Reverse | | ✓ | | | | | ✓ | 2 |
| Spatial Working Memory | SWM | ✓ | | | | | ✓ | 2 |
| Speed of Processing - Inspection Time | | | ✓ | | | | ✓ | 2 |
| Stroop Colour Subtest | SCS | ✓ | ✓ | | | | | 2 |
| Stroop Colour Word Interference Score | SCWT | ✓ | ✓ | | | | | 2 |
| Stroop Word Subtest | SWS | ✓ | ✓ | | | | | 2 |
| Supermarket Trolley Task | | ✓ | ✓ | | ✓ | | ✓ | 4 |

| | | | | | | | | |
|---|---|---|---|---|---|---|---|---|
| Swinburne University Computerised Cognitive Assessment Battery | SUCCAB | ✓ | ✓ | | ✓ | | ✓ | 4 |
| Symbol Digit Modalities Test | SDMT | | ✓ | | | | ✓ | 2 |
| Taylor Complex Figure test | TCFT | | | | ✓ | | ✓ | 2 |
| Telephone Montreal Cognitive Assessment | T-MoCA | ✓ | ✓ | | ✓ | ✓ | ✓ | 5 |
| Tower of London Task | | ✓ | | | | | | 1 |
| Trail Making Test Part A | TMT-A | | ✓ | | | | ✓ | 2 |
| Trail Making Test Part B | TMT-B | ✓ | ✓ | | | | ✓ | 3 |
| Verbal Fluency - Letter (Study Generated) | | | | | ✓ | | | 1 |
| Verbal Free Recall Task | | | | | ✓ | | | 1 |
| Verbal Paired Associates | VPA | | | | ✓ | | | 1 |
| Verbal Recognition Memory | VRM | | | | ✓ | | | 1 |
| Verbal Selective Reminding Test | VSRT | | | | ✓ | | | 1 |
| Virtual Morris Water Maze | vMWM | | | | ✓ | | ✓ | 2 |
| Visospatial Problem Solving | | ✓ | | | | | ✓ | 2 |
| Visual Memory Task - Taxi Driver | | | | | ✓ | | ✓ | 2 |
| Visual Object and Spatial Perception Battery | VOSP | | | | | | ✓ | 1 |
| Visual Paired Associates (Immediate & Delayed Recall) | VPA | | | | ✓ | | ✓ | 2 |
| WAIS-III Block Design | WAIS-III-BD | | ✓ | | | | ✓ | 2 |
| WAIS-III Digit Span | WAIS-III-DSp | ✓ | ✓ | | ✓ | | | 3 |
| WAIS-III Digital Symbol | WAIS-III-Dsy | ✓ | ✓ | | | | ✓ | 3 |
| WAIS-III Letter-Number sequencing | WAIS-III-LNS | ✓ | ✓ | | ✓ | | | 3 |
| WAIS-III Spatial Span | WAIS-III-SpS | ✓ | ✓ | | | | | 2 |
| WAIS-III Symbol Search | WAIS-III-SyS | | ✓ | | | | ✓ | 2 |
| WAIS-IV Letter-Number Sequencing | WAIS-IV-LNS | ✓ | ✓ | | ✓ | | | 3 |
| WAIS-IV Perceptual Reasoning Matrix | WAIS-IV-PR | | | | | | ✓ | 1 |

| | | | | | | | | |
|---|---|---|---|---|---|---|---|---|
| WAIS-R Digit Symbol Substitution Test | DSST | ✓ | ✓ | | | | ✓ | 3 |
| Wechsler Memory Scale | WMS | ✓ | | | ✓ | | | 2 |
| Wechsler Memory Scale - Digit Span | WMS DS | ✓ | | | ✓ | | | 2 |
| Wechsler Memory Scale - Verbal Memory Task | WMS VM | ✓ | | | ✓ | | | 2 |
| Wechsler Memory Scale - Visual Reproduction | WMS-IV VR | ✓ | | | ✓ | | | 2 |
| Wechsler Memory Scale Fourth Edition | WMS-IV | ✓ | | | ✓ | | | 2 |
| Wechsler Memory Scale Fourth Edition - Logical Memory | WMS-IV LM | ✓ | | | ✓ | | | 2 |
| Wechsler Memory Scale-Revised | WMS-R | ✓ | | | ✓ | | | 2 |
| Wechsler Test of Adult Reading | WTAR | ✓ | | | ✓ | ✓ | | 3 |
| Wisconsin Card Sorting Test-64 | WCST-64 | ✓ | | | | | | 1 |
| Word List Memory Test (Learning & Delayed Recall) | WLMT | | | | ✓ | | | 1 |
| Word List Recall Test | | | | | ✓ | | | 1 |
| Word List Recognition Test | | | | | ✓ | | | 1 |
| **Cognitive Measure** | **Abbreviation** | **Executive Function** | **Complex Attention** | **Social Cognition** | **Learning & Memory** | **Language** | **Perceptual-Motor Function** | **Number of Domains** |

**Supplementary Table S6a.** Between-group differences in cognitive outcomes at post-intervention in multi-domain interventions (n = 30)

| **Multi-Domain Dietary Intervention Studies (n = 30)** | | | | | | **+ = Favours intervention**<br>**— = Favours control**<br>***ns* = No significant effect/difference in CF**<br><br>***=p<0.05**<br>**†=p<0.01**<br>**‡=p<0.001** |
|---|---|---|---|---|---|---|
| Author (Year) | Cognitive Status | INT Type | Global Cognitive Function | Single-Domain Cognition | Brain Imaging | Blood Biomarkers |
| Andrieu (2017)<br>*Chhetri (2018)* | At-Risk | NNG+PA, CT, GP, PUFA | *ns* MMSE<br>*ns* CDR-SB<br>*ns* composite scores | +MMSE Orientation*<br>*ns* FCSRT<br>*ns* DSST<br>*ns* Category Naming Test<br>*ns* Lexical Fluency Test (COWAT)<br>*ns* TMT-A<br>*ns* TMT-B<br>*ns* Memory functioning | +amyloid-positive PET scans showed less cognitive decline‡ | +DHA‡<br>+EPA‡ |
| Blumenthal (2020)<br>*Blumenthal (2019)* | MCI | DASH+PA | +CDR-SB† | +Composite executive function†<br>*ns* Memory<br>*ns* Language / verbal fluency | *NR* | *ns* TC, LDL-C, HDL-C, VLDL-C<br>*ns* insulin<br>*ns* glucose<br>*ns* triglycerides |
| Brodaty (2025) | Normal | MEDI+PA, CT, CBT | +Global cognitive composite‡ | +Complex attention‡<br>+Executive function‡<br>+Learning and memory‡ | *NR* | *NR* |
| Chatterjee (2022) | Normal | MEDI+PA + CT<br>MEDI + CT only | *NR* | +PGI-MS Total‡ (MEDI+PA+CT)<br>+Mental balance†<br>+Attention/concentration‡ (MEDI+PA+CT)<br>*ns* Immediate recall<br>+Visual retention*(MEDI+PA+CT) | *NR* | *NR* |
| Chou (2022) | Normal | MEDI+CT | +MoCA†<br>*ns* CFQ | *NR* | *NR* | *NR* |
| Han (2023) | Normal | NNG+PA, SM | +MMSE* | *NR* | *NR* | *NR* |
| Hardman (2020) | Normal | MEDI+PA | *ns* SUCCAB composite | +Spatial working memory*<br>*ns* Simple choice reaction time<br>*ns* Immediate recognition memory<br>*ns* Delayed recognition memory | *NR* | +Vitamin B-12*<br>*ns* TC<br>*ns* Fasting Glucose<br>*ns* hs-CRP |

| | | | | | | |
|---|---|---|---|---|---|---|
| | | | | *ns* Stroop Colour Word<br>*ns* Contextual memory | | *ns* Vitamin B-6, D<br>*ns* HbA1c<br>*ns* IGF-I<br>*ns* BDNF<br>*ns* homocysteine |
| Jennings (2024) | Normal | MEDI+PA | *NR* NTB | *NR* Verbal memory<br>*NR* Processing speed<br>*NR* Executive function | *NR* | *NR* |
| Keawtep (2024) | Normal | IF+PA, CT | *ns* MoCA | +Logical memory*<br>+TMT B-A*<br>*ns* Stroop Colour Word<br>*ns* Verbal fluency<br>*ns* Digit Span | *NR* | +BDNF*<br>+Total-C*<br>+adiponectin*<br>+Insulin*<br>+HOMA-IR*<br>*ns* IL-6<br>*ns* Triglycerides<br>*ns* Glucose |
| Koblinsky (2022) | Normal | MEDI/<br>DASH+PA | *NR* | *ns* RAVLT | *ns* MRI hippocampal volume | +HbA1c*<br>*ns* Vitamin K* |
| Komulainen (2021)<br><br>*Komulainen (2010)* | Normal | NNG+PA | *NR* MMSE | *ns* Verbal fluency<br>*ns* Modified Boston Naming<br>*ns* Word list memory<br>*ns* Constructional praxis<br>*ns* Word list recall<br>*ns* Word list recognition<br>*NR* CDT (2010) | *NR* | *NR* Total-C<br>*NR* LDL-C<br>*NR* HDL-C<br>*NR* Triglycerides<br>*NR* Glucose |
| Lee (2023) | MCI/AD | MIND+PA,<br>CT, GP,<br>VRM, NS,<br>Motivation | +RBANS‡<br>+MMSE*<br>*ns* CDR-SB | *ns* Immediate memory<br>*ns* Delayed memory<br>+Visuoconstruction‡<br>*ns* Language<br>*ns* Attention | *NR* EEG | *ns* BDNF<br>*ns* cortisol |
| Liang (2021) | Normal | NNG+PA,<br>CT, HE | +MoCA†(at-risk) | *ns* Visuospatial<br>*ns* Naming<br>+Concentration‡(at-risk)<br>*ns* Language<br>*ns* Abstract thinking<br>+Delayed recall*(at-risk)<br>*ns* Orientation | *NR* | *NR* |
| Martin (2007) | Normal | CR+PA | *NR* | *ns* RAVLT<br>*ns* ACT | *NR* | *NR* |

| | | | | | | |
|---|---|---|---|---|---|---|
| | | | | *ns* BVRT<br>*ns* CPT | | |
| Masley (2008) | Normal | HFD-LSF+ PA, SM | *NR* | *ns* CNS Mental Speed<br>*ns* CNS Reaction Time<br>*ns* CNS Attention<br>*ns* CNS Cognitive Flexibility | *NR* | +TC:HDL Ratio*<br>*NR* Total-C<br>*NR* HDL-C |
| McMaster (2020) | MCI | MEDI+PA, CT | +Composite*<br>*ns* ADAS-Cog | *ns* SDMT<br>*ns* Category Fluency<br>*ns* TMT-B | *NR* | *NR* |
| Mendoza-Ruvalcaba (2015) | Normal | NNG+PA, CT,RO | *NR* | +Processing Speed†<br>*ns* working memory | *NR* | *NR* |
| Moon (2021) | At-risk | MIND+PA, CT, VRM, SE, Motivation | +RBANS†<br>*ns* MMSE<br>*ns* CDR-SB | *ns* RBANS Immediate Memory<br>+RBANS Visuoconstruction†<br>*ns* RBANS Language<br>+RBANS Attention*<br>+RBANS Delayed memory* | *NR* | +BDNF*<br>+Cortisol*<br>+C-Peptide*<br>*ns* NfL<br>*ns* Total-C, LDL-C, HDL-C<br>*ns* Triglycerides<br>*ns* Lipoprotein<br>*ns* Fasting Glucose<br>*ns* HbA1c<br>*ns* Vitamin B-12<br>*ns* Folate<br>*ns* Vitamin D<br>*ns* Homocysteine<br>*ns* YKL-40 |
| Nakazeko (2023) | Normal, MCI | COMB-FP+PA | *NR* | +MPI Memory* | *NR* | *NR* |
| Napoli (2014) | Normal | CR+PA | +3MS (MMSE)‡ | +TMT-A‡<br>+TMT-B*<br>+Word list fluency‡ | *NR* | *ns* hs-CRP<br>*ns* Insulin Sensitivity Index<br>*ns* IGF-1 |
| Ngandu (2015)<br><br>*Rosenberg (2018)* | At-risk | NNG+PA, CT, VRM | +NTB Total Score* | +NTB Executive Function*<br>+NTB Processing Speed*<br>*ns* NTB Memory score | *NR* | *ns* Total-C, LDL-C, HDL-C<br>*ns* Fasting glucose<br>*ns* Glucose tolerance test |
| Ornish (2024) | MCI, early AD | Vegan+PA, SM, HE, SS, NS | +CDR-Global*<br>+CDR-SB*<br>*ns* ADAS-Cog | *NR* | *NR* | +Aβ42/40 ratio†<br>*ns* pTau181<br>+Insulin*<br>+B-HB*<br>+LDL-C‡<br>*ns* GFAP |

| | | | | | | |
|---|---|---|---|---|---|---|
| | | | | | | *ns* CRP<br>*ns* Serum amyloid<br>+GlycA[†]<br>*ns* Telomere length |
| Roach (2023) | MCI, early AD | MIND+PA, CT, SM, SS, Sleep, MA | *NR* MoCA | +MPI Memory Index* | *NR* | *NR* |
| Sakurai (2024) | MCI | NNG+PA, CT, VRM | *ns* MMSE<br>*ns* Composite | *ns* FCSRT<br>*ns* Logical memory<br>*ns* DSST<br>*ns* TMT-A, TMT-B<br>*ns* Digit span test<br>*ns* Letter word fluency test | *NR* | *NR* |
| Smith (2010) | Normal | DASH+PA, BM | +EFML Composite* | +Psychomotor speed*<br>*ns* DSST<br>+Ruff 2&7[†]<br>+TMT B-A*<br>*ns* VPA<br>*ns* COWAT<br>+Stroop*<br>*ns* Digit Span<br>*ns* Verbal fluency (animal naming) | *NR* | *NR* |
| Smith (2020) | MCI | DASH+PA | *NR* | *NR* Executive Function*<br>*NR* Memory<br>*NR* Language | *NR* | +Metabolic Composite*<br>*ns* IL-6<br>*ns* hs-CRP<br>*ns* BDNF<br>+HOMA-IR*<br>*ns* IGF-1<br>*ns* Leptin<br>*ns* VEGF |
| Thunborg (2024) | MCI | NNG+PA, CT, VRM, SS | +CDR-SB*<br>*ns* CDR-Global | *NR* | *NR* | *NR* HDL-C |
| Tussing-Humphreys (2022) | MCI | MEDI+CR +PA | *NR* | *NR* AIP Composite<br>*NR* EF Composite<br>*NR* LMR Composite | *NR* | +HOMA-IR*<br>+Fasting Insulin*<br>*ns* HbA1c<br>*ns* Total-C, LDL-C, HDL-C<br>*ns* Triglycerides<br>*ns* hs-CRP |

| | | | | | | |
|---|---|---|---|---|---|---|
| | | | | | | *ns* Fasting glucose |
| Zhu (2024) | Normal | HFD+PA | *NR* | +PRMQ Memory Total[†]<br>*ns* BRIEF Executive function | *NR* | *NR* |
| Zulke (2024) | At-risk | NNG+PA, CT, VRM, SS, MA, MH | *ns* Global cognitive composite | *NR* | *NR* | *NR* |

Abbreviations: 3MS: Modified Mini-Mental State Examination; Aβ42/40: Amyloid-beta 42 to 40 ratio; AD: Alzheimer's Disease; ACT: Auditory Consonant Trigrams; ADAS-Cog: Alzheimer's Disease Assessment Scale – Cognitive; AIP: Attention, Information, & Processing; BDNF: Brain-Derived Neurotrophic Factor; B-HB: Beta-Hydroxybutyrate; BM: Behavioural Management; BRIEF: Behaviour Rating Inventory of Executive Function; BVRT: Benton Visual Retention Test; C-Peptide: Connecting peptide; CBT: Cognitive Behavioural Therapy; CDR-Global: Clinical Dementia Rating – Global Score; CDR-SB: Clinical Dementia Rating – Sum of Boxes; CDT: Clock Drawing Task/Test; CERAD-TS: Consortium to Establish a Registry for Alzheimer's Disease – Total Score; CFQ: Cognitive Failures Questionnaire; CNS: Central Nervous System; COMB-FP: Completely Balanced for Frailty Prevention Diet; COWAT: Controlled Oral Word Association Test; CPT: Continuous Performance Test; CR: Caloric Restriction; CRP / hs-CRP: (High-sensitivity) C-Reactive Protein; CT: Cognitive Training; DASH: Dietary Approaches to Stop Hypertension; DHA: docosahexaenoic acid; DSST: Digit Symbol Substitution Test; EEG: Electroencephalogram; EF Composite: Executive Function Composite; EFML: Executive Function / Memory Learning Composite; EPA: ecosapentaenoic acid; FCSRT: Free and Cued Selective Reminding Test; GFAP: Glial Fibrillary Acidic Protein; GlycA: Glycoprotein Acetylation; HbA1c: Haemoglobin A1c; HDL-C: High-Density Lipoprotein Cholesterol; HFD: High Fibre Diet; HOMA-IR: Homeostatic Model Assessment of Insulin Resistance; IF: Intermittent Fasting; IGF-1: Insulin-like Growth Factor I; IL-6: Interleukin 6; LDL-C: Low-Density Lipoprotein Cholesterol; LMR Composite: Learning-Memory Recognition Composite; LSF: Low Saturated Fat; MA: Medical Adherence; MCI: Mild Cognitive Impairment; MEDI: Mediterranean Diet; MH: Mental Health; MIND: Mediterranean-DASH Intervention for Neurodegenerative Delay Diet; MMSE: Mini-Mental State Examination; MoCA: Montreal Cognitive Assessment; MPI: Memory Performance Index; NfL: Neurofilament Light Chain; NNG: National Nutrition Guidelines; ns: non-significant; NS: Nutrition Supplement; NTB: Neuropsychological Test Battery; PA: Physical Activity; pTau181: Phosphorylated Tau - 181; NNG: National Nutrition Guidelines; NR: Not Reported; NS: Nutrition Supplement; PRMQ: Prospective and Retrospective Memory Questionnaire; PUFA: Polyunsaturated Fatty Acids; RBANS: Repeatable Battery for the Assessment of Neuropsychological Status; RO: Reality Orientation; Ruff 2&7: Ruff 2 & 7 Selective Attention Test; SCWT: Stroop Colour and Word Test; SE: Social Engagement; SM: Stress Management; SS: Social Support; SUCCAB: Swinburne University Computerized Cognitive Assessment Battery; TC: Total Cholesterol; TC:HDL Ratio: Total Cholesterol to HDL-C Ratio; TMT-A/B: Trail Making Test Part A / B; TMT B-A: Trail Making Test (B minus A); Triglycerides: Triglycerides; VEGF: Vascular Endothelial Growth Factor; VLDL-C: Very Low-Density Lipoprotein Cholesterol ; VRM: Vascular Risk Management; VPA: Visual Paired Associates; YKL-40: chitinase-3-like protein biomarker, a glycoprotein linked to inflammation and neurodegeneration.

**Supplementary Table S6b.** Between-group differences in cognitive outcomes as post-intervention in total diet interventions (*n* = 33)

| **Total Diet Interventions (n = 33)** | | | | | | **+ = Favours intervention**<br>**— = Favours control**<br>***ns* = No significant effect/difference in CF**<br><br>***=p<0.05**<br>**†=p<0.01**<br>**‡=p<0.001** |
|---|---|---|---|---|---|---|
| Author (year) | Cognitive Status | INT Type | Global Cognitive Function | Single-Domain Cognition | Brain Imaging | Blood Biomarkers |
| Arjmand (2022) | Normal | MIND+CR | *NR* | +TMT-A†<br>*ns* TMT-B<br>+FDST*<br>+BDST*<br>+LNST‡<br>+SDMT†<br>+AVLT‡<br>*ns* Stroop | *ns* MRI Cortical thickness<br>*ns* MRI Cortical volume<br>+MRI SA Inf. frontal gyrus*<br>*ns* MRI Cerebellum<br>*ns* MRI Cortical cortex | +Homocysteine†<br>*ns* Amyloid Beta<br>*ns* BDNF |
| Barnes (2023) | Normal | MIND+CR | *ns* Global Cognition composite | *ns* Episodic memory<br>*ns* Semantic memory<br>*ns* Executive function<br>*ns* Perceptual speed | *ns* MRI White matter volume<br>*ns* MRI Hippocampal volume<br>*ns* MRI Total brain volume | *ns* Lutein<br>*ns* Zeaxanthin<br>*ns* Alpha Carotene<br>*ns* Beta Carotene |
| Bartholomew (2021) | Normal | IF | *ns* MicroCog Composite | *NR* | *NR* | +HOMA-IR†<br>+Fasting insulin‡<br>*ns* BDNF<br>*ns* Total-C, LDL-C, VLDL-C<br>+HDL-C*<br>+Fasting Glucose*<br>*ns* Triglycerides<br>*ns* Apolipoprotein<br>*ns* hs-CRP |
| Blondal (2022) | MCI | NNG | +MMSE‡ | *NR* | *NR* | *NR* |
| Blumenthal (2020)<br>*Blumenthal (2019)* | MCI | DASH | *ns* CDR-SB | *ns* Executive function<br>*ns* Memory<br>*ns* Language, verbal fluency | *NR* | +Total-C*, LDL-C*<br>*ns* Triglycerides<br>*ns* HDL-C, VLDL-C<br>*ns* Insulin<br>*ns* Glucose |
| Brinkworth (2009) | Normal | KETO | *NR* | *NR* Working memory | *NR* | *ns* Insulin |

| | | | | | | |
|---|---|---|---|---|---|---|
| | | | | *NR* Processing speed | | *ns* Glucose |
| Buchholz (2024)<br>*Brandt (2019)* | MCI, early AD | KETO | *ns* MMSE | *ns* Memory composite | *NR* | +Lipids*<br>+Metabolites*<br>+Lysophospholipids*<br>+Fatty acids* |
| Chlebowski (2020) | Normal | LF | *ns* MMSE | *NR* | *NR* | *NR* |
| Halyburton (2007) | Normal | KETO | *NR* | *ns* Working memory<br>*NR* Processing speed* | *NR* | +Hydroxybutyrate*<br>*NR* Total-C, LDL-C, HDL-C |
| Hardman (2020) | Normal | MEDI | *ns* SUCCAB composite | *ns* Memory composite<br>*ns* Simple reaction time<br>*ns* Choice reaction time<br>*ns* Immediate recognition<br>*ns* Delayed recognition<br>*ns* Congruent Stroop<br>*ns* Incongruent Stroop<br>*ns* Spatial working memory<br>*ns* Contextual memory | *NR* | *ns* Total-C<br>*ns* Fasting Glucose<br>*ns* hs-CRP<br>*ns* Vitamin B-12, D, B-6<br>*ns* HbA1c<br>*ns* IGF-I<br>*ns* BDNF<br>*ns* Homocysteine |
| Horie (2016) | MCI | CR | *ns* CAMCog<br>*ns* IQCODE | *ns* RAVLT<br>*ns* Executive Function | *NR* | *NR* CRP<br>*NR* Leptin<br>*NR* HOMA-IR<br>*NR* Lipids |
| Hoscheidt (2021) | Normal, MCI | MEDI | *ns* Cognitive composite | *NR* | +MRI whole brain CBF*(Normal)<br>+MRI L infer. Frontal cortex*(Normal)<br>+MRI R mid temporal gyrus*(Normal)<br>+MRI Parahippocampal gyri*(Normal)<br>+MRI Posterior cingulate*(Normal)<br>+MRI Precuneus*(Normal)<br>+MRI Hippocampus*(Normal)<br>+MRI Mean Cerebral Perfusion†(Normal)<br>*ns* MRI Mean Cerebral Perfusion(MCI) | +CSF Amyloid Beta*(Normal)<br>+AB42/40 Ratio*<br>+T-Tau*(MCI)<br>+AB42/Tau Ratio* (MCI)<br>+Total-C, LDL-C, HDL-C<br>*ns* insulin<br>*ns* Glucose<br>*ns* HbA1c |

| | | | | | | |
|---|---|---|---|---|---|---|
| Jakobsen (2011)<br>*Zellner (2011)* | Normal | HP | *ns* ACE Score | +Reaction time*<br>+ACE Orientation*<br>+Go/No-Go *<br>*ns* Verbal fluency<br>*ns* Language<br>*ns* Sustained attention | *NR* | +Vitamin B-12*<br>+MaoB*<br>*ns* Homocysteine<br>*ns* Vitamin B-9 (Folate)<br>*ns* Haemoglobin<br>*ns* TSH, $T_3$, $T_4$<br>*ns* IGF-1<br>*ns* Insulin<br>*ns* Glucose<br>*ns* Urea<br>*ns* Growth Hormone<br>*ns* Creatinine<br>*ns* Alkaline Phosphatase |
| James (2024) | Normal | IF | *ns* T-MoCA<br>*ns* E-COG-12 | *NR* | *NR* | *NR* |
| Jennings (2024) | Normal | MEDI | *NR* NTB | *NR* Verbal Memory*<br>*NR* Processing Speed<br>*NR* Executive Function<br>*NR* COWAT<br>*NR* Category Fluency Test<br>*NR* TMT B-A<br>*NR* Wechsler Memory Digit Span<br>*NR* Digit Symbol Substitution<br>*NR* TMT-A<br>*NR* VPA (Immediate)<br>*NR* VPA (Delayed)<br>*NR* RVLT | *NR* | *NR* |
| Keawtep (2024) | Normal | IF | *ns* MoCA | *ns* TMT B-A<br>*ns* Logical memory<br>*ns* SCWT<br>*ns* Verbal Fluency<br>*ns* Digit Span Test | NR | +Total-C*<br>+BDNF*<br>*ns* HOMA-IR,<br>*ns* insulin<br>*ns* Adiponectin<br>*ns* Glucose<br>*ns* Triglycerides<br>*ns* IL-6 |
| Knight (2016) | Normal | MEDI | *ns* Cognitive composite | *ns* Executive function<br>*ns* Memory<br>*ns* Processing speed<br>*ns* Visual-spatial score | *NR* | *NR* |

| | | | | | | |
|---|---|---|---|---|---|---|
| Komulainen (2021)<br>*Komulainen (2010)* | Normal | NNG | *ns* CERAD-TS<br>*ns* MMSE | *ns* Verbal fluency<br>*ns* Modified Boston Naming Test<br>*ns* Word list memory<br>*ns* Constructional praxis<br>*ns* Word list recall<br>*ns* Word list recognition | *NR* | *NR* |
| Krikorian (2012b) | MCI | KETO | *NR* | +VPAL†<br>*ns* TMT-B | NR | +Insulin†<br>+Fasting glucose† |
| Makris (2013) | Normal | KETO | *NR* | *ns* Stroop<br>*ns* CPT<br>*ns* Word recall<br>*ns* Wisconsin card-sorting task | *NR* | *NR* |
| Marseglia (2018) | Normal | MEDI | *ns* MMSE<br>*ns* CERAD-TS | *ns* Executive function<br>*ns* Perceptual speed<br>*ns* Episodic memory (Immediate)<br>*ns* Episodic memory (Delayed)<br>*ns* Verbal abilities<br>*ns* Constructional praxis | *NR* | *NR* |
| Martin (2007) | Normal | CR, LCD | *NR* | *ns* RAVLT<br>*ns* ACT<br>*ns* BVRT<br>*ns* CPT | *NR* | *NR* |
| Martinez-Lapiscina (2013a, *2013b)* | Normal, MCI | MEDI ± EVOO or Nuts | +MMSE†(EVOO)<br>+MMSE*(NUTS) | +CDT*(EVOO)<br>+CDT*(NUTS) | *NR* | *NR* |
| Napoli (2014) | Normal | CR | +3MS (MMSE)* | *ns* Word List Fluency<br>*ns* TMT-A<br>+TMT-B* | *NR* | *ns* hs-CRP<br>*ns* Insulin Sensitivity Index<br>*ns* IGF-1 |
| Rizvi (2024) | Normal | IF, CD | *NR* | *NR* MMQ memory | *NR* | +Total-C*(IF)<br>+Triglyceride*(IF)<br>+HDL-C*(IF)<br>*ns* LDL-C(IF, CD)<br>*ns* Total-C(CD)<br>*ns* Triglyceride(CD)<br>*ns* HDL-C(CD) |
| Silver (2023)<br>*Leclerc (2019)*<br>*Grigolon (2020)* | Normal | CR | *NR* | *ns* SWM | *NR* | *NR* |
| Smith (2010) | Normal | DASH | *ns* EFML Composite | +Psychomotor speed*<br>*ns* DSST | NR | NR |

| | | | | | | |
|---|---|---|---|---|---|---|
| | | | | +Ruff 2&7*<br>*ns* TMT B-A<br>*ns* VPA<br>*ns* COWAT<br>*ns* Stroop<br>*ns* Digit Span | | |
| Smith (2020) | MCI | DASH | *NR* | *NR* Executive Function<br>*NR* Trail Making Test<br>*NR* Stroop Test<br>*NR* Digit Span Forward<br>*NR* Digit Span Backwards<br>*NR* Digit Symbol Substitution Test<br>*NR* Ruff 2 & 7<br>*NR* Animal Naming Test<br>*NR* Memory<br>*NR* Language | *NR* | +Metabolic Composite*<br>*ns* IL-6<br>*ns* hs-CRP<br>*ns* BDNF<br>+HOMA-IR*<br>*ns* Leptin<br>*ns* IGF-1<br>*ns* VEGF |
| Tussing-Humphreys (2022) | MCI | MEDI | *NR* | *NR* AIP Composite<br>*NR* EF Composite<br>*NR* LMR Composite | *NR* | *ns* HOMA-IR<br>*ns* Fasting Insulin<br>*ns* HbA1c<br>*ns* Total-C, LDL-C, HDL-C<br>*ns* Triglycerides<br>*ns* hs-CRP<br>*ns* Glucose |
| Uchiyama-Tanaka (2024) | Normal | Low AGE ± MAF | +MCIS‡ | *NR* | *NR* | *ns* Aβ40/42<br>*NR* HbA1c<br>*NR* Alkaline Phosphatase<br>*NR* Total-C, HDL-C<br>*NR* Triglycerides<br>*NR* Urea<br>*NR* Creatine<br>*NR* Ferritin<br>*NR* Vitamin D<br>*NR* Vitamin B-12, B-1<br>*NR* TSH, $T_4$<br>*NR* Albumin<br>*NR* Zinc, Copper<br>*NR* Sodium, Potassium<br>*NR* Chloride<br>*NR* Folic Acid<br>*NR* GPT |

| | | | | | | |
|---|---|---|---|---|---|---|
| | | | | | | *NR* Uric Acid |
| Valls-Pedret (2015) | Normal | MEDI + EVOO or Nuts | *ns* MMSE<br>+Global Cognition composite†(EVOO) | +Frontal cognition composite† (EVOO)<br>+Memory composite*(NUTS)<br>+RAVLT Total Learning*(EVOO)<br>*ns* RAVLT Delayed Recall<br>*ns* Paired Associates<br>*ns* Verbal fluency<br>*ns* Digit span<br>*ns* Colour Trail Test (CTT) Part 1<br>+CTT Part 2*(EVOO | *NR* | *NR* |
| Wardle (2000) | Normal | LF, MEDI | *NR* | — Bakan sustained attention‡<br>*ns* Motor speed<br>*ns* Memory<br>*ns* Choice reaction time | NR | +Total-C*<br>+LDL-C*<br>*ns* Triglycerides<br>*ns* HDL-C |
| Zhu (2024) | Normal | HFD | *NR* | +PRMQ Memory Total*<br>*ns* BRIEF Executive function | *NR* | *NR* |

Abbreviations: 3MS: Modified Mini-Mental State Examination; Aβ42/40 Ratio: amyloid-beta 42 ($Aβ_{42}$) to amyloid-beta 40 ($Aβ_{40}$) peptides ; ACE: Adverse Childhood Experiences Score; ACT: Auditory Consonant Trigrams; AD: Alzheimer's Disease; ADAS-Cog: Alzheimer's Disease Assessment Scale – Cognitive Subscale; AGE: Advanced glycation end-products; AIP: Attention, Information, & Processing; AVLT: Auditory Verbal Learning Test; BDNF: Brain-Derived Neurotrophic Factor; BDST: Backward Digit Span Test; BRIEF: Behaviour Rating Inventory of Executive Function; BVRT: Benton Visual Retention Test; CAMCog: Cambridge Cognition Examination; CBF: Cerebral Blood Flow; CD: Custom Diet; CDR-SB: Clinical Dementia Rating – Sum of Boxes; CDT: Clock Drawing Test; CERAD-TS: Consortium to Establish a Registry for Alzheimer's Disease – Total Score; CNS: Central Nervous System; COWAT: Controlled Oral Word Association Test; CPT: Continuous Performance Test; CR: Caloric Restriction; CRP: C-Reactive Protein; CSF: Cerebrospinal Fluid; CTT: Colour Trail Test; DASH: Dietary Approaches to Stop Hypertension; DSST: Digit Symbol Substitution Test; EFML: Executive Function and Memory Learning Composite; E-COG-12: Everyday Cognition Scale – 12-item version; EVOO: Extra Virgin Olive Oil; FCSRT: Free and Cued Selective Reminding Test; FDST: Forward Digit Span Test; GFAP: Glial Fibrillary Acidic Protein; GPT: Glutamate Pyruvate Transaminase; HbA1c: Haemoglobin A1c; HDL-C: High-Density Lipoprotein Cholesterol; HF: High fat; HFD: High Fibre Diet; HOMA-IR: Homeostatic Model Assessment of Insulin Resistance; HP: High Protein; hs-CRP: High-sensitivity C-Reactive Protein; IF: Intermittent Fasting; IGF-I: Insulin-like Growth Factor I; IL-6: Interleukin 6; IQCODE: Informant Questionnaire on Cognitive Decline in the Elderly; Keto: Ketogenic diet; L: Left; LCD: Low Carbohydrate Diet; LDL-C: Low-Density Lipoprotein Cholesterol; LF: Low Fat; LMR: Learning Memory Recognition Composite; LNST: Letter Number Sequencing Test; MAF: Macrophage-Activating Factor; MCI: Mild Cognitive Impairment; MCIS: Memory Capacity Impairment Score; MEDI: Mediterranean Diet; MIND: Mediterranean-DASH Intervention for Neurodegenerative Delay; MMQ: Multifactorial Memory Questionnaire; MMSE: Mini-Mental State Examination; MoCA: Montreal Cognitive Assessment; MRI: Magnetic Resonance Imaging; NNG: National Nutrition Guidelines; ns: non-significant; NR: Not Reported; NTB: Neuropsychological Test Battery; pTau181: Phosphorylated Tau - 181; PRMQ: Prospective and Retrospective Memory Questionnaire; R: Right; RBANS: Repeatable Battery for the Assessment of Neuropsychological Status; RAVLT: Rey Auditory Verbal Learning Test; SCWT: Stroop Colour and Word Test; SUCCAB: Swinburne University Computerized Cognitive Assessment Battery; SWM: Spatial Working Memory; TC: Total Cholesterol; TMT-A: Trail Making Test – Part A; TMT-B: Trail Making Test – Part B; TMT B-A: Trail Making Test B minus A; TSH: Thyroid Stimulating Hormone, T3: Triiodothyronine, T4: Thyroxine; VEGF: Vascular Endothelial Growth Factor; VEGF: Vascular Endothelial Growth Factor VLDL-C: Very-Low-Density Lipoprotein Cholesterol; VPA: Verbal/Visual Paired Associates; VPAL: Verbal/Visual Paired Associates Learning

**Supplementary Table S6c.** Between-group differences in cognitive outcomes at post-intervention in single food interventions (*n* = 31)

| **Single Food Dietary Interventions (n = 31)** | | | | | | **+ = Favours intervention**<br>**— = Favours control**<br>***ns* = No significant effect/difference in CF**<br><br>***=p<0.05**<br>**†=p<0.01**<br>**‡=p<0.001** |
|---|---|---|---|---|---|---|
| Author (Year) | Cognitive Status | INT Type | Global Cognition | Single-Domain Cognition | Brain Imaging | Blood Biomarkers |
| Babateen (2022) | Normal | Fruit & Vegetables: Beetroot | *ns* COMPASS composite | *ns* Word presentation<br>*ns* IWR, delayed recall<br>*ns* Working memory<br>*ns* Choice reaction time<br>*ns* Stroop<br>*ns* Digit vigilance<br>*ns* Corsi blocks<br>*ns* Peg & ball<br>*ns* Word recognition<br>*ns* TMT-A, TMT-B | *ns* qNIRS CBF<br>*ns* qNIRS Oxyhaemoglobin<br>*ns* qNIRS Deoxyhaemoglobin<br>*ns* qNIRS Oxygen Saturation<br>*ns* qNIRS Total Haemoglobin | *NR* |
| Boespflug (2017) | MCI | Fruit & Vegetables: Blueberry | *NR* MoCA | *ns* working memory (n-back) | NR L pre-central gyrus<br>NR L post-central gyrus<br>NR L middle frontal gyrus<br>NR L inferior parietal lobe | *NR* |
| Bohn (2021) | At-risk | Fruits & Vegetables: Bilberry/ Grape | *NR* | *ns* CANTAB DMS<br>*ns* CANTAB PAL<br>*ns* CANTAB PRM<br>*ns* CANTAB SRM<br>*ns* Grooved Pegboard Test | *NR* | +Monocytes*<br>+Cytokines*<br>+Carotenoids*<br>+Polyphenols‡<br>+Oxidative stress*<br>+TNF*<br>+Interleukins (IL-6, 9,10)*<br>*ns* Fibroblast GF<br>+Epidermal GF*<br>+VEGF*<br>+Macrophage inflammatory protein |

| | | | | | | |
|---|---|---|---|---|---|---|
| | | | | | | *ns* Granulocyte-macrophage colony-stimulating factor<br>*ns* Eotaxin immunoprotein<br>*ns* Factalkine immunoprotein<br>*ns* IL-4, -7, -8, -12, -13, -17<br>*ns* Interferon<br>*ns* Glucose<br>*ns* Insulin<br>*ns* Total-C, HDL-C, LDL-C<br>*ns* Triglycerides<br>*ns* Homocysteine<br>*ns* CRP<br>*ns* Uric acid<br>*ns* Sodium<br>*ns* Potassium<br>*ns* Albumin<br>*ns* Creatinine<br>*ns* WBC<br>*ns* Glutamyl transferase |
| Bookheimer (2013) | At-risk | Fruits & Vegetables: Pomegranate | *NR* | +Buschke Total Recall*<br>+Buschke LT Retrieval* | +fMRI basal ganglia*<br>+fMRI thalamus*<br>+fMRI R occipital*<br>+fMRI L occipital*<br>+fMRI R fusiform*<br>+fMRI L fusiform*<br>+fMRI parahippocampal cortex*<br>+fMRI L inf frontal gyrus*<br>+fMRI L mid frontal gyrus*<br>+fMRI L temporal gyrus* | +TEAC‡<br>*NR* urolithin A-glucuronide |
| Bowtell (2017) | Normal | Fruits & Vegetables: Blueberry | *NR* | *ns* Groton maze task<br>*ns* Delayed memory recall<br>*ns* Shopping list task<br>*ns* Processing speed | +fMRI Brodmann area‡<br>+fMRI precuneus‡<br>+fMRI anterior cingulate‡<br>+fMRI insula‡<br>+fMRI thalamus‡<br>+fMRI parietal lobe*<br>+fMRI occipital lobes*<br>*ns* fMRI frontal lobes | *ns* hs-CRP<br>*ns* BDNF<br>*ns* Glutathione |

| | | | | | | |
|---|---|---|---|---|---|---|
| Cardoso (2016) | MCI | Nuts & Seeds: Brazil Nut | *ns* CERAD-TS | +Verbal fluency†<br>+Constructional praxis*<br>*ns* Boston naming test<br>*ns* Word list learning<br>*ns* Word list recall | *NR* | +Selenium*<br>+Glutathione†<br>+Antioxidant*<br>*NR* Total-C<br>*NR* LDL-C, HDL-C, VLDL-C<br>*NR* Malondialdehyde |
| Chai (2019) | Normal | Fruit & Vegetables: Cherry | *NR* | +SWM*<br>— Digit span‡<br>+PAL*<br>+Reaction time*<br>*ns* RVP | *NR* | *NR* |
| Chan (2017) | AD | Oils & Fats: Coconut Oil | *ns* MMSE | *ns* CDT score | *NR* | *NR* Triglycerides<br>*NR* Glucose<br>*NR* Aspartate aminotransferase<br>*NR* Alanine aminotransferase |
| Cheatham (2023) | Normal | Fruits & Vegetables: Blueberry | *NR* | +RVP†<br>*ns* SWM<br>*ns* PAL<br>*ns* Reaction time | +EEG frontal L region*<br>+EEG temporal L region*<br>*ns* EEG frontal R region<br>*ns* EEG midline region<br>*ns* EEG central region<br>*ns* EEG temporal R region | *NR* |
| Coates (2020) | Normal | Nuts & Seeds: Almond | *NR* | *ns* Attention reaction time composite<br>*ns* Attention accuracy composite<br>*ns* WM reaction time composite<br>*ns* WM accuracy composite<br>*ns* Long-term memory accuracy composite<br>+EF accuracy composite*<br>*ns* Simple reaction time<br>*ns* Choice reaction time<br>*ns* Four choice reaction time<br>*ns* RVP correct responses<br>*ns* RVP reaction time<br>*ns* RVP false alarms<br>*ns* Numeric WM accuracy<br>*ns* Numeric WM reaction time<br>*ns* N-Back correct responses<br>*ns* N-Back reaction time | *NR* | +Triglycerides†<br>*ns* Total-C, HDL-C, LDL-C<br>*ns* Total-C:HDL-C Ratio<br>*ns* Glucose<br>*ns* Insulin<br>*ns* hs-CRP<br>*ns* HOMA2-IR<br>*ns* alpha-tocopherol |

| | | | | | | |
|---|---|---|---|---|---|---|
| | | | | *ns* Corsi Blocks-span<br>*ns* Corsi Blocks reaction time<br>*ns* Peg and Ball planning time<br>*ns* Peg and Ball execution time<br>*ns* Peg and Ball errors<br>*ns* Immediate word recall<br>*ns* Delayed word recall<br>*ns* Word recognition<br>*ns* Word recognition reaction time<br>*ns* Picture recognition<br>*ns* Stroop congruent accuracy<br>+Stroop incongruent accuracy*<br>*ns* Stroop congruent reaction time<br>*ns* Stroop incongruent reaction time<br>*ns* Serial 3<br>*ns* Serial 3 accuracy<br>+Serial 7 number of responses*<br>+Serial 7 accuracy* | | |
| Curtis (2024) | MCI | Fruits & Vegetables: Elderberry | *ns* MMSE | *ns* HVLT<br>*ns* VPS<br>*ns* Rey-O CFT<br>*ns* Boston Naming Test | *NR* | *ns* LDHA |
| Handajani (2020) | MCI | Meat & Alternative Protein: Tempeh | *NR* MMSE | *NR* Boston Naming Test[†]<br>*NR* Verbal fluency<br>*NR* Immediate memory recall | *NR* | *ns* Uric acid |
| Kamoun (2024) | Normal | Nuts & Seeds: Walnut | +MoCA* | *NR* | *NR* | *NR* |
| Kimble (2022) | Normal | Fruits & Vegetables: Cherry | *NR* | +Digit vigilance*<br>*ns* RVP<br>*ns* Working memory | *ns* NIRS CBF | +Amino acids*<br>+Plasma Metabolome*<br>+Polyphenols*<br>+Quinic acid |
| Krikorian (2009) | MCI | Fruits & Vegetables: Grape | *NR* | +CVLT Learning*<br>*ns* CVLT recall<br>*ns* S-PAL | *NR* | *ns* Fasting glucose<br>— Fasting insulin* |
| Krikorian (2012a) | MCI | Fruits & Vegetables: Grape | *NR* | *ns* CVLT Learning<br>*ns* CVLT recognition* | +fMRI R sup. parietal cortex*<br>+fMRI R mid frontal cortex* | *ns* Fasting glucose<br>*ns* Fasting insulin |

| | | | | | | |
|---|---|---|---|---|---|---|
| Krikorian (2022) | At-risk | Fruits & Vegetables: Blueberry | *NR* | +COWAT lexical access[†]<br>*NR* CVLT recall errors*<br>*ns* CVLT learning<br>*ns* CVLT delayed recall<br>*ns* CVLT recognition<br>*ns* V-PAL<br>*ns* EMQ-Total<br>+EMQ forgetfulness*<br>+EMQ encoding* | *NR* | +Insulin*<br>*ns* Glucose<br>*ns* HOMA2-IR<br>*ns* HbA1c<br>*ns* Total-C, LDL-C, HDL-C<br>*ns* Triglycerides<br>*ns* Mitochondrial uncoupling |
| Krikorian (2023) | At-risk | Fruits & Vegetables: Strawberry | *NR* | *ns* EF composite<br>*ns* Lexical access composite<br>*ns* Verbal memory composite<br>*ns* Visuospatial memory composite<br>+CVLT intrusion errors* | *NR* | *ns* Fasting glucose<br>*ns* Fasting insulin<br>*ns* HOMA2-IR<br>*ns* HbA1c<br>*ns* TG/HDL ratio |
| Lee (2017) | MCI | Fruits & Vegetables: Grape | *NR* ADAS-Cog<br>*NR* MMSE | *ns* HVLT Immediate Memory<br>*ns* HVLT Delayed Memory<br>*ns* Benton Visual Retention<br>*ns* Rey-O CFT (delayed memory)<br>*ns* Rey-O CFT Visuospatial<br>*ns* Boston Naming Test<br>*ns* Letter Fluency<br>*ns* Category Fluency<br>*ns* Stroop Interference<br>*ns* TMT-A or TMT-B<br>*ns* WCST-64<br>*ns* WAIS-III Digital Symbol<br>*ns* WAIS-III Processing Speed<br>*ns* WAIS-III Block Design<br>*ns* WAIS-III Symbol Search<br>*ns* WAIS-III Spatial Span<br>*ns* WAIS-III L-N Sequencing<br>*ns* WAIS-III Digit Span<br>*ns* WTAR<br>*ns* WTAR Verbal IQ<br>*ns* MFQ Forgetting<br>*ns* MFQ Seriousness<br>*ns* MFQ Retrospective Function<br>*ns* MFQ Mnemonics Usage | +PET R sup. Parietal cortex*<br>*ns* L inf. lateral Cortex<br>*NR* R post cingulate cortex<br>*NR* L sup. posterolateral temporal cortex | *NR* |

| | | | | | | |
|---|---|---|---|---|---|---|
| Mazza (2018) | Normal | Oils & Fats:<br>EVOO | +ADAS-Cog*<br>*ns* MMSE | *ns* Verbal fluency | *NR* | +HDL-C*<br>*ns* Total-C, LDL-C<br>*ns* Glucose<br>*ns* Triglycerides |
| Miller (2018) | Normal | Fruits & Vegetables:<br>Blueberry | *NR* | *NR* CVLT repetition errors*<br>*NR* CVLT Recall<br>*NR* CVLT Recognition<br>*NR* TMT-A, TMT-B<br>*NR* ANT Reaction time<br>*NR* vMWM Spatial cognition<br>*NR* TST | *NR* | *NR* |
| Miller (2021) | Normal | Fruits & Vegetables:<br>Strawberry | *NR* | *NR* Virtual spatial navigation*<br>*ns* CVLT Word recognition*<br>*NR* vMWM Spatial cognition<br>*NR* CVLT Recall<br>*NR* DS<br>*NR* ANT<br>*NR* TST<br>*NR* TMT-A, TMT-B | *NR* | *NR* |
| Mirheidary (2019) | Normal | Fruits & Vegetables:<br>Grape | *NR* | + DS Acoustic Working Memory Score‡<br>*ns* DS Visual Working Memory Score<br>+ DS Acoustic Working Memory Span*<br>*ns* DS Visual Working Memory Span<br>+ N-Back Task Accuracy*<br>+ N-Back Task Reaction Time* | *NR* | *NR* |
| Parilli-Moser (2021) | Normal | Nuts & Seeds:<br>Peanuts (SRP or PB) | *NR* | *ns* VPA Immediate memory<br>*ns* DS<br>*ns* LNS<br>*ns* TMT-A<br>*ns* TMT-B<br>*ns* Semantic/Formal Fluency | *NR* | *ns* Glucose<br>*ns* Triglycerides<br>*ns* Total-C, LDL-C, HDL-C<br>*ns* CRP<br>*ns* VLCSFAs(SRP) |
| Rakic (2022) | Normal | Nuts & Seeds:<br>Almond | *NR* | *ns* Motor Screening Task<br>*ns* Delayed Matching to Sample<br>*ns* Spatial Span Forward<br>*ns* Spatial Span Reverse<br>*ns* Paired Associate Learning<br>*ns* Rapid Visual Processing<br>*ns* One Touch Stockings | *NR* | *ns* IL-6<br>*ns* CRP<br>*ns* sICAM-1<br>*ns* Antioxidants (glutathione)<br>*ns* Total-C, LDL-C, HDL-C, VLDL-C<br>*ns* Triglycerides<br>*ns* Tocopherols |

| | | | | | | |
|---|---|---|---|---|---|---|
| Reeder (2022) | Normal | Nuts & Seeds: Peanut | *ns* NCI | +Reaction time*<br>*ns* Composite memory<br>*ns* Verbal memory<br>*ns* Visual memory<br>*ns* Complex attention<br>*ns* Simple attention<br>*ns* Cognitive flexibility<br>*ns* Processing speed<br>*ns* Attention<br>*ns* Psychomotor speed<br>*ns* Executive function | *NR* | *NR* |
| Rodrigo-Gonzalo (2023) | Normal | Fruits & Vegetables: Raisin | +MOCA‡<br>*ns* MMSE | + MoCA Orientation*<br>+ MoCA Language*<br>+ MoCA Visuospatial†<br>*ns* MoCA other subdomains<br>+ MMSE Time Orientation*<br>*ns* MMSE other subdomains<br>+RAVLT-IR†<br>+RAVLT-DR†<br>ns RAVLT fluency | *NR* | *NR* |
| Rutledge (2021) | Normal | Fruits & Vegetables: Blueberry | *NR* | *NR* Task-switching<br>*NR* CVLT-II Verbal memory | *NR* | +Polyphenols‡<br>+Anthocyanins* |
| Sala-Vila (2020) | Normal | Nuts & Seeds: Walnut | *ns* Global cognitive composite | *ns* Memory<br>*ns* Language<br>*ns* Perception<br>*ns* Frontal function | *ns* fMRI cortical thickness<br>*ns* fMRI brain volume<br>*ns* fMRI activation<br>*ns* fMRI Brain perfusion<br>*ns* fMRI White matter hyperintensity | +Fatty acids‡ |
| Siddarth (2019) | MCI | Fruits & Vegetables: Pomegranate | *NR* | +BVMT-R Learning†<br>*ns* BVMT-R Total Recall<br>*ns* BVMT-R Delayed Recall<br>*ns* BVMT-R Learning<br>*ns* SRT Total Recall<br>*ns* SRT Delayed Recall<br>*ns* SRT CLTR | *NR* | *NR* |
| Wood (2023) | Normal | Fruits & Vegetables: Blueberry | *NR* | + RAVLT Immediate recall*<br>+RAVLT Delayed recall*<br>+ TST accuracy* | *ns* Doppler CBF | *ns* Polyphenols<br>*ns* Fasting Glucose<br>*ns* LDL-C, HDL-C |

| | | | | |
|---|---|---|---|---|
| | | | | *ns* Corsi blocks<br>*ns* Serials 3 & 7 Subtraction |

Abbreviations: AD: Alzheimer's Disease; ADAS-Cog: Alzheimer's Disease Assessment Scale – Cognitive Subscale; ANT: Attention Network Test; BDNF: Brain-Derived Neurotrophic Factor; BNT: Boston Naming Test; BVMT-R: Brief Visuospatial Memory Test – Revised; CBF: Cerebral Blood Flow; CDR: Clinical Dementia Rating; CDT: Clock Drawing Test; CERAD-TS: Consortium to Establish a Registry for Alzheimer's Disease – Total Score; CLTR: Consistent Long-Term Retrieval; COMPASS: Cognitive and Physical Performance Assessment Battery; COWAT: Controlled Oral Word Association Test; CRP: C-Reactive Protein; CVLT: California Verbal Learning Test; Doppler CBF: Doppler Cerebral Blood Flow ;DMS: Delayed Matching to Sample (CANTAB subtest); DS: Digit Span; EEG: Electroencephalogram; EF: Executive Function; EMQ: Everyday Memory Questionnaire; fMRI: Functional Magnetic Resonance Imaging; GF: Growth Factor; HDL-C: High-Density Lipoprotein Cholesterol; HbA1c: Haemoglobin A1c; HOMA2-IR: Homeostatic Model Assessment of Insulin Resistance (version 2); hs-CRP: High-sensitivity C-Reactive Protein; HVLT: Hopkins Verbal Learning Test; IL-6: Interleukin 6; IWR: Immediate Word Recall; L: Left; LDHA: Lactate Dehydrogenase A; LDL-C: Low-Density Lipoprotein Cholesterol; LNS: Letter-Number Sequencing; MCI: Mild Cognitive Impairment; MFQ: Memory Functioning Questionnaire; MMSE: Mini-Mental State Examination; MoCA: Montreal Cognitive Assessment; N-Back: N-Back Working Memory Task; NCI: Neurocognitive Index; NIRS: Near-Infrared Spectroscopy; NR: Not Reported; PAL: Paired Associates Learning; PB: Peanut Butter; PET: Positron Emission Tomography; PMT: Probabilistic Matching Test; PRM: Pattern Recognition Memory (CANTAB); R: Right; RAVLT: Rey Auditory Verbal Learning Test; Rey-O CFT: Rey-Osterreith Complex Figure Test; RVP: Rapid Visual Information Processing (CANTAB); S-PAL: Spatial Paired Associates Learning; SRM: Spatial Recognition Memory (CANTAB); SRP: Skin Roasted Peanuts; SRT: Simple Reaction Time; SWM: Spatial Working Memory (CANTAB); TEAC: Trolox Equivalent Antioxidant Capacity; TMT-A: Trail Making Test Part A; TMT-B: Trail Making Test Part B; TNF: Tumour Necrosis Factor; TST: Task-Switching Task; TG/HDL: Triglyceride to HDL-C Ratio; VEGF: Vascular Endothelial Growth Factor; VLDL-C: Very Low-Density Lipoprotein Cholesterol ; VLCSFAs: Very Long Chain Saturated Fatty Acids VPA: Verbal Paired Association; VPAL: Visual Paired Associates Learning; vMWM: Virtual Morris Water Maze; VPS: Visuospatial Problem Solving, WAIS-III: Wechsler Adult Intelligence Scale; WBC: White Blood Cell; WCST: Wisconsin Card-Sorting Test; WM: Working Memory; WTAR: Wechsler Test of Adult reading

**Supplementary Table S7.** Hedges' *g* pre-post change sensitivity analysis across correlation coefficients 0.10-0.90

| *r* | Overall (n= 35) | | Intervention Type | | | | | |
|---|---|---|---|---|---|---|---|---|
| | | | Multidomain (n= 15) | | Total Diet (n= 11) | | Single Food (n= 9) | |
| | *g* (95% CI) | $I^2$ | *g* (95% CI) | $I^2$ | *g* (95% CI) | $I^2$ | *g* (95% CI) | $I^2$ |
| 0.10 | 0.17 (0.09, 0.26) | 0.0% | 0.14 (0.05, 0.23) | 0.0% | 0.23 (0.05, 0.40) | 4.5% | 0.25 (-0.03, 0.52) | 0.0% |
| 0.20 | 0.18 (0.10, 0.27) | 1.8% | 0.15 (0.05, 0.24) | 0.0% | 0.24 (0.05, 0.42) | 18.3% | 0.25 (-0.01, 0.51) | 0.0% |
| 0.30 | 0.20 (0.11, 0.29) | 14.0% | 0.22 (0.08, 0.36) | 48.2% | 0.24 (0.07, 0.42) | 20.8% | 0.24 (0.00, 0.48) | 0.0% |
| 0.40 | 0.22 (0.13, 0.31) | 22.4% | 0.22 (0.08, 0.36) | 50.6% | 0.26 (0.08, 0.43) | 24.4% | 0.26 (-0.06, 0.59) | 43.8% |
| 0.50 | 0.25 (0.15, 0.36) | 40.9% | 0.25 (0.10, 0.41) | 67.5% | 0.27 (0.10, 0.44) | 30.6% | 0.27 (-0.13, 0.66) | 67.1% |
| 0.60 | 0.27 (0.16, 0.38) | 58.9% | 0.27 (0.11, 0.43) | 73.3% | 0.28 (0.11, 0.45) | 39.3% | 0.26 (-0.17, 0.70) | 78.3% |
| 0.70 | 0.29 (0.18, 0.41) | 73.8% | 0.28 (0.12, 0.45) | 83.2% | 0.31 (0.13, 0.48) | 50.2% | 0.26 (-0.21, 0.73) | 85.7% |
| 0.80 | 0.31 (0.18, 0.44) | 85.7% | 0.30 (0.13, 0.47) | 89.4% | 0.33 (0.14, 0.52) | 75.2% | 0.25 (-0.25, 0.75) | 90.6% |
| 0.90 | 0.33 (0.19, 0.47) | 94.0% | 0.31 (0.14, 0.49) | 94.7% | 0.37 (0.15, 0.59) | 90.5% | 0.25 (-0.28, 0.77) | 95.7% |

**Supplementary Table S8.** Meta-regression analysis of moderators of dietary intervention effects on global cognitive function

| Multidomain Interventions (*k* = 15) | | | | | | | |
|---|---|---|---|---|---|---|---|
| **Subgroup (Moderators)** | **Number of Comparisons** | **Regression Coefficients (SE)** | **95% CI** | **p Value** | **QM (p)** | **$I^2$** | **$R^2$** |
| Risk of Bias | | | | | | | |
| Low/ Moderate (R) | 10 | 0.33 (0.10) | 0.14 to 0.52 | < 0.001 | 2.24 (0.13) | 0.57 | 0.33 |
| High | 5 | -0.21 (0.14) | -0.49 to 0.07 | 0.13 | | | |
| Cognitive Status | | | | | | | |
| MCI/ AD (R) | 4 | 0.41 (0.16) | 0.09 to 0.73 | 0.01 | 1.28 (0.26) | 0.66 | 0.009 |
| Normal/ At-risk | 11 | -0.21 (0.19) | -0.58 to 0.15 | 0.26 | | | |
| Intervention Duration | | | | | | | |
| ≤ 12 weeks (R) | 4 | 0.59 (0.16) | 0.28 to 0.90 | < 0.001 | **15.36 (< 0.001)** | 0.0003 | 0.99 |
| 24-52 weeks | 5 | -0.22 (0.19) | -0.59 to 0.15 | 0.25 | | | |
| > 52 weeks | 6 | -0.52 (0.16) | -0.84 to -0.20 | 0.002 | | | |
| Comorbidity | | | | | | | |
| No (R) | 12 | 0.18 (0.07) | 0.04 to 0.31 | 0.009 | **4.24 (0.04)** | 0.52 | 0.51 |
| Yes | 3 | 0.45 (0.22) | 0.02 to 0.88 | 0.04 | | | |
| Control Condition | | | | | | | |
| Passive (R) | 13 | 0.20 (0.08) | 0.05 to 0.36 | 0.01 | 1.91 (0.17) | 0.63 | 0.18 |
| Active | 2 | 0.31 (0.22) | -0.13 to 0.74 | 0.17 | | | |
| Intervention Components | | | | | | | |
| Diet + 1, 2, 3 (R) | 9 | 0.33 (0.11) | 0.12 to 0.53 | 0.002 | 1.04 (0.31) | 0.69 | 0.00 |
| Diet + 4, 5, 6 | 6 | -0.17 (0.17) | -0.51 to 0.16 | 0.31 | | | |
| Guided Support | | | | | | | |
| No (R) | 1 | 0.27 (0.40) | -0.51 to 1.05 | 0.50 | 0.001 (0.97) | 0.71 | 0.00 |
| Yes | 14 | -0.01 (0.41) | -0.81 to 0.78 | 0.97 | | | |
| Cognitive Training | | | | | | | |
| No (R) | 4 | 0.28 (0.16) | -0.04 to 0.60 | 0.08 | 0.02 (0.89) | 0.71 | 0.00 |
| Yes | 11 | -0.03 (0.19) | -0.40 to 0.35 | 0.89 | | | |
| Exercise | | | | | | | |
| No (R) | 1 | 0.54 (0.33) | -0.11 to 1.19 | 0.11 | 0.83 (0.36) | 0.67 | 0.06 |
| Yes | 14 | -0.31 (0.34) | -0.97 to 0.36 | 0.36 | | | |
| Instrument | | | | | | | |
| MMSE (R) | 10 | 0.11 (0.04) | 0.02 to 0.19 | 0.01 | **5.87 (0.02)** | 0.00 | 1.00 |
| Other | 5 | 0.28 (0.11) | 0.05 to 0.50 | 0.02 | | | |

| Total Diet Interventions (*k* = 11) | | | | | | | | |
|---|---|---|---|---|---|---|---|---|
| **Subgroup (Moderators)** | | **Number of Comparisons** | **Regression Coefficients (SE)** | **95% CI** | **p Value** | **QM (p)** | **$I^2$** | **$R^2$** |
| Risk of Bias | | | | | | | | |
| | Low/ Moderate (R) | 6 | 0.51 (0.13) | 0.26 to 0.77 | < 0.001 | **7.41 (0.007)** | 0.0001 | 1.00 |
| | High | 5 | -0.41 (0.15) | -0.70 to -0.11 | < 0.001 | | | |
| Cognitive Status | | | | | | | | |
| | MCI/ AD (R) | 1 | 0.19 (0.42) | -0.64 to 1.02 | 0.65 | 0.05 (0.83) | 0.36 | 0.00 |
| | Normal/ At-risk | 10 | 0.09 (0.43) | -0.75 to 0.94 | 0.83 | | | |
| Intervention Duration | | | | | | | | |
| | ≤ 12 weeks (R) | 3 | 0.19 (0.22) | -0.25 to 0.62 | 0.40 | **6.35 (0.04)** | 0.03 | 0.93 |
| | 24-52 weeks | 5 | 0.30 (0.26) | -0.20 to 0.80 | 0.24 | | | |
| | > 52 weeks | 3 | -0.08 (0.24) | -0.54 to 0.38 | 0.73 | | | |
| Type of Diet | | | | | | | | |
| | National Guidelines (R) | 2 | 0.16 (0,13) | -0.10 to 0.41 | 0.23 | **10.88 (0.05)** | 0.22 | 0.35 |
| | Intermittent Fasting | 2 | -0.04 (0.25) | -0.53 to 0.44 | 0.86 | | | |
| | High Protein | 2 | 0.16 (0.36) | -0.55 to 0.86 | 0.66 | | | |
| | Caloric Restriction | 1 | 0.36 (0.33) | -0.29 to 1.02 | 0.28 | | | |
| | AGE | 2 | 1.25 (0.41) | 0.45 to 2.04 | 0.002 | | | |
| | MEDI | 2 | 0.05 (0.20) | -0.34 to 0.44 | 0.81 | | | |
| Comorbidity | | | | | | | | |
| | No (R) | 5 | 0.38 (0.18) | 0.02 to 0.74 | 0.04 | 0.27 (0.60) | 0.44 | 0.00 |
| | Yes | 6 | -0.12 (0.22) | -0.56 to 0.32 | 0.60 | | | |
| Control Condition | | | | | | | | |
| | Passive (R) | 7 | 0.33 (0.12) | 0.08 to 0.57 | 0.008 | 0.22 (0.64) | 0.40 | 0.00 |
| | Active | 4 | -0.10 (0.21) | -0.50 to 0.31 | 0.64 | | | |
| Guided Support | | | | | | | | |
| | No (R) | 3 | 0.17 (0.22) | -0.25 to 0.59 | 0.42 | 0.34 (0.56) | 0.40 | 0.00 |
| | Yes | 8 | 0.14 (0.24) | -0.33 to 0.61 | 0.56 | | | |
| Instrument | | | | | | | | |
| | MMSE (R) | 5 | 0.22 (0.11) | 0.02 to 0.43 | 0.04 | 0.81 (0.37) | 0.33 | 0.00 |
| | Other | 6 | 0.18 (0.20) | -0.21 to 0.57 | 0.37 | | | |
| Single Food Interventions (*k* = 9) | | | | | | | | |
| **Subgroup (Moderators)** | | **Number of Comparisons** | **Regression Coefficients (SE)** | **95% CI** | **p Value** | **QM (p)** | **$I^2$** | **$R^2$** |
| Risk of Bias | | | | | | | | |

| | | | | | | | | |
|---|---|---|---|---|---|---|---|---|
| | Low/ Moderate(R) | 2 | -0.01 (0.49) | -0.98 to 0.95 | 0.98 | 0.42 (0.52) | 0.74 | 0.00 |
| | High | 7 | 0.36 (0.55) | -0.72 to 1.43 | 0.52 | | | |
| Cognitive Status | | | | | | | | |
| | MCI/AD (R) | 4 | 0.08 (0.34) | -0.59 to 0.74 | 0.81 | 0.64 (0.42) | 0.77 | 0.00 |
| | Normal/ At-risk | 5 | 0.39 (0.48) | -0.56 to 1.33 | 0.43 | | | |
| Intervention Duration | | | | | | | | |
| | ≤ 12 weeks (R) | 2 | 0.80 (0.50) | -0.17 to 1.77 | 0.10 | 1.48 (0.22) | 0.74 | 0.00 |
| | 24-52 weeks | 7 | -0.68 (0.56) | -1.78 to 0.41 | 0.22 | | | |
| | > 52 weeks | 0 | - | - | - | | | |
| Intervention Content | | | | | | | | |
| | Fruits and Vegetables (R) | 3 | -0.38 (0.45) | -1.25 to 0.49 | 0.39 | 3.55 (0.31) | 0.72 | 0.00 |
| | Nuts and Seeds | 3 | 1.10 (0.63) | -0.13 to 2.33 | 0.08 | | | |
| | Oil and Fats | 1 | 0.54 (0.80) | -1.04 to 2.12 | 0.50 | | | |
| | Protein | 2 | 0.95 (0.67) | -0.37 to 2.28 | 0.16 | | | |
| Comorbidity | | | | | | | | |
| | No (R) | 9 | 0.27 (0.20) | -0.13 to 0.66 | 0.18 | - | - | - |
| | Yes | 0 | - | - | - | | | |
| Control Condition | | | | | | | | |
| | Passive (R) | 5 | -0.06 (0.26) | -0.57 to 0.44 | 0.80 | 3.63 (0.06) | 0.60 | 0.21 |
| | Active | 4 | 0.72 (0.38) | -0.02 to 1.45 | 0.06 | | | |
| Guided Support | | | | | | | | |
| | No (R) | 6 | 0.08 (0.29) | -0.49 to 0.66 | 0.78 | 1.13 (0.29) | 0.74 | 0.00 |
| | Yes | 3 | 0.52 (0.49) | -0.44 to 1.48 | 0.29 | | | |
| Instrument | | | | | | | | |
| | MMSE (R) | 5 | 0.22 (0.30) | -0.36 to 0.81 | 0.45 | 0.06 (0.81) | 0.75 | 0.00 |
| | Other | 4 | 0.12 (0.49) | -0.84 to 1.07 | 0.81 | | | |

*Note. (R)* = Reference. AD: Alzheimer's Disease; AGE: Advanced Glycation End Products; MCI: Mild Cognitive Impairment; MEDI: Mediterrranean Diet; MMSE: Mini-Mental State Examination.

**Supplementary Table S9a.** GRADE assessment for certainty of evidence for multidomain intervention arms (*k* = 15)

| Certainty assessment | | | | | | | № of patients | | Effect | | Certainty | Importance |
|---|---|---|---|---|---|---|---|---|---|---|---|---|
| **№ of studies** | **Study design** | **Risk of bias** | **Inconsistency** | **Indirectness** | **Imprecision** | **Other considerations** | **Multidomain Interventions** | **Comparison** | **Relative (95% CI)** | **Absolute (95% CI)** | | |
| **Global Cognitive Function (assessed with: MMSE, ADAS-Cog, etc.)** | | | | | | | | | | | | |
| 15 | randomised trials | not serious[a] | serious[b] | serious[c] | serious[d] | publication bias strongly suspected all plausible residual confounding would reduce the demonstrated effect[e] | 1736 | 1054 | - | SMD **0.25 SD higher** (0.1 higher to 0.41 higher) | ⨁◯◯◯ Very low[a,b,c,d,e] | CRITICAL |

**CI:** confidence interval; **SMD:** standardised mean difference

*Explanations*

a. Most information is from low/some risk (10/15); most issues with handling of missing data. No downgrade.
b. Downgraded one level for serious inconsistency due to substantial unexplained heterogeneity ($I^2$ = 67.5%)
c. Downgraded one level for serious indirectness due to variations in the intervention components and dosage across studies, as well as differences in follow-up time points
d. Overall CI does not cross no effect (0.1 to 0.41), but they do go above and below the MID (0.2); 11 of the 15 comparisons had CIs cross 0 indicating both harm and benefit (imprecision); total sample meets OIS. Downgrade 1 level.
e. Egger’s test was statistically significant (p = 0.003), and the funnel plot showed asymmetry; upgraded due to plausible residual confounding; no change.

**Supplementary Table S9b.** GRADE assessment for certainty of evidence for total diet intervention arms (*k* = 11)

| Certainty assessment | | | | | | | № of patients | | Effect | | Certainty | Importance |
|---|---|---|---|---|---|---|---|---|---|---|---|---|
| **№ of studies** | **Study design** | **Risk of bias** | **Inconsistency** | **Indirectness** | **Imprecision** | **Other considerations** | **Total Diet Interventions** | **Comparison** | **Relative (95% CI)** | **Absolute (95% CI)** | | |
| **Global Cognitive Function (assessed with: MMSE, ADAS-Cog, etc.)** | | | | | | | | | | | | |
| 11 | randomised trials | serious[a] | not serious[b] | serious[c] | serious[d] | publication bias strongly suspected all plausible residual confounding would reduce the demonstrated effect[e] | 625 | 468 | - | SMD **0.27 SD higher** (0.1 higher to 0.44 higher) | ⨁◯◯◯ Very low[a,b,c,d,e] | CRITICAL |

**CI:** confidence interval; **SMD:** standardised mean difference

*Explanations*

a. 5/11 studies = 46% (HIGH ROB) vs. 6/11 = 55% (Some ROB); issues with randomisation and allocation, and selective reporting.

b. There was low heterogeneity across studies (I2 = 30.6%). The Q-statistic was insignificant (Q(10) = 17.51, p = 0.06) suggesting minor possibility of variability in effect sizes across studies. Not downgraded.

c. Downgraded 1 level for variation in more than one PICO element. Intervention dosage/type varied; comparators varied; outcome measurement tools varied.

d. Overall CI does not cross line of no effect, but does span below and above the MID of 0.2. 0.1-0.44 - moderate width; some uncertainty of whether the intervention is beneficial 6/11 = 0.545 (crossed line of no effect - some imprecision) Total sample meets optimal information size; Results are based on few studies. Downgraded 1 level.

e. Egger's test indicated significant funnel plot asymmetry (t = 3.41, p = .008) suggesting potential for publication bias (Supplementary File Figure S7). Need to account for publication bias. Indicated plausible residual confounding. No overall change in level.

**Supplementary Table S9c.** GRADE assessment for certainty of evidence for single food intervention arms (*k* = 15)

| Certainty assessment | | | | | | | № of patients | | Effect | | Certainty | Importance |
|---|---|---|---|---|---|---|---|---|---|---|---|---|
| **№ of studies** | **Study design** | **Risk of bias** | **Inconsistency** | **Indirectness** | **Imprecision** | **Other considerations** | **Single Food Interventions** | **Comparison** | **Relative (95% CI)** | **Absolute (95% CI)** | | |
| **Global Cognitive Function (assessed with: MMSE, ADAS-Cog)** | | | | | | | | | | | | |
| 9[a] | randomised trials | very serious[b] | serious[c] | serious | very serious[d] | none | 221 | 188 | - | SMD **0.27 SD higher** (0.13 lower to 0.66 higher) | ⨁◯◯◯ Very low[b,c,d] | NOT IMPORTANT |

**CI:** confidence interval; **SMD:** standardised mean difference

*Explanations*

a. No evidence of publication bias.
b. over half (55.6%) of the comparisons were rated “high” risk of bias, and 41.7% showed “some concerns” of bias; Meta-Analysis had 7/9 comparisons (78%) (high ROB) and 2 (22%) SOME ROB.
c. Substantial heterogeneity was observed across studies (I2 = 67.1%; Q(8) = 21.11, p = 0.007) suggesting considerable between-study variability.
d. CIs cross line of no-effect in 7 of 9 comparisons and include both harm and benefit; underpowered sample (221 intervention; 188 control) increases uncertainty of precision; wide confidence intervals. Downgraded 2 levels.

| Study (Experimental Arm) | D1 | D2 | D3 | D4 | D5 | Overall |
|---|---|---|---|---|---|---|
| Andrieu et al., 2017 (Multi-domain intervention + PUFA) | + | + | X | + | + | X |
| Andrieu et al., 2017 (Multi-domain intervention + Placebo) | + | + | X | + | + | X |
| Blumenthal et al., 2019 (DASH + AE) | + | + | + | + | + | + |
| Blumenthal et al., 2020 (DASH + AE) | + | + | + | + | + | + |
| Brodaty et al., 2025 (Diet + Exercise + CT + MH) | - | + | + | + | + | - |
| Chatterjee et al., 2022 (MEDI + CBCT) | + | + | + | X | - | X |
| Chatterjee et al., 2022 (MEDI + CBCT + Exercise) | + | + | + | X | - | X |
| Chhetri et al., 2018 (Multi-domain intervention + Omega 3) | + | + | + | + | + | + |
| Chhetri et al., 2018 (Multi-domain intervention + Placebo) | + | + | + | + | + | + |
| Han et al. 2023 (Diet + Exercise + SM) | + | + | X | + | + | X |
| Hardman et al., 2020 (Diet + Exercise) | - | X | X | X | + | X |
| Jennings et al., 2024 (MEDI + PA) | X | + | + | + | + | X |
| Komulainen at al., 2010 (Diet + AE) | + | - | + | + | + | - |
| Komulainen at al., 2010 (Diet + RE) | + | - | + | + | + | - |
| Komulainen at al., 2021 (Diet + AE) | + | + | X | + | - | X |
| Komulainen at al., 2021 (Diet + RE) | + | + | X | + | - | X |
| Keawtep et al., 2024 (IF + Physical-Cognitive Exercise) | - | + | + | + | + | - |
| Lee et al., 2023 (Multi-domain intervention + Supplement) | + | + | + | + | + | + |
| Martin et al., 2007 (CR + Exercise) | - | + | + | + | - | - |
| Masley et al., 2008 (Diet + Exercise + Meditation) | - | - | + | X | - | X |
| McMaster et al., 2020 (MEDI + Exercise +Brain Training + Support) | - | + | + | + | + | - |
| Mendoza-Ruvalcaba et al., 2015 (Diet + Exercise + CF) | - | + | + | - | - | - |
| Moon et al., 2021 (Multi-domain intervention, facility-based) | + | + | - | + | + | - |
| Moon et al., 2021 (Multi-domain intervention, home-based) | + | + | - | + | + | - |
| Nakazeko et al., 2023 (COMB-FP + Exercise) | - | X | + | + | + | X |
| Napoli et al., 2014 (CR + Exercise) | - | + | + | + | + | - |
| Ngandu et al., 2015 (FINGER) | + | + | + | + | + | + |
| Ornish et al., 2024 (Vegan diet + Exercise + SM) | - | + | + | + | + | - |
| Roach et al., 2023 (COCOA) | - | + | + | + | + | - |
| Rosenberg et al., 2018 (FINGER) | + | + | + | + | + | + |
| Smith et al., 2010 (DASH + WM) | - | + | + | + | - | - |
| Smith et al., 2020 (DASH + AE) | - | + | + | + | + | - |
| Sakurai et al., 2024 (Diet + Exercise + CT) | - | + | + | + | + | - |
| Thunborg et al., 2024 (Diet + Exercise + CT + SS + RM) | - | + | + | + | + | - |
| Tussing-Humphreys et al., 2022 (MEDI + Exercise + CR) | + | + | + | + | + | + |
| Zhu et al., 2024 (FR + RS) | + | + | + | + | + | + |

D1: Bias arising from the randomization process
D2: Bias due to deviations from the intended interventions
D3: Bias due to missing outcome data
D4: Bias in measurement of the outcome
D5: Bias in selection of the reported result

**Supplementary Figure S1.** Risk of bias assessment for multidomain intervention arms

| Study (Experimental Arm) | D1 | D2 | D3 | D4 | D5 | Overall |
|---|---|---|---|---|---|---|
| Arjmand et al., 2022 (MIND) | - | - | + | + | + | - |
| Barnes et al., 2023 (MIND + CR) | - | + | + | + | + | - |
| Bartholomew et al., 2021 (IF) | - | x | + | x | + | x |
| Blondal et al., 2022 (Icelandic Nutrition) | - | + | + | + | + | - |
| Blumenthal et al., 2019 (DASH) | + | + | + | + | + | + |
| Blumenthal et al., 2020 (DASH) | + | + | + | + | + | + |
| Brandt et al., 2019 (AD) | - | x | x | + | - | x |
| Brinkworth et al., 2009 (VLC - HFER) | - | - | x | + | - | x |
| Buchholz et al., 2024 (AD) | - | + | x | + | + | x |
| Chlebowski et al., 2020 (LF) | - | + | + | + | - | - |
| Grigolon et al., 2020 (CR) | - | + | + | + | - | - |
| Halyburton et al., 2007 (LCHF) | - | + | - | x | - | x |
| Hardman et al., 2020 (MEDI) | - | x | x | + | - | x |
| Horie et al., 2016 (CR) | - | - | + | + | + | - |
| Hoscheidt et al., 2021 (MEDI - HC) | - | + | + | + | + | - |
| Hoscheidt et al., 2021 (MEDI - MCI) | - | + | + | + | + | - |
| Jakobsen et al., 2011 (HP) | + | + | + | + | - | - |
| James et al., 2024 (IF) | - | + | + | - | + | - |
| Jennings et al., 2024 (MEDI) | x | + | + | + | + | x |
| Keawtep et al., 2024 (IF) | - | + | + | + | + | - |
| Knight et al., 2016 (MEDI) | - | - | + | x | - | x |
| Komulainen et al., 2010 (FNR rec.) | + | + | + | + | - | - |
| Komulainen et al., 2021 (FNR rec.) | + | + | x | + | - | x |
| Krikorian et al., 2012 (KD) | - | + | + | - | - | x |
| Leclerc et al., 2019 (CR) | - | x | x | x | - | x |
| Makris et al., 2013 (KD + CR) | - | + | + | + | - | - |
| Marseglia et al., 2018 (MEDI) | - | + | + | + | + | - |
| Martin et al., 2007 (CR) | - | + | + | - | - | - |
| Martin et al., 2007 (LC) | - | + | + | - | - | - |
| Martin et al., 2007 (CR + Exercise) | - | + | + | - | - | - |
| Martinez-Lapiscina et al., 2013 (a, MEDI + EVOO) | + | - | + | + | - | - |
| Martinez-Lapiscina et al., 2013 (a, MEDI + Nuts) | + | - | + | + | - | - |
| Martinez-Lapiscina et al., 2013 (b, MEDI + EVOO) | + | x | x | + | - | x |
| Martinez-Lapiscina et al., 2013 (b, MEDI + Nuts) | + | x | x | + | - | x |
| Napoli et al., 2014 (CR) | - | + | + | + | + | - |
| Rizvi et al., 2024 (IF) | - | + | + | - | + | + |
| Silver et al., 2023 (CR) | - | + | x | + | + | x |
| Smith et al., 2010 (DASH) | - | + | + | + | - | - |
| Smith et al., 2020 (DASH) | - | + | + | + | - | - |
| Tussing-Humphreys et al., 2022 (MEDI) | + | + | + | + | + | + |
| Tussing-Humphreys et al., 2022 (MEDI + CR) | + | + | + | + | + | + |
| Uchiyama-Tanaka et al., 2024 (AGE, Group D) | + | + | + | - | - | - |
| Valls-Pedret et al., 2015 (MEDI + EVOO) | + | + | x | + | + | x |
| Valls-Pedret et al., 2015 (MEDI + Nuts) | + | + | x | + | + | x |
| Wardle et al., 2000 (MEDI) | + | - | + | + | - | - |
| Wardle et al., 2000 (LF) | + | - | + | + | - | - |
| Zellner et al., 2011 (HP) | - | - | + | + | - | - |
| Zhu et al., 2024 (FR) | + | - | + | + | + | - |

D1: Bias arising from the randomization process
D2: Bias due to deviations from the intended interventions
D3: Bias due to missing outcome data
D4: Bias in measurement of the outcome
D5: Bias in selection of the reported result

**Supplementary Figure S2.** Risk of bias assessment for total diet intervention arms

| Study (Experimental Arm) | D1 | D2 | D3 | D4 | D5 | Overall |
|---|---|---|---|---|---|---|
| Babateen et al., 2022 (High Beetroot) | X | - | + | + | + | X |
| Babateen et al., 2022 (Medium Beetroot) | - | - | + | + | + | - |
| Babateen et al., 2022 (Low Beetroot) | - | - | + | + | + | - |
| Boespflug et al., 2017 (Blueberry) | + | + | X | X | X | X |
| Bohn et al., 2021 (Grape Juice) | - | + | + | + | + | - |
| Bookheimer et al., 2013 (Pomegranate) | - | + | + | + | - | - |
| Bowtell et al., 2017 (Blueberry) | - | + | + | + | - | - |
| Cardoso et al., 2016 (Brazil Nut) | + | + | - | + | + | - |
| Chai et al., 2019 (Cherry Juice) | - | + | + | + | + | - |
| Chan et al., 2017 (Coconut Oil) | + | + | + | - | X | X |
| Cheatham et al., 2023 (Blueberry) | - | + | + | - | + | - |
| Coates et al., 2020 (Almond) | - | + | + | + | + | - |
| Curtis et al., 2024 (Eldberry) | - | - | + | + | + | - |
| Handajani et al., 2020 (Tempeh A, lower microorganism count) | X | - | X | X | X | X |
| Handajani et al., 2020 (Tempeh B, higher microorganism count) | X | - | X | X | X | X |
| Kamoun et al., 2024 (CT + Walnut) | - | X | + | - | - | X |
| Kimble et al., 2022 (Cherry Juice) | + | + | + | + | + | + |
| Krikorian et al., 2009 (Concord Grape) | - | + | + | - | + | - |
| Krikorian et al., 2012 (Concord Grape) | + | + | + | - | X | X |
| Krikorian et al., 2022 (Blueberry) | - | X | X | X | - | X |
| Krikorian et al., 2023 (Strawberry) | - | X | X | X | + | X |
| Lee et al., 2017 (Grape) | + | X | - | + | - | X |
| Mazza et al., 2018 (Extra Virgin Olive Oil) | - | X | X | + | - | X |
| Miller et al., 2018 (Blueberry) | + | - | + | + | + | - |
| Miller et al., 2021 (Strawberry) | + | X | + | X | + | X |
| Mirheidary et al., 2019 (Raisin) | X | X | + | - | - | X |
| Parilli-Moser et al., 2021 (Roasted Peanut) | X | X | X | X | + | X |
| Parilli-Moser et al., 2021 (Peanut Butter) | X | X | X | X | + | X |
| Rakic et al., 2022 (Almond, 3 oz) | X | - | + | + | + | X |
| Rakic et al., 2022 (Almond, 1.5 oz) | X | - | + | + | + | X |
| Reeder et al., 2022 (Peanut) | X | - | + | - | - | X |
| Rodrigo-Gonzalo et al., 2023 (Raisin) | X | + | + | - | + | X |
| Rutledge et al., 2021 (Blueberry) | - | X | + | + | + | X |
| Sala-Vila et al., 2020 (Walnut) | - | + | + | + | + | - |
| Siddarth et al., 2019 (Pomegranate) | - | + | + | + | + | - |
| Wood et al., 2023 (Blueberry) | - | - | + | + | + | - |

D1: Bias arising from the randomization process
D2: Bias due to deviations from the intended interventions
D3: Bias due to missing outcome data
D4: Bias in measurement of the outcome
D5: Bias in selection of the reported result

**Supplementary Figure S3.** Risk of bias assessment for single food intervention arms

| Study (Experimental Arm) | D1 | D1b | D2 | D3 | D4 | D5 | Overall |
|---|---|---|---|---|---|---|---|
| Chou et al., 2022 (MEDI-Game) | - | + | + | + | + | - | - |
| Koblinsky et al., 2022 (Diet + Exercise) | - | + | + | + | + | + | - |
| Liang et al., 2021 (Exercise + CT + NA + HE, PCD) | - | + | + | + | + | + | - |
| Liang et al., 2021 (Exercise + CT + NA + HE, CD) | - | + | + | + | + | + | - |
| Liang et al., 2021 (Exercise + CT + NA + HE, MTF) | - | + | + | + | + | + | - |
| Zülke et al., 2024 (Multi-domain intervention) | + | + | - | + | + | + | - |

D1: Bias arising from the randomization process
D1b: Bias arising from the timing of identification and recruitment of individual participants in relation to the timing of randomization
D2: Bias due to deviations from the intended interventions
D3: Bias due to missing outcome data
D4: Bias in measurement of the outcome
D5: Bias in selection of the reported result

**Supplementary Figure S4.** Risk of bias assessment for cluster randomised controlled trials (rcts)

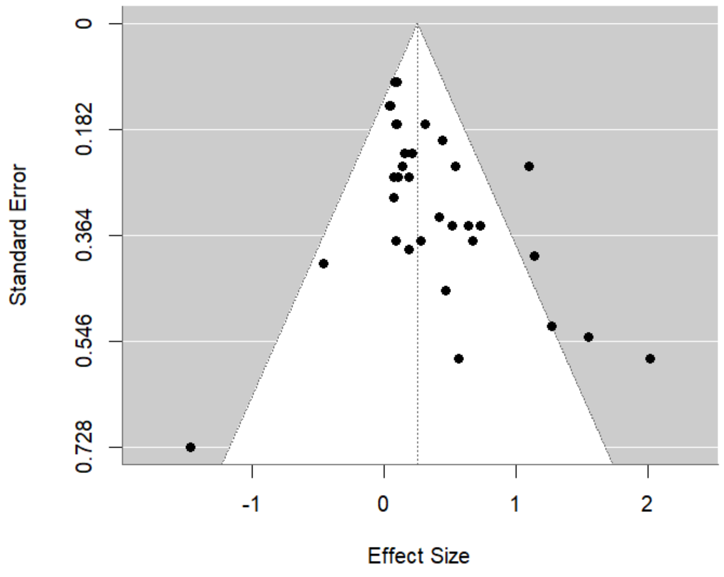

**Supplementary Figure S5.** Funnel plot of effect sizes versus standard errors for all studies included in the main meta-analysis

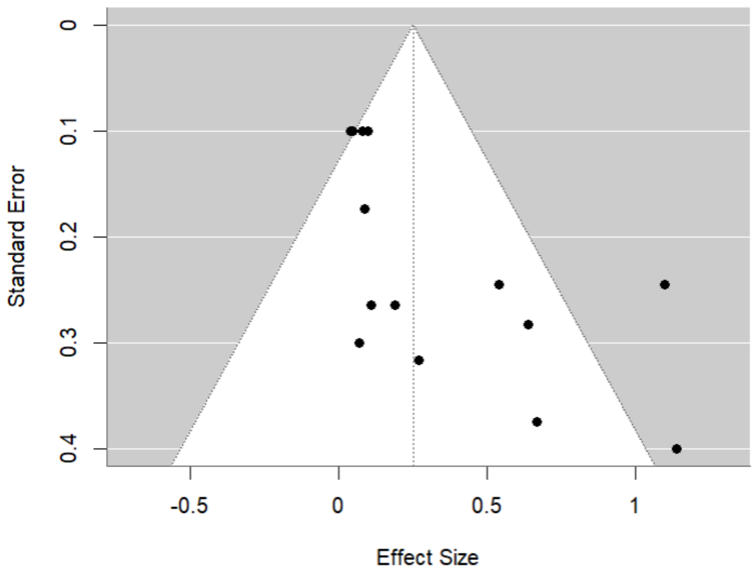

**Supplementary Figure S6.** Funnel plot of effect sizes versus standard errors for all multidomain studies included in the meta-analysis

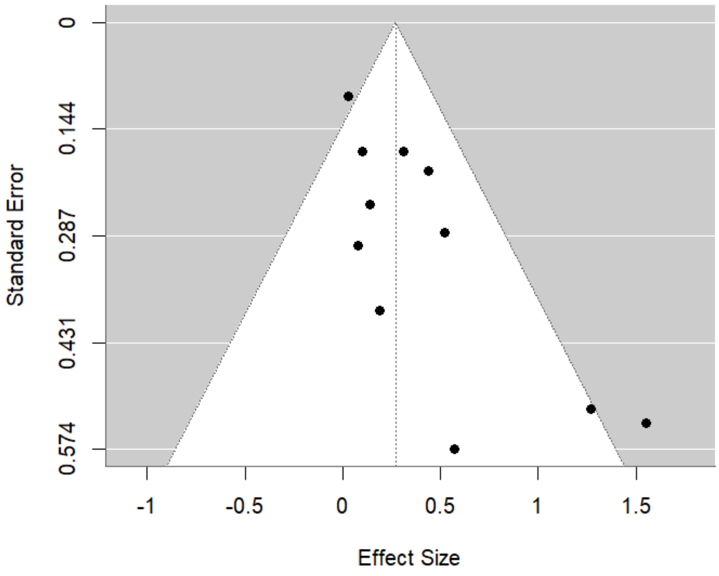

**Supplementary Figure S7.** Funnel plot of effect sizes versus standard errors for all total diet intervention studies included in the meta-analysis

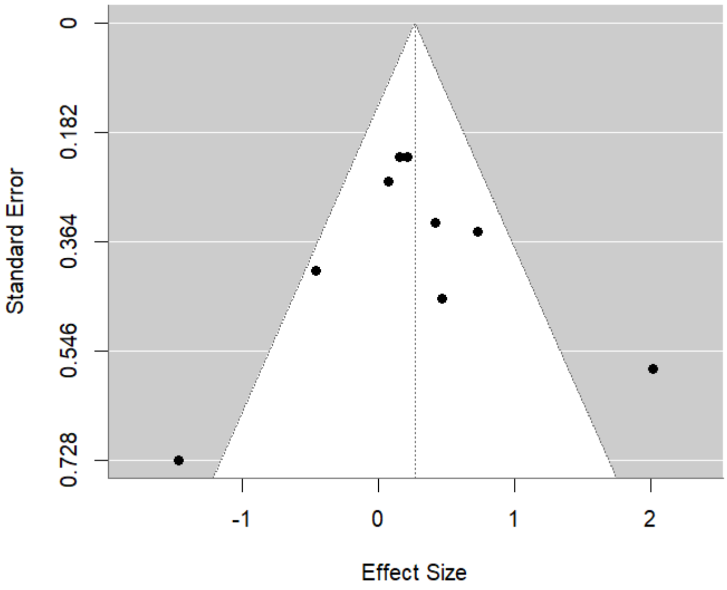

**Supplementary Figure S8.** Funnel plot of effect sizes versus standard errors for all single-food intervention studies included in the meta-analysis
